# Global and Regional Patterns of Parkinson’s Disease: Temporal Trends, Inequality Gaps, and Projections to 2040

**DOI:** 10.1101/2025.10.22.25338533

**Authors:** Kaleem Maqsood, Mahnoor Fatima, Humera Naveed, Wanhar Afzal, Zulfiqar Ali Beg, Fawaz Alhussain, Turki Abualait, Shahid Bashir

**Author notes:** Corresponding author **Dr Shahid Bashir**.

## Abstract

**Background:** Among the neurological disorders Parkinson deasease (PD) has emerged as one of the rapidly growing disorder, worldwide. This study is aimed to analyse the global, regional and national burden and trends of PD from 1990-2023, across the genders, age groups and sociodemographic context and forecasts of global trends through 2040.

**Methods:** For this study the data were sourced from the publically available most recent Global Burden of Disease (GBD) 1990-2023. The trends in age-standardized rates for key metrices including disability-adjusted life years (ASDR), mortality (ASMR), incidence (ASIR), and prevalence (ASPR) were analysed. Temporal trends were evaluated using estimated annual percentage changes (EAPCs). Associations with the Socio-Demographic Index (SDI) were explored using LOWESS regression. Future burden (2024–2040) was projected using an autoregressive integrated moving average (ARIMA) model.

**Results:** Globally, the ASDR increased from 77.71 to 88.62 per 100,000, ASMR from 4.27 to 4.85, ASIR from 11.92 to 15.11, and ASPR from 93.76 to 128.46. In 2023, PD accounted for 11.67 million cases and 0.42 million deaths. The greatest DALY increases occurred in low-middle SDI regions (33.23%), while high-middle SDI regions showed mild declines, which was also endorsed by LOWESS analysis. Regionally, the highest ASDR growth occurred in High-income North America (EAPC = 1.54%) and Central Sub-Saharan Africa (EAPC = 1.04%), whereas East Asia and Oceania showed declines. Prevalence rose most in North Africa and the Middle East (+97.74%; EAPC = 2.00). Nationally, the burden reduced in Kuwait and Qatar but rose sharply in Maldives and Ethiopia. PD burden increased exponentially with age and was consistently higher among males. Forecasts suggest continued global increases by 2040.

**Conclusions:** From 1990 to 2023, PD burden rose substantially worldwide, with the steepest increases in low- and middle-SDI regions. Despite stabilization in high-income areas, the global burden is projected to rise through 2040, underscoring the need for enhanced prevention, early diagnosis, and equitable access to long-term care.

## Introduction

Parkinson’s disease (PD) is a chronic, progressive neurodegenerative condition with motor (tremor and bradykinesia) [1] and non-motor (depression, cognitive impairment, and sleep difficulties) symptoms [2]. According to the Global Burden of Disease (GBD) Study 2015, PD is the neurological disease with the fastest-growing prevalence and disability [3]. The global prevalence of PD increased by 274%, from 3.15□million cases in 1990 to 11.77□million in 2021 [4]. To date, studies have projected the future global number of patients with PD upto 2040 based on GDB 2021 data; predictions of PD prevalence in most other countries and territories are still lacking, according to the 2023 data. This projection is largely driven by demographic aging, environmental exposure, and increased life expectancy, escalating a global public health challenge.

The incidence and duration of PD are influenced by a combination of biological and environmental factors [5]. Key biological determinants include age, with risk significantly increasing after 60 years, genetics (most cases are sporadic), sex [6], and ethnic/geographic variations [7]. Environmental factors contributing to PD risk include exposure to pesticides and air pollution, as well as other lifestyle factors such as physical inactivity and poor diet [8]. According to GBD study 2019, North Africa and the Middle East were shown to have highest PD incidence and disability burden, while Asia Pacific and Central Sub-Saharan Africa had highest death rates highlighting the geographic disparities in PD burden [9].

Though several studies assessed the Global Burden of PD, yet relied on GBD 2019 and GBD 2021 datasets. Further, an assessment using a more recent dataset will likely yield recent trends, new patterns of risk and updated regional estimates on the evolving epidemiology of the disease worldwide. With the GBD 2023 dataset now available, there is scope for more up-to-date and comprehensive evidence regarding the global and regional dynamics of PD burden.

This research study aims to use the GBD 2023 datasets to provide a detailed and up-to-date assessment of the burden of PD between 1990 and 2023. Specifically, it aims to study temporal trends in the following: age-standardized incidence, prevalence, mortality, and disability-adjusted life years of PD. The study will also look at trends by sex, age group, and SDI regions. It also aims at forecasting the PD trajectory up to 2040, including expected global and regional challenges at that time. This study will help health policymakers and clinicians to develop evidence-based strategies for preventing PD and optimally managing PD across the world by filling current data gaps and including updated estimates.

## Methodology

### Data Source

This study utilized the data from GBD database of 2023 study, which contain comprehensive information on Parkinson disease from 1990-2023. This database is developed by the Institute of Health Metrics and Evaluation, at University of Washington and provides comprehensive estimates of disease burden for 204 countries and territories across 21 regions [10]. The GBD 2023 uses mortality databases, national registries, hospital records, national surveys, scientific literature, health insurance databases etc. data from 1990-2023 [11]. For countries lacking data, statistical models and predictive algorithm were used to generate estimates, with validation conducted from an internal consistency check and external benchmarking.

### Study Design

This epidemiological study analyzed the trends in disease burden due to parkinson (PD) at global and regional levels from 1990 to 2023. The study aimed to investigate how the burden of PD evolved over time and by region during the study years and highlights the differences between countries-regions-age groups. Key metrics included: DALYs rate: A weighted composite of years lost due to disability per 100,000 people, reflecting the health loss attributable to PD. Mortality: The number of deaths due to PD per 100,000 people. Incidence: The number of new PD cases per 100,000 people. Prevalence: total number of PD cases per 100,000 people.

Additionally, the study considered data from the global outlook, the 21 GBD regions, and 204 countries and territories. To ensure the accuracy of the analysis, PD burdens were assessed across different age groups (0-4, 5-9, 10-14, 15-19, 20-24, 25-29, 30-34, 35-39, 40-44, 45-49, 50-54, 55-59, 60-64, 65-69, 70-74, 75-79, 80-84, 85-89, 90-94 and 95 years and above) [12]. This age stratification helped elucidate susceptibility variations, particularly among high-risk populations such as children and the elderly.

### Statistical Analysis

#### Descriptive Statistics

The study used multiple descriptive methods to present trends in DALYs mortality incidences and prevalence rates of PD globally regionally and nationally from 1990–2023. In addition to the calculated estimated values of each metric corresponding 95% uncertainty intervals were also presented to indicate estimation uncertainties. They also grouped and summarized patterns based on sex, age, and socioeconomic development index (SDI) assembly.

#### Trend Analysis

The EAPC was employed for studying the trends of age-standardized rates for temporal analysis of 1990-2023. To calculate this metric, the natural logarithm of age-standardized rates over time was analyzed using a linear regression model. The formula applied was y = a + bx + E where b positive indicates that the trends of age-standardized rates are increasing and b negative that they are decreasing. The confidence interval was determined at 95% using a formula 100 [exp(b) – 1]. The interpretation of EAPC trends was based on its 95% CI: If the 95% CI included 0, the disease burden was considered stable over time. If b > 0 and the lower bound of the CI (LCI) > 0, the disease burden was considered to be increasing. If b < 0 and the upper bound of the CI (UCI) < 0, the disease burden was interpreted as decreasing.

#### Forecasting

The expected development of PD in the next 17 years (2024-2040) was forecasted for ASDR, ASMR, ASIR and ASPR in India using ARIMA model. The ARIMA model incorporates autoregression (AR), differencing (I) and moving average (MA) that make it capable of capturing the time series pattern and seasonality. The Augmented Dickey-Fuller (ADF) test was used to assess the stationarity, while the autocorrelation function (ACF) and the partial autocorrelation function (PACF) were used to determine optimal values of p and q. The Ljung–Box Q test confirms that the selected models’ residuals are independent and normally distributed.

Statistical analyses and graphical presentations were made using R software (version 4.5.1), with results rounded to two decimal places for consistency.

## RESULTS

### Global Trends

Globally, the age-standardized DALY rate (ASDR) for PD increased from 77.71 per 100,000 in 1990 to 88.62 in 2023, showing a 14.04% rise (EAPC=0.33) (Table 1; Fig 1A). Similarly, the global age-standardized mortality rate (ASMR) also increased from 4.27 to 4.85 per 100,000, representing a 13.43% rise (EAPC= 0.33) (Table 2; Fig 1B). The absolute number of deaths reached 427,104 in 2023. The age-standardized incidence rate (ASIR) rose globally from 11.92 to 15.11 per 100,000, with an overall 26.77% increase (EAPC= 0.73) (Table 3; Fig 1C) and prevalence rate (ASPR) increased from 93.76 to 128.46 per 100,000, showing a 37.01% rise (EAPC: 0.94). Total global cases reached 11.67 million in 2023 (Table 4; Fig 1D).

**Table 1:** Global and regional DALYs data related to Parkinson disease. It includes location-based total numbers, age-standardized rates (ASR) per 100,000 for 1990 and 2023, percentage changes, and EAPC from 1990 to 2023.

| Regions | ASDR, 1990 (95% CI) | ASDR, 2023 (95% CI) | Numbers, 2023 | PC 1990-2023 | EAPC, %, 1990-2023 |
| --- | --- | --- | --- | --- | --- |
| Global | 77.71(71.00,84.65) | 88.62(81.09,97.91) | 7979498 | 14.04(6.33,21.65) | 0.33(0.28,0.38) |
| High-middle SDI | 86.20(77.04,97.49) | 83.41(74.35,95.99) | 1632029 | -3.24(-13.98,8.27) | -0.31(-0.39,-0.23) |
| Middle SDI | 74.10(65.75,82.43) | 88.85(78.80,101.99) | 675417 | 19.91(5.94,34.26) | 0.35(0.29,0.40) |
| Low-middle SDI | 67.98(56.75,78.87) | 90.57(77.05,105.40) | 723913 | 33.23(12.61,57.27) | 0.93(0.88,0.98) |
| Low SDI | 67.15(54.76,80.31) | 93.34(76.66,109.71) | 692981 | 39.00(17.62,68.94) | 0.87(0.79,0.95) |
| Andean Latin America | 72.71(65.44,80.68) | 109.04(97.14,122.73) | 67615 | 49.96(35.61,69.22) | 0.73(0.50,0.97) |
| Australasia | 83.51(76.41,90.04) | 86.13(77.72,95.22) | 56082 | 3.14(-3.08,9.96) | 0.15(0.03,0.27) |
| Caribbean | 72.46(66.79,78.51) | 96.86(88.64,106.51) | 54421 | 33.69(26.09,41.22) | 0.73(0.66,0.80) |
| Central Asia | 64.37(59.42,69.29) | 70.85(64.91,76.24) | 49436 | 10.07(0.58,18.29) | 0.43(0.32,0.55) |
| Central Europe | 80.86(76.16,85.26) | 87.11(80.43,93.15) | 216194 | 7.73(3.96,11.07) | 0.22(0.15,0.28) |
| Central Latin America | 80.72(73.82,87.47) | 88.30(81.07,96.48) | 223899 | 9.39(4.52,14.27) | 0.00(-0.10,0.10) |
| Central Sub-Saharan Africa | 49.55(38.45,61.14) | 71.66(54.49,86.57) | 33030 | 44.64(17.67,80.38) | 1.04(0.84,1.24) |
| East Asia | 92.40(80.94,107.73) | 78.91(68.74,92.10) | 1860899 | -14.59(-28.14,1.04) | -0.85(-1.01,-0.68) |
| Eastern Europe | 60.96(56.75,64.68) | 70.09(64.31,75.31) | 265171 | 14.98(7.73,22.99) | 0.53(0.43,0.64) |
| Eastern Sub-Saharan Africa | 56.77(43.27,70.38) | 83.80(65.69,100.90) | 129803 | 47.60(22.39,83.47) | 1.13(1.02,1.23) |
| High-income Asia Pacific | 57.30(51.96,62.15) | 71.07(62.11,78.26) | 416495 | 24.02(14.47,32.99) | 0.69(0.62,0.77) |
| High-income North America | 78.56(69.12,86.67) | 109.09(97.61,118.11) | 801398 | 38.86(31.10,47.02) | 1.54(1.46,1.63) |
| North Africa and Middle East | 90.31(76.92,104.34) | 111.52(96.81,128.82) | 437427 | 23.49(4.44,49.13) | 0.28(0.19,0.37) |
| Oceania | 104.70(83.27,127.34) | 111.87(90.61,133.19) | 6829 | 6.85(-10.29,27.41) | -0.22(-0.33,-0.12) |
| South Asia | 69.99(54.62,83.89) | 94.03(77.57,112.20) | 1225110 | 34.33(7.72,67.57) | 0.88(0.80,0.95) |
| Southeast Asia | 64.64(54.94,76.23) | 81.23(70.31,96.26) | 505808 | 25.66(6.54,46.28) | 0.46(0.42,0.51) |
| Southern Latin America | 93.07(86.44,99.17) | 100.01(90.93,107.77) | 100183 | 7.45(2.48,13.11) | -0.02(-0.11,0.08) |
| Southern Sub-Saharan Africa | 49.82(42.58,56.99) | 60.39(52.77,69.03) | 34907 | 21.22(2.78,41.69) | 0.35(0.10,0.61) |
| Tropical Latin America | 89.75(79.08,101.52) | 102.09(87.54,116.03) | 266583 | 13.74(9.81,18.27) | 0.34(0.30,0.39) |
| Western Europe | 76.96(70.52,82.04) | 95.84(85.24,104.40) | 1087547 | 24.52(20.47,28.53) | 0.63(0.57,0.69) |
| Western Sub-Saharan Africa | 65.16(53.99,78.25) | 85.62(71.98,102.57) | 140661 | 31.41(8.56,60.85) | 0.77(0.66,0.87) |

**Table 2:** Global and regional deaths data related to Parkinson disease. It includes location-based total numbers, age-standardized rates (ASR) per 100,000 for 1990 and 2023, percentage changes, and EAPC from 1990 to 2023.

| Regions | ASMR, 1990 (95% CI) | ASMR, 2023 (95% CI) | Numbers, 2023 | PC 1990-2023 | EAPC, %, 1990-2023 |
| --- | --- | --- | --- | --- | --- |
| Global | 4.27(3.90,4.68) | 4.85(4.29,5.34) | 427104 | 13.43(5.16,22.69) | 0.33(0.28,0.38) |
| High-middle SDI | 4.55(4.03,5.33) | 4.35(3.79,5.08) | 81173 | -4.48(-17.08,9.69) | -0.31(-0.39,-0.23) |
| Middle SDI | 4.00(3.52,4.63) | 4.63(4.01,5.30) | 31891 | 15.74(-0.50,33.00) | 0.35(0.29,0.40) |
| Low-middle SDI | 4.02(3.33,4.78) | 5.39(4.42,6.36) | 38830 | 34.31(10.82,63.26) | 0.93(0.88,0.98) |
| Low SDI | 4.10(3.22,5.01) | 5.72(4.57,6.85) | 37906 | 39.49(15.39,73.94) | 0.87(0.79,0.95) |
| Andean Latin America | 3.84(3.41,4.35) | 5.91(5.22,6.62) | 3560 | 53.92(33.64,76.92) | 0.73(0.50,0.97) |
| Australasia | 4.53(4.17,4.76) | 4.71(4.10,5.07) | 3212 | 3.95(-2.75,9.68) | 0.15(0.03,0.27) |
| Caribbean | 3.84(3.58,4.05) | 5.02(4.55,5.43) | 2810 | 30.96(21.28,40.06) | 0.73(0.66,0.80) |
| Central Asia | 4.16(3.80,4.52) | 4.57(4.11,4.90) | 2907 | 9.85(-0.57,19.39) | 0.43(0.32,0.55) |
| Central Europe | 4.71(4.44,4.90) | 5.02(4.65,5.25) | 12471 | 6.67(2.25,10.24) | 0.22(0.15,0.28) |
| Central Latin America | 4.30(3.99,4.52) | 4.57(4.24,4.84) | 11183 | 6.34(1.12,12.62) | 0.00(-0.10,0.10) |
| Central Sub-Saharan Africa | 2.95(2.23,3.74) | 4.21(3.09,5.23) | 1678 | 42.50(14.68,84.14) | 1.04(0.84,1.24) |
| East Asia | 4.69(4.02,5.66) | 3.84(3.21,4.55) | 86898 | -18.27(-34.10,0.53) | -0.85(-1.01,-0.68) |
| Eastern Europe | 3.65(3.36,3.91) | 4.30(3.88,4.61) | 15967 | 17.81(9.18,27.43) | 0.53(0.43,0.64) |
| Eastern Sub-Saharan Africa | 3.58(2.68,4.45) | 5.23(4.01,6.31) | 7499 | 46.11(18.34,87.74) | 1.13(1.02,1.23) |
| High-income Asia Pacific | 3.30(2.93,3.56) | 4.01(3.33,4.44) | 26621 | 21.76(9.91,32.92) | 0.69(0.62,0.77) |
| High-income North America | 3.92(3.47,4.15) | 6.29(5.46,6.77) | 47050 | 60.52(53.36,68.80) | 1.54(1.46,1.63) |
| North Africa and Middle East | 5.11(4.28,6.19) | 5.84(4.93,6.79) | 19841 | 14.26(-7.38,45.12) | 0.28(0.19,0.37) |
| Oceania | 5.41(4.13,6.75) | 5.65(4.35,7.00) | 290 | 4.48(-14.97,28.86) | -0.22(-0.33,-0.12) |
| South Asia | 4.24(3.25,5.22) | 5.83(4.68,7.15) | 68109 | 37.34(7.07,76.20) | 0.88(0.80,0.95) |
| Southeast Asia | 3.61(2.99,4.46) | 4.36(3.68,5.31) | 24697 | 20.86(-0.07,44.00) | 0.46(0.42,0.51) |
| Southern Latin America | 5.36(4.97,5.62) | 5.62(5.08,5.99) | 5754 | 4.81(-0.54,10.27) | -0.02(-0.11,0.08) |
| Southern Sub-Saharan Africa | 2.95(2.42,3.47) | 3.48(2.96,4.00) | 1880 | 17.91(-2.40,41.38) | 0.35(0.10,0.61) |
| Tropical Latin America | 4.13(3.73,4.38) | 4.38(3.79,4.71) | 11163 | 6.24(0.08,11.77) | 0.34(0.30,0.39) |
| Western Europe | 4.36(3.96,4.56) | 5.31(4.62,5.69) | 65574 | 21.80(16.37,26.23) | 0.63(0.57,0.69) |
| Western Sub-Saharan Africa | 4.08(3.30,4.97) | 5.37(4.36,6.52) | 7939 | 31.47(6.50,64.77) | 0.77(0.66,0.87) |

**Table 3:** Global and regional incidence data related to Parkinson disease. It includes location-based total numbers, age-standardized rates (ASR) per 100,000 for 1990 and 2023, percentage changes, and EAPC from 1990 to 2023.

| Regions | ASIR, 1990 (95% CI) | ASIR, 2023 (95% CI) | Numbers, 2023 | PC 1990-2023 | EAPC, %, 1990-2023 |
| --- | --- | --- | --- | --- | --- |
| Global | 11.92(10.54,13.42) | 15.11(13.59,16.85) | 1383388 | 26.77(22.88,31.07) | 0.73(0.71,0.75) |
| High-middle SDI | 12.49(10.76,14.39) | 15.45(13.51,17.69) | 312634 | 23.69(19.14,29.26) | 0.72(0.67,0.77) |
| Middle SDI | 10.96(9.66,12.48) | 15.20(13.44,17.27) | 123979 | 38.66(33.84,44.37) | 1.00(0.96,1.04) |
| Low-middle SDI | 8.41(7.37,9.50) | 11.25(10.01,12.74) | 96619 | 33.79(28.76,39.03) | 0.87(0.81,0.93) |
| Low SDI | 8.06(7.23,8.94) | 9.94(8.96,11.11) | 79325 | 23.31(18.91,28.00) | 0.67(0.65,0.68) |
| Andean Latin America | 14.35(12.46,16.20) | 20.87(18.27,23.35) | 13321 | 45.49(33.88,60.27) | 1.11(1.03,1.18) |
| Australasia | 15.13(14.12,16.22) | 17.09(14.96,19.72) | 10419 | 13.00(1.92,28.32) | 0.26(0.02,0.51) |
| Caribbean | 13.01(11.75,14.51) | 19.12(17.03,21.82) | 10800 | 46.95(40.45,55.22) | 1.16(1.00,1.33) |
| Central Asia | 6.05(5.70,6.46) | 6.83(6.40,7.31) | 5166 | 12.97(9.68,16.81) | 0.42(0.39,0.45) |
| Central Europe | 11.45(10.44,12.54) | 13.61(12.87,14.40) | 32968 | 18.89(13.03,24.51) | 0.36(0.30,0.42) |
| Central Latin America | 15.23(13.51,17.09) | 17.97(16.18,20.02) | 47204 | 17.96(13.66,22.30) | 0.44(0.39,0.50) |
| Central Sub-Saharan Africa | 6.60(5.96,7.35) | 7.74(7.06,8.57) | 3767 | 17.35(9.76,28.23) | 0.56(0.54,0.58) |
| East Asia | 13.25(11.24,15.55) | 16.40(14.07,18.97) | 395943 | 23.82(17.73,30.99) | 0.87(0.78,0.95) |
| Eastern Europe | 7.80(6.70,9.01) | 8.53(7.42,9.70) | 32530 | 9.34(4.47,14.59) | 0.32(0.27,0.36) |
| Eastern Sub-Saharan Africa | 7.43(6.61,8.29) | 8.63(7.81,9.56) | 13878 | 16.09(12.21,20.88) | 0.49(0.45,0.53) |
| High-income Asia Pacific | 8.05(6.90,9.32) | 11.86(10.64,13.28) | 61189 | 47.38(39.53,57.08) | 0.79(0.33,1.26) |
| High-income North America | 17.53(15.06,20.14) | 18.88(16.99,20.76) | 135861 | 7.71(-2.09,18.44) | 0.13(-0.05,0.31) |
| North Africa and Middle East | 13.31(11.74,15.30) | 21.78(19.10,24.64) | 97867 | 63.63(52.77,77.28) | 1.47(1.40,1.53) |
| Oceania | 14.80(13.10,16.80) | 18.00(15.96,20.72) | 1197 | 21.58(12.08,32.44) | 0.47(0.44,0.51) |
| South Asia | 7.83(6.69,9.01) | 10.07(8.82,11.43) | 140840 | 28.54(23.05,34.44) | 0.79(0.75,0.82) |
| Southeast Asia | 8.46(7.53,9.49) | 11.91(10.64,13.38) | 78632 | 40.80(35.20,47.63) | 1.11(1.08,1.15) |
| Southern Latin America | 14.60(13.43,15.81) | 18.23(16.76,19.37) | 17799 | 24.92(16.15,35.78) | 0.50(0.23,0.78) |
| Southern Sub-Saharan Africa | 7.25(6.23,8.28) | 9.03(7.97,10.30) | 5294 | 24.59(19.78,30.53) | 0.74(0.69,0.80) |
| Tropical Latin America | 23.43(19.88,27.76) | 29.77(25.23,35.08) | 80219 | 27.06(20.79,33.99) | 0.14(-0.08,0.36) |
| Western Europe | 13.45(12.39,14.52) | 17.91(16.79,19.22) | 183537 | 33.17(28.56,37.67) | 0.91(0.86,0.96) |
| Western Sub-Saharan Africa | 7.17(6.34,8.06) | 8.75(7.93,9.76) | 14956 | 22.17(17.34,27.12) | 0.66(0.62,0.70) |

**Table 4:**
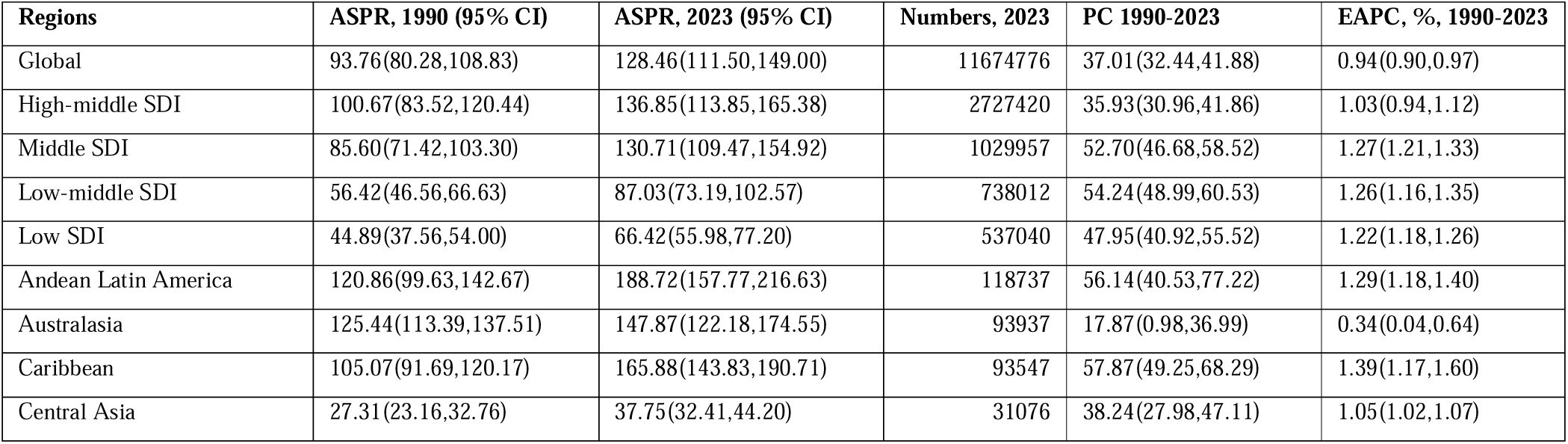
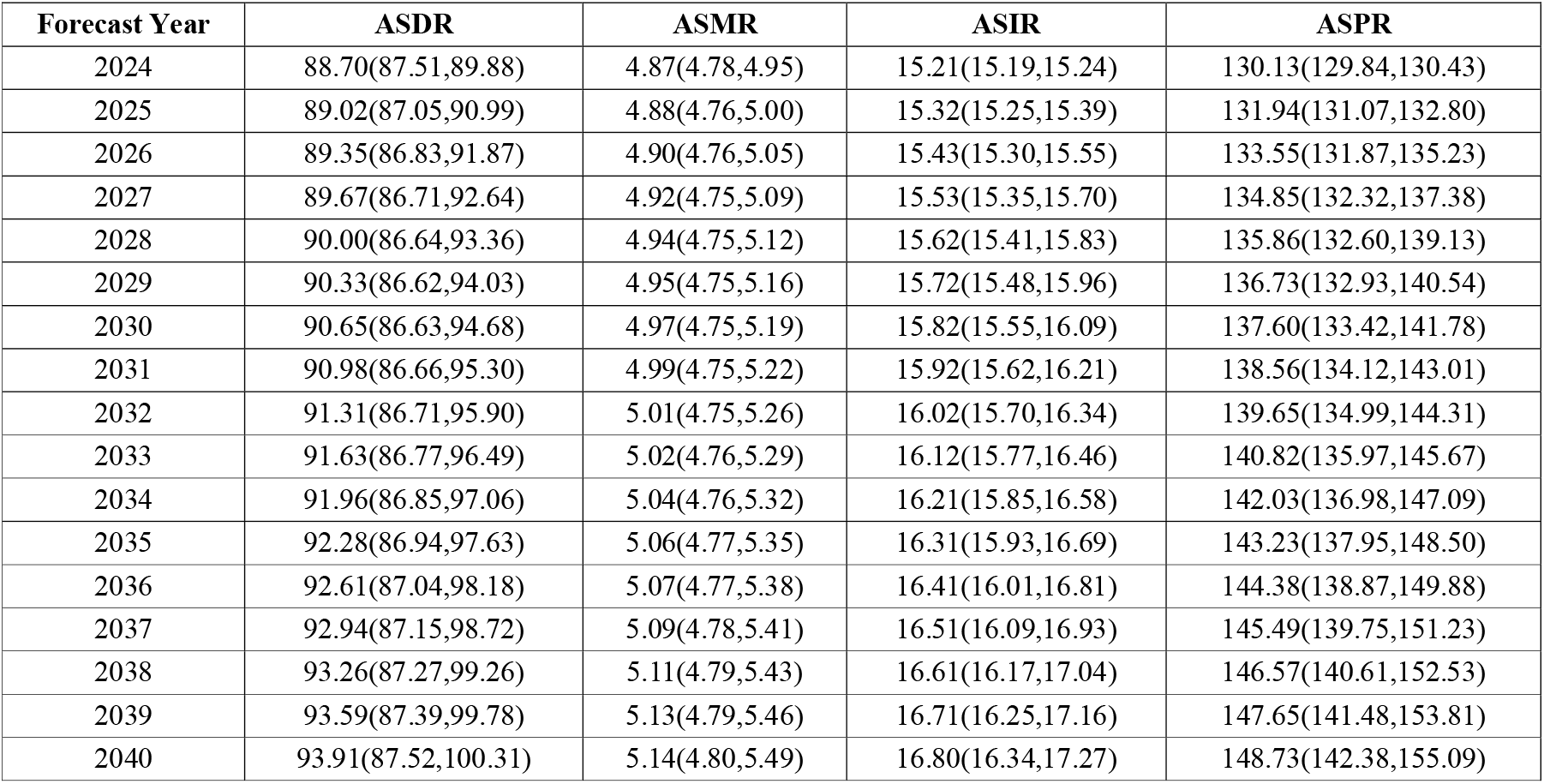
Global and regional prevalence data related to Parkinson disease. It includes location-based total numbers, age-standardized rates (ASR) per 100,000 for 1990 and 2023, percentage changes, and EAPC from 1990 to 2023.

**Figure 1.**
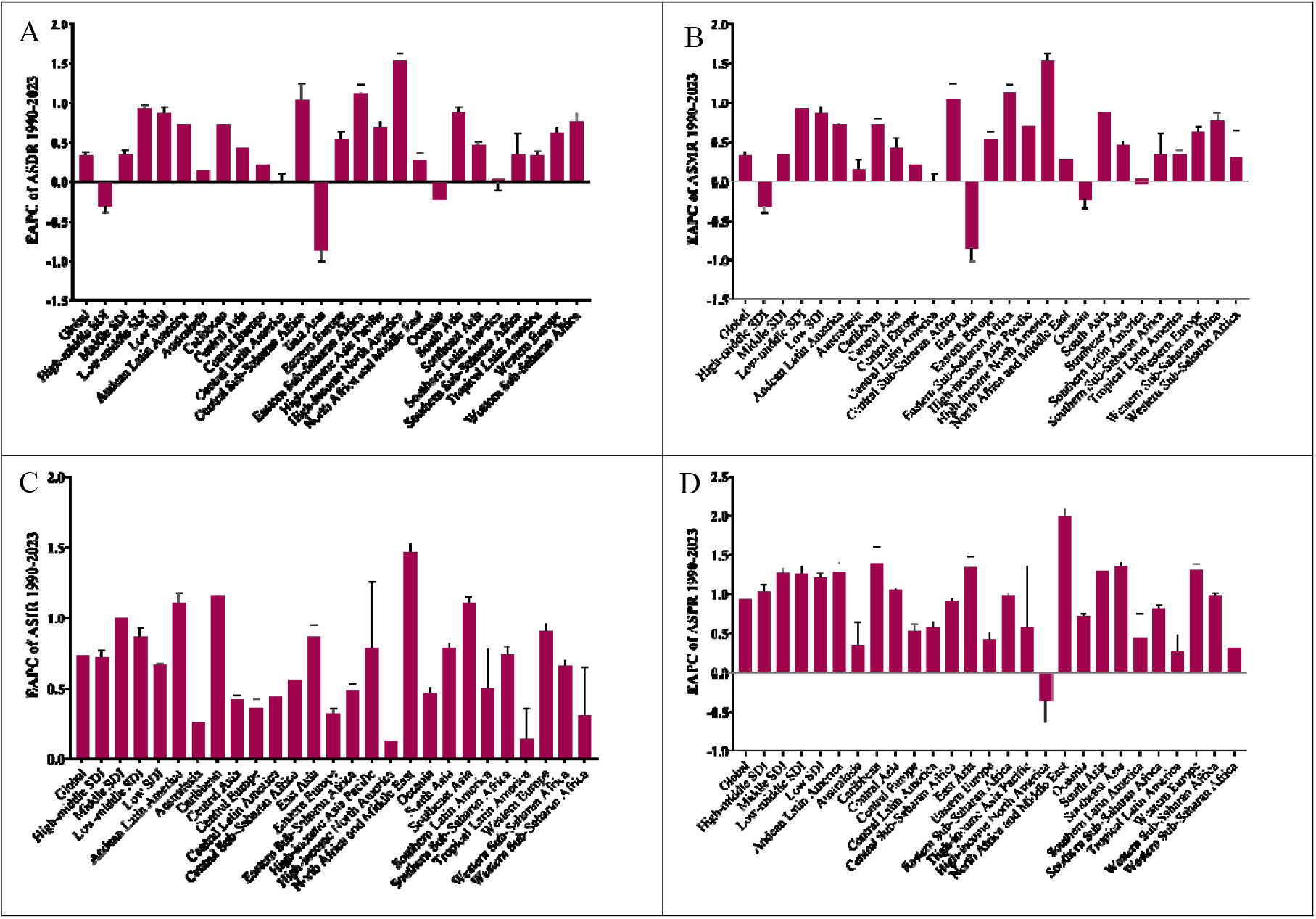
EAPC trends of Global, SDI and GBD Regions Age-Standardized Rate of Parkinson DALYs (A), Deaths (B), Incidence (C), and Prevalence (D) from 1990–2023.

### SDI-Based Trends

The low-middle SDI regions showed the largest increases (33.23%) in ASDR with the highest EAPC (0.93%). In contrast, high-middle SDI regions showed a slight decline (-3.24%) (Table 1; Fig 1A). Mortality rate increased substantially in low-middle and Low SDI regions (34.10-39.49% increases; EAPCs 0.93-0.87%), while high-middle SDI regions showed a slight decline (EAPC -0.31%) (Table 2; Fig 1B). The ASIR increase was highest in middle SDI (38.66%) and low-middle SDI countries (33.79%), with corresponding EAPCs of 1.00 and 0.87, respectively. However, high-middle SDI countries also saw notable increases (23.69%) (Table 3; Fig 1C). The prevalence also rose consistently across all SDI groups, with middle, low-middle, and low SDI regions showing the most pronounced increases (PC= 52.70, 5.24 and 47.95%; EAPCs = 1.27, 1.26, and 1.22%, respectively). In contrast, high-middle SDI areas experienced moderate growth (35.93%) (Table 4; Fig 1D).

The PD burden indicators from 1990-2023 across 21 GBD regions for their SDI index were evaluated using LOWESS regression method. It was observed that as SDI rises to middle levels, there was a marked increase in all metrics, including ASDR (Fig 2A), ASMR (Fig 2B), ASIR (Fig 2C), and ASPR (Fig 2D) of all regions. In high-SDI regions, including Western Europe, North America, and Australasia, these indicators reach their highest levels but tend to plateau or show mild fluctuations.

**Figure 2.**
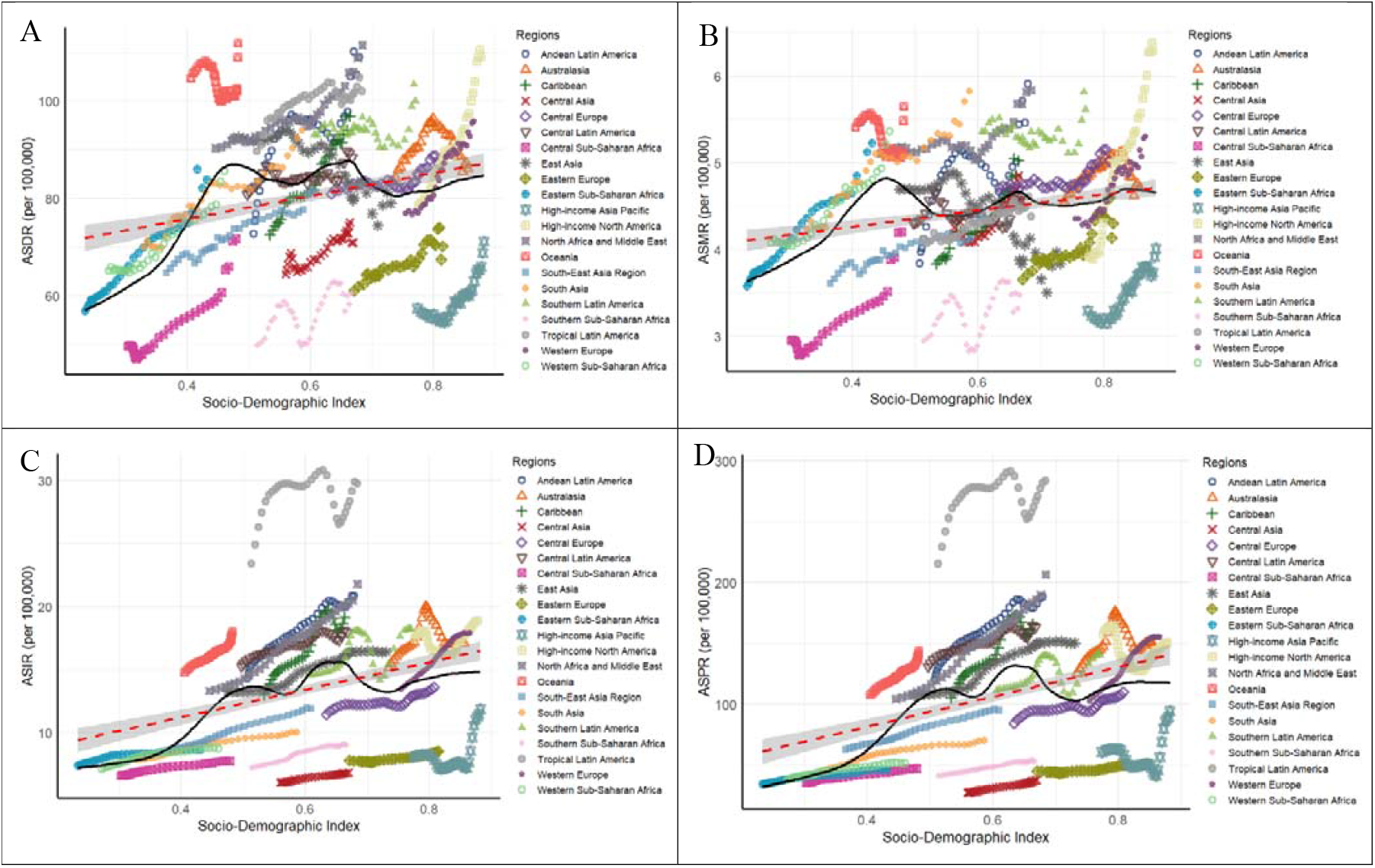
LOWESS regression of SDI vs Parkinson DALYs (A), Deaths (B), Incidence (C), and Prevalence (D) rates in GBD regions (1990-2023).

### Regional Patterns

At the regional level, the ASDR trends in High-income North America (EAPC= 1.54%) and Central Sub-Saharan Africa (EAPC= 1.04%) showed the steepest increases. The Andean Latin America, Eastern Sub-Saharan Africa, and Caribbean regions also demonstrated significant DALY growth. Conversely, East Asia, Oceania and South Latin America experienced a decline (EAPC= -0.85%, -0.22, and -0.02%) (Table 1; Fig 1A). Marked increases in ASMR were observed in High-income North America (EAPC: 1.54%), and Eastern Sub-Saharan Africa (EAPC: 1.13%). Central and Western Sub-Saharan Africa also had large percentage changes (>40%). In contrast, East Asia with the highest decreasing mortality trend (-18.27%, EAPC: -0.85%) (Table 2; Fig 1B).

Regionally, the ASIR in North Africa and the Middle East exhibited the largest surge (63.63%; EAPC= 1.47%), followed by Caribbean, Southeast Asia, and High-income Asia Pacific (EAPC= 1.16, 1.11 and 0.79 %). High-income North America had the smallest change (7.71%) (Table 3; Fig 1C). Lastly, the ASPR in North Africa and Middle East region recorded the largest growth (PC=97.74% and EAPC=2.00), followed by Caribbean (PC=57.87% and EAPC=1.39), South Asia (PC=54.08% and EAPC=1.30), and Southeast Asia (PC=51.83% and EAPC=1.35%). Western Europe also exhibited substantial increases (51%). In contrast, High-income North America showed a minor decline (-4.82%) (Table 4; Fig 1D).

### Country Specific Trends

The ASDR of PD, declined most in Kuwait (-48.7%), Qatar (-34.6%), and the Republic of Moldova (-29.0%), while it rose sharply in Maldives (+122.6%), Ethiopia (+117.0%), and Equatorial Guinea (+90.1%) (Supplementary Table S1; Fig 3A). The ASR of mortality also decreased in Kuwait (-53.7%), Qatar (-38.9%), and Moldova (-38.1%), whereas substantial increases occurred in Maldives (+166.9%), Ethiopia (+119.9%), and Equatorial Guinea (+85.9%) (Supplementary Table S2; Fig 3B). However, Incidence rates fell in the Republic of Moldova (-16.8%), Mongolia (-10.2%), and Ukraine (-7.8%), but increased sharply in Turkiye (+123.6%), Norway (+115.3%), and Germany (+96.5%) (Supplementary Table S3; Fig 3C). Prevalence decreased in Ukraine (-29.5%), the United States (-7.9%), and the Netherlands (-4.1%), while the highest rises were noted in Norway (+270.6%), Turkiye (+191.8%), and Germany (+145.5%) (Supplementary Table S4; Fig 3D).

**Figure 3.**
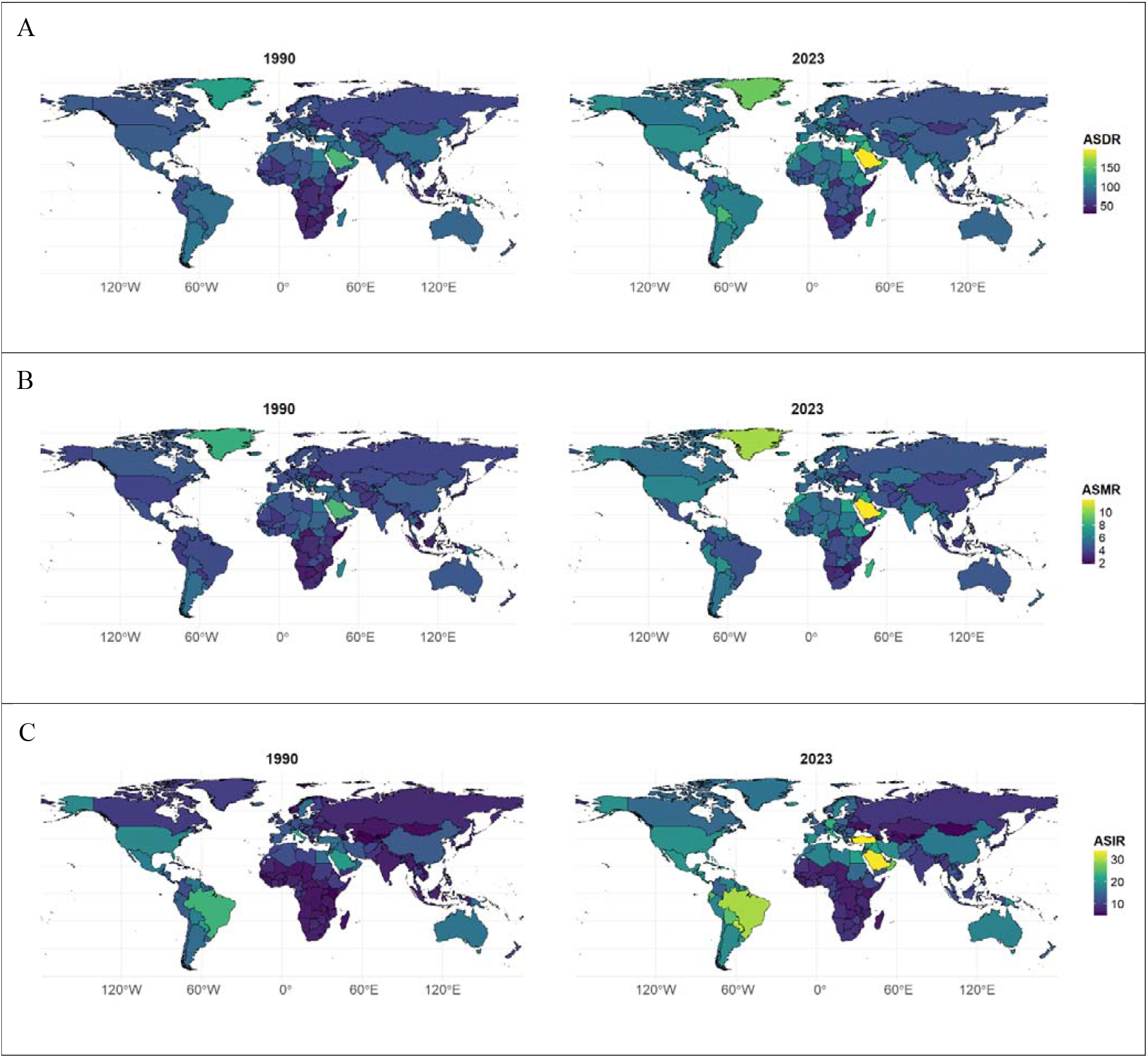
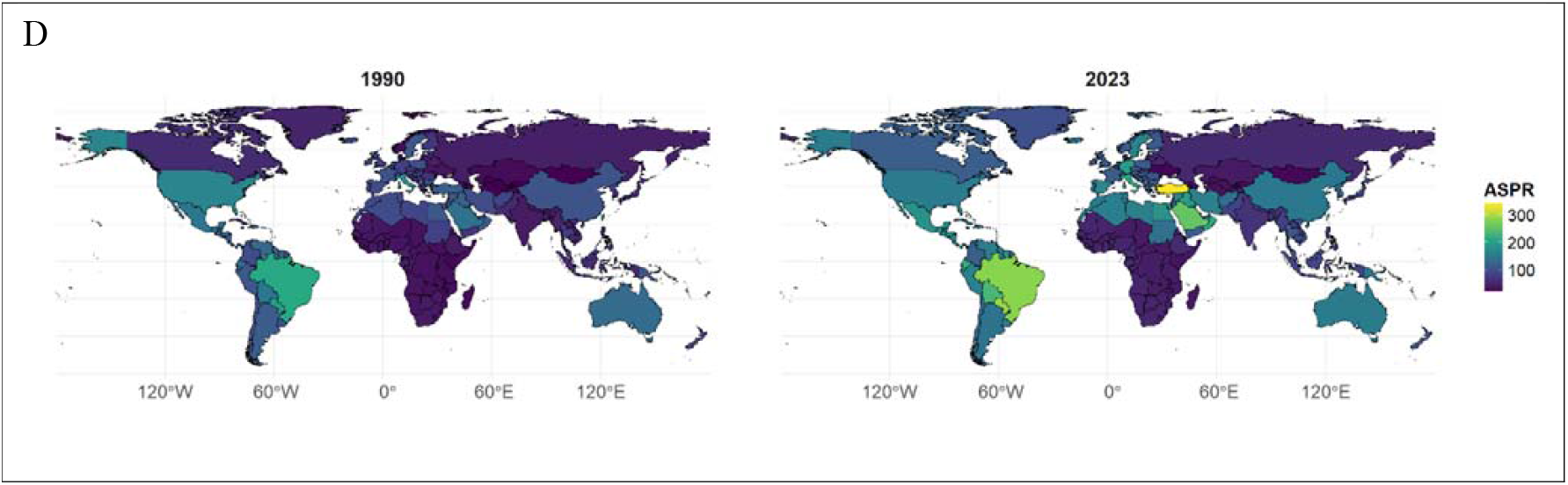
Global distribution of Age-Standardized Rates (ASR) of Parkinson DALYs (A), Deaths (B), Incidence (C), and Prevalence (D) of 1990 vs. 2023.

### Age and Sex Patterns

The global data show that the DALYs of PD, is negligible at younger ages and rises sharply after 40 years, increasing exponentially with age. The highest burden occurs between 80 and 94 years, followed by a slight decline among those aged 95 and above. Across all age groups, males consistently exhibit higher DALY rates than females, indicating a greater disease burden among men (Fig 4A). The ASMR also increased progressively with age, from 0.0027 per 100,000 males and 0.0019 females at ages 20-24 to 273.6 and 150.4 at ages 90-94. Males exhibited consistently higher mortality than females, reflecting a persistent male predominance in fatal disease burden (Fig 4B).

**Figure 4.**
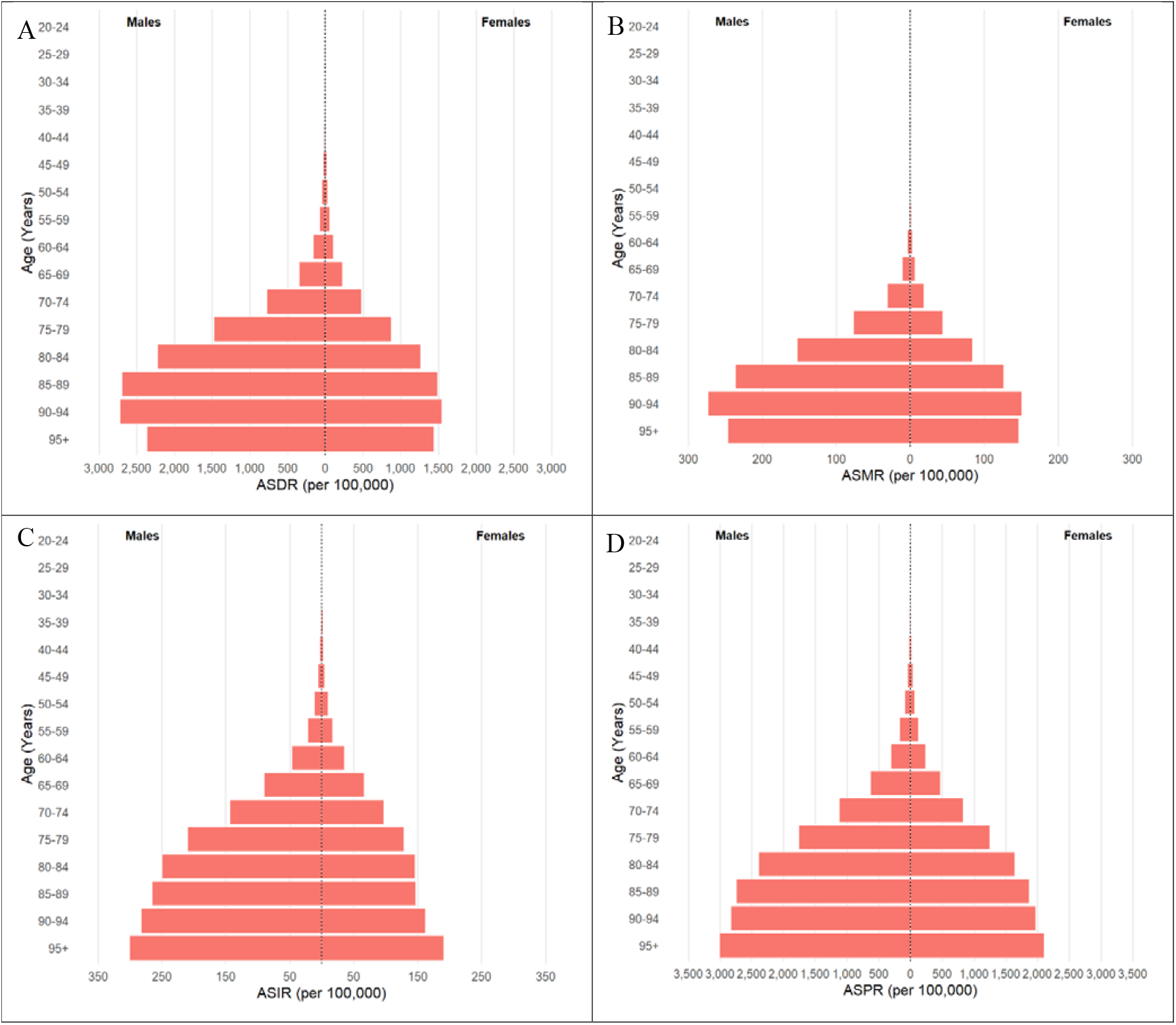
Global Gender-and age groups-based Trends of Age-Standardized rates of Parkinson DALYs (A), Deaths (B), Incidence (C), and Prevalence (D) (2023).

The incidence rate rose steadily with advancing age, increasing from 0.16 per 100,000 males and 0.10 females at ages 20-24 to 282.3 and 161.5 at ages 90-94. Minimal incidence was observed below 40 years, followed by a sharp increase beyond 50 years. Males showed higher incidence across all age groups, typically 1.40-1.70 times greater than females (Fig 4C). The ASPR also rose sharply with age, from 0.27 per 100,000 males and 0.18 females at ages 20-24 to 3006.4 and 2102.8 at ages 95 and above and increased exponentially after 60 years, with males consistently exhibiting higher prevalence than females (Fig 4D).

### Forecast to 2040

The ARIMA forecast for the global burden of Parkinson, between 2024 and 2040, the burden is projected to rise across all key age-standardized indicators. The ASDR is expected to increase from 88.70 to 93.90 (Table 4; Fig 5A), while the ASMR rises slightly from 4.87 to 5.14, suggesting a continued increase in Parkinson’s-related deaths (Table 4; Fig 5B). The ASIR shows a moderate rise from 15.21 to 16.80, indicating a growing number of new cases worldwide (Table 4; Fig 5C). The most pronounced increase is seen in the ASPR, which climbs from 130.13 to 148.73 (Table 4; Fig 5D).

**Table 4:**
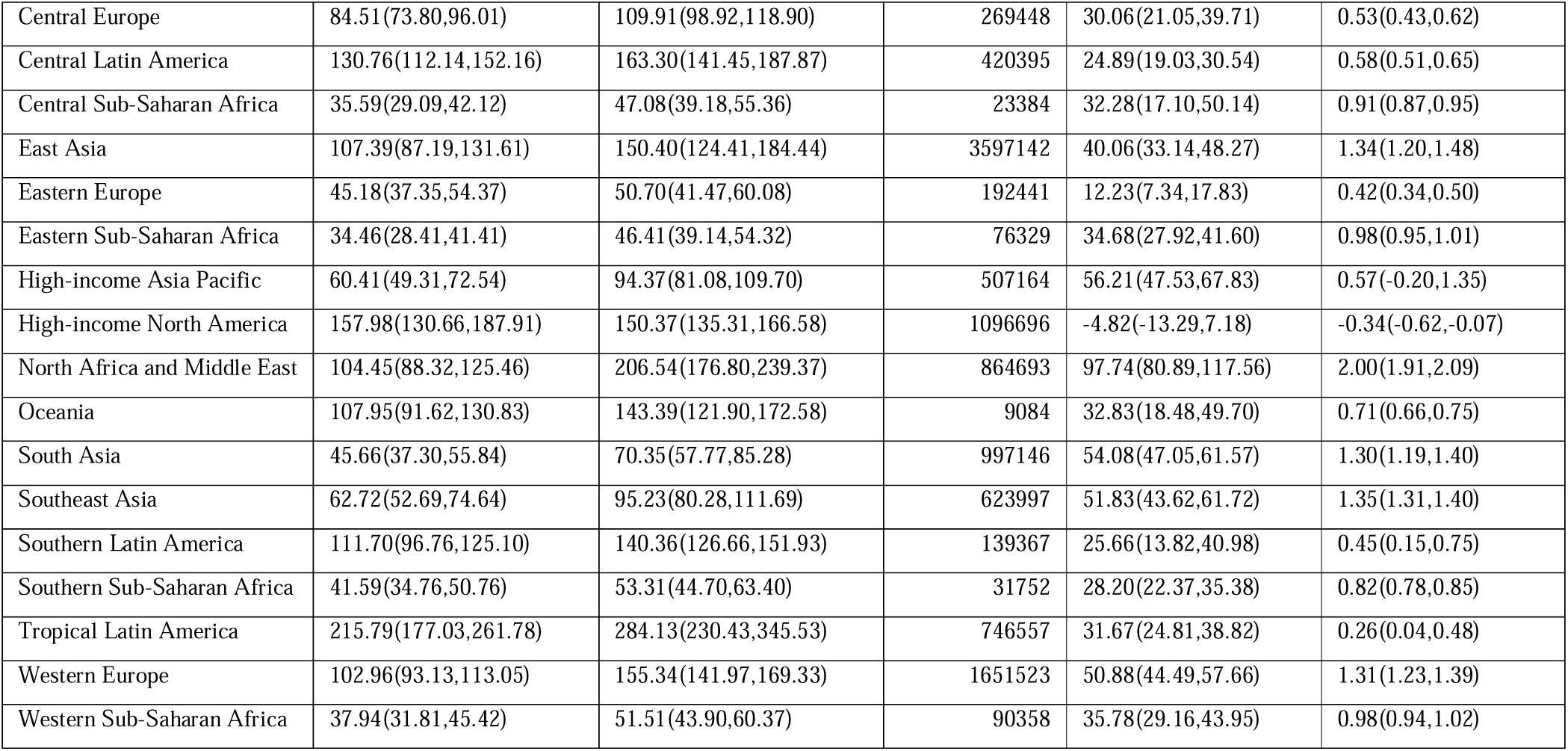
Global forecast data related to Parkinson disease from 2024 to 2040, produced through ARIMA forecasting model.

**Figure 5.**
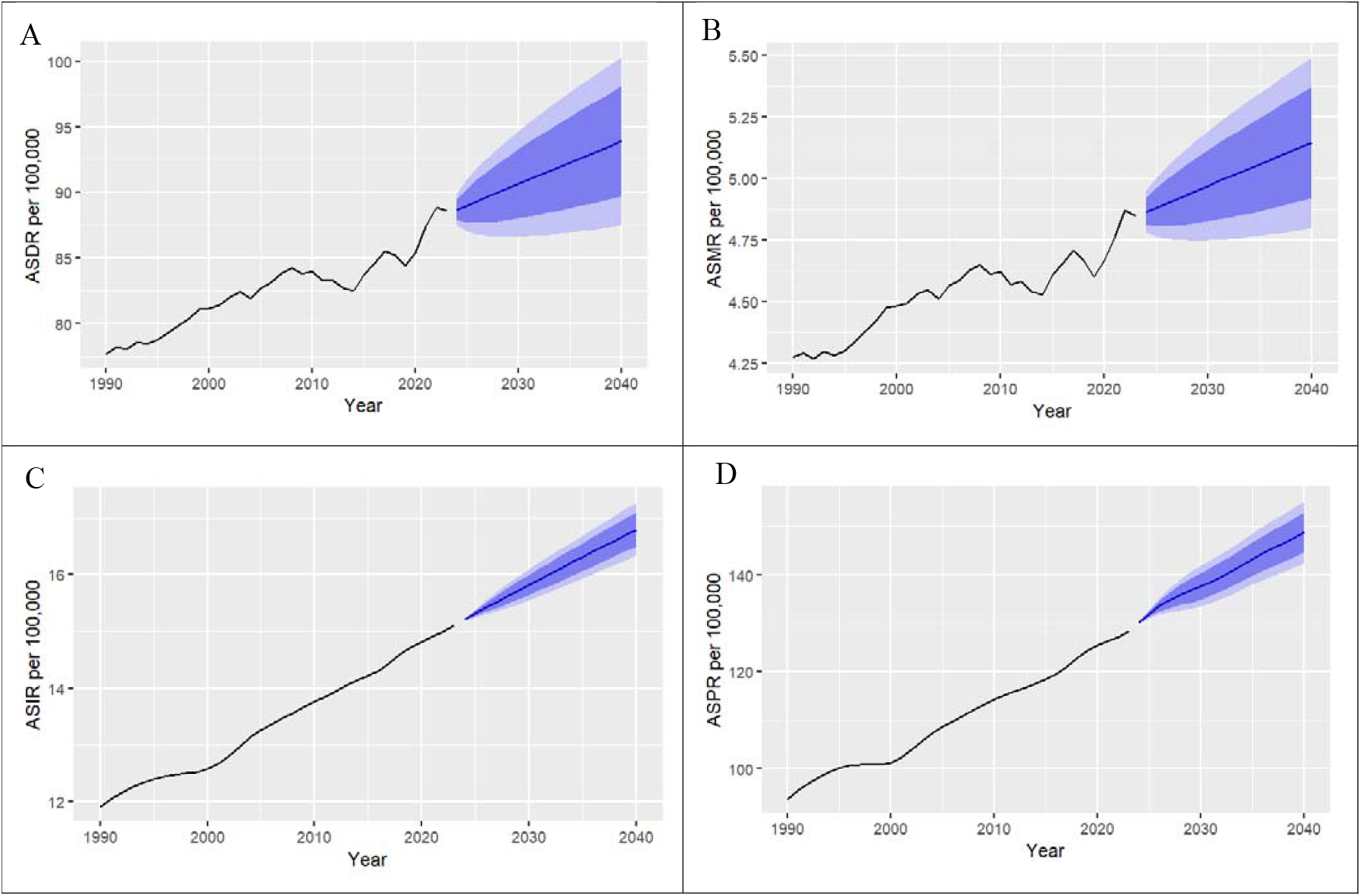
ARIMA forecasts of global Parkinson DALYs (A), Deaths (B), Incidence (C), and Prevalence (D) rates (2024–2040).

## DISCUSSION

The present study provides comprehensive insight and progressive growth of the PD at global, regional, and national levels from 1990 to 2023, along with the projections through 2040. According to the analysis of the GBD Study data from 1990-2023, there has been a consistent and perpetual rise in disability, mortality, incidence and prevalence due to PD. The increasing prevalence of PD across the world calls for urgent targeted assessments and resource planning.

The incidence rate has increased by 26.77%, supporting a true increase in disease occurrence, consistent with trends reported in the Global Burden of Disease Study 2021 [13]. One of the most prominent and highlighted the future analytical factors, is the continued prevalence of PD. There is a 37.01% rise in prevalence mainly linked with the longevity of PD patients, environmental triggers [14], lifestyle [15] and demographic changes [16], aligned with a forecast of global PD prevalence through 2050. Both the aging and improved disease management systems have contributed to clinical and epidemiological factors in the overall uplift of PD. These factors have made PD a very significant neurological worldwide [17].

Socio-Demographic Index (SDI) levels, have a sharp increase in DALYs and death rates in poor and low-middle SDI regions. The leading factors are longer lifespans, growing diagnostic facilities, and limited access to the specialized health care centers, increasing global burden of PD [18]. These findings are comparable with a steady or slight decrease in high-middle and high SDI locations due an efficient access to specialized health centers [19]. Interestingly, incidence and prevalence have the same rate in all SDI regions, highlighting the need for specialized healthcare policies to address the diagnostic and treatment gaps in lower SDI regions [20, 21].

There is an obvious difference in the prevalence of various GBD regions globally. The most rapid increase was seen in parts of Africa, the Middle East, and certain high-income regions, followed by East Asia and Oceania. In North America alone, the occurrence and global burden had been recorded (1 million and 51.9 billion), and projected to be about (1.6 million and $79 billion) by 2037. Aging, exposure to toxins, and genetics are prominent factors [22]. In contrast, poor and low-middle SDI regions like Sub-Saharan Africa face a rapid increase due to a lack of regular PD treatment facilities, professional medical staff, and the unavailability of PD drugs, along with aging and environmental factors [23-25]. According to GBD 2021 Collaborators (2025), one of the major causes of PD incidence and prevalence in North Africa and the Middle East, is the genetic basis (cousin marriages) combined with enhanced diagnostic recognition and demographic changes [16, 26].

On the other hand, in East Asia, Oceania, and Southern Latin America a sharp decline has been observed with the key improvements in healthcare delivery, timely diagnosis, and management of co-existing conditions [27]. Some of the health centers are: in 2015, China’s Center for Disease Control and Prevention (CDC) launched the nationwide Prevention and Intervention on Neurodegenerative Disease for the Elderly in China (PINDEC) program. Utilization of FDA approved DBS (deep-brain stimulation) treatment for PD in Australia [28, 29]. The healthcare systems must be strengthened as the most prioritized factor for a widened gap in PD burden across various SDI regions [30].

On national level, recent data has shown a marked decline in Kuwait, Qatar, and Moldova, in DALYs and mortality rates. Efficient healthcare systems, immediate access to specialists, and early detection are the key contributors [31]. These findings are in contrast with a rapid increase in Maldives, Ethiopia, and Equatorial Guinea with worse conditions of healthcare systems [32]. An obvious difference in incidence of cases more highlights this nationwide divergence. Moldova and Ukraine report decreasing incidence, contrary to Turkey, Germany, and Norway have shown a sharp increase, likely due to effective monitoring, aging, and greater public awareness [33]. Norway and Turkey, despite being the high-income countries are facing an increase due to lack of long-term care systems, as patients with PD are living long [30].

The global burden of PD is closely linked with aging, particularly beyond the sixth decade of life [32]. Sex difference further increases this link with the advancement of age, reflecting growing demographic and lifestyle changes as key factors [34]. Genetics, hormonal differences, and differential exposure to environmental risk factors make men consistently showing higher rates of PD than women [8, 35].

Forecasting models predict that the global burden of PD will continue to increase till 2040, due to a continuous rise in the incidence, mortality, and DALYs, suggesting that the disease will remain a growing public health challenge even with continued medical advancement. Aging and healthcare system evolution will make developing and low-middle income countries [17, 36]. As per the future analysis, presence of an adequate, advanced, and long-term healthcare systems will mitigate the challenge of the persistent rise in PD cases.

This study has several limitations that should be acknowledged. First, the analysis relied on GBD data, which may include inconsistencies due to variations in diagnostic accuracy, data completeness, and reporting quality across countries and time periods. Secondly, the aggregation of data at regional and national levels may mask within-country disparities and local trends, and cannot assess individual-level risk factors. Additionally, the projections to 2040 are modeled on current epidemiological patterns, which may change with advances in medical treatment, demographic shifts, or emerging risk factors.

Research partner-ships that integrate basic, applied and epidemiological research at the international level are needed to advance global understanding of PD and its prevention. Preventing Parkinson disease entails carefully outlining an effective strategy. Primary prevention on a population-wide basis to prevent PD from developing. Secondary prevention to limit disease progression and damage in high-risk groups and populations. Tertiary prevention following the analyses of disease-modifying treatments [32]. Future population-level estimates of PD burden are essential for guiding these strategies. A preprint of this work has previously been published (Maqsood et al., 2025) [37].

## Conclusion

In spite of advancements in diagnosis and management, the growing public health challenge of PD is highlighted by the increasing age-standardized rates of incidence, prevalence and mortality. Most of the increase in DALYs occurred in low and middle-SDI regions. This reflects the inequity in prevention, access to care and control. The burden remains substantially higher among males. The projections to 2040 indicate a continued global rise. We must conduct research, prevention efforts and prepare health systems in an international manner, the findings point out.

## Statements and Declarations

### Publication statmnet

A preprint of this work has previously been published (Maqsood et al., 2025). [37].

### CRediT authorship contribution statement

Kaleem Maqsood and Mahnoor Fatima: Data curation, Writing – original draft, Validation. Humera Naveed and Wanhar Afzal: Writing – original draft, Validation. Zulfiar Ali Beg: Writing – review & editing. Fawaz Al-hussain: Writing – review & editing, Turki Abualait: Writing – review & editing, Shahid Bashir: Conceptualization, Supervision, Validation.

## Acknowledgement

The authors have no acknowledgments to report.

## Ethics approval statement

Ethical approval was not required for this study as it involved the analysis of previously collected, anonymized data and did not involve any direct patient intervention.

## Data availability

The datasets presented in this study are accessible through an online database. You can locate them in the following database: (https://vizhub.healthdata.org/gbd-results/).

## Clinical trail

NA

