## Supplementary Table for "Global and Regional Patterns of Parkinson’s Disease: Temporal Trends, Inequality Gaps, and Projections to 2040" (from another version of this preprint: version 1, 2025-10-23)

**Table S1:** DALYs data for countries and territories related to Parkinson disease. It includes location-based total numbers, age-standardized rates (ASR) per 100,000 for 1990 and 2023, percentage changes, and EAPC from 1990 to 2023.

| **Regions** | **ASDR, 1990 (95% CI)** | **ASDR, 2023 (95% CI)** | **Numbers, 2023** | **PC 1990-2023** | **EAPC, %, 1990-2023** |
| --- | --- | --- | --- | --- | --- |
| Afghanistan | 62.30(49.32,74.14) | 74.23(60.75,87.56) | 6342 | 19.15(-1.35,45.29) | 0.53(0.42,0.64) |
| Albania | 87.57(75.15,100.93) | 98.73(81.11,117.09) | 4443 | 12.73(-5.61,36.00) | 0.33(0.23,0.42) |
| Algeria | 80.39(64.52,97.99) | 107.50(83.65,136.39) | 36373 | 33.73(8.16,65.82) | 1.05(0.95,1.15) |
| American Samoa | 122.26(102.22,143.88) | 118.59(98.03,139.93) | 46 | -3.00(-21.35,17.07) | -0.38(-0.50,-0.26) |
| Andorra | 72.91(59.93,87.14) | 76.90(64.07,89.77) | 139 | 5.48(-13.88,31.69) | 0.10(0.05,0.15) |
| Angola | 48.52(37.90,60.78) | 66.51(53.55,79.64) | 6752 | 37.07(11.52,71.74) | 1.14(1.00,1.27) |
| Antigua and Barbuda | 94.93(86.26,104.01) | 91.64(81.84,103.29) | 101 | -3.47(-12.34,6.91) | -0.09(-0.22,0.04) |
| Argentina | 94.84(87.89,101.45) | 102.10(92.62,110.23) | 66947 | 7.66(2.24,14.73) | 0.01(-0.12,0.13) |
| Armenia | 56.47(50.73,62.81) | 45.86(41.18,50.12) | 2416 | -18.79(-29.55,-9.00) | -0.31(-0.57,-0.06) |
| Australia | 85.90(78.80,93.11) | 85.49(76.63,95.09) | 47423 | -0.47(-6.85,6.51) | -0.02(-0.20,0.15) |
| Austria | 76.69(70.42,81.34) | 90.77(82.03,98.66) | 20049 | 18.37(11.95,25.74) | 0.60(0.47,0.73) |
| Azerbaijan | 63.96(52.08,79.54) | 66.21(52.80,80.96) | 5593 | 3.53(-16.13,32.64) | 0.75(0.54,0.97) |
| Bahrain | 113.54(99.17,131.31) | 109.71(92.37,129.77) | 662 | -3.37(-19.19,18.69) | 0.01(-0.18,0.19) |
| Bangladesh | 75.77(60.23,94.38) | 120.64(90.67,152.01) | 121701 | 59.21(26.01,109.97) | 0.98(0.83,1.12) |
| Barbados | 81.98(75.37,91.09) | 103.55(92.96,113.92) | 586 | 26.31(16.40,36.65) | 0.46(0.31,0.60) |
| Belarus | 58.56(52.59,65.24) | 80.32(72.61,88.04) | 13724 | 37.16(20.86,54.06) | 0.92(0.79,1.04) |
| Belgium | 76.68(69.19,81.64) | 86.05(76.43,94.07) | 23876 | 12.21(6.39,18.85) | 0.51(0.45,0.58) |
| Belize | 76.06(68.19,84.30) | 103.44(92.56,116.73) | 272 | 36.00(24.32,49.82) | 0.81(0.48,1.14) |
| Benin | 59.84(47.93,72.62) | 64.50(51.32,78.06) | 3133 | 7.80(-13.22,35.78) | 0.09(-0.02,0.20) |
| Bermuda | 84.35(75.66,93.88) | 126.46(106.38,146.54) | 192 | 49.92(30.12,69.95) | 0.71(0.53,0.89) |
| Bhutan | 72.73(54.73,89.72) | 89.48(71.78,107.77) | 541 | 23.02(-0.73,60.38) | 0.43(0.32,0.53) |
| Bolivarian Republic of Venezuela | 82.33(75.05,89.94) | 102.06(91.54,114.56) | 27334 | 23.96(13.67,36.16) | 0.59(0.45,0.72) |
| Bosnia and Herzegovina | 92.19(80.99,102.68) | 90.89(78.71,104.91) | 5926 | -1.40(-14.01,11.71) | 0.19(0.10,0.28) |
| Botswana | 42.07(34.20,49.58) | 54.95(46.30,63.90) | 867 | 30.61(9.68,59.83) | 0.25(-0.08,0.58) |
| Brazil | 90.09(79.46,101.97) | 101.76(87.18,115.89) | 260658 | 12.95(8.96,17.57) | 0.25(0.17,0.33) |
| Brunei Darussalam | 74.19(55.11,95.37) | 71.58(57.83,91.22) | 238 | -3.52(-24.41,24.64) | -0.06(-0.15,0.03) |
| Bulgaria | 90.04(83.70,95.71) | 87.28(80.35,94.77) | 14757 | -3.06(-8.98,3.62) | -0.18(-0.26,-0.09) |
| Burkina Faso | 71.19(52.57,89.21) | 76.62(57.03,94.73) | 6543 | 7.64(-12.23,34.39) | 0.06(-0.13,0.26) |
| Burundi | 45.43(34.18,59.17) | 73.63(52.83,93.55) | 2723 | 62.08(28.53,110.14) | 1.84(1.65,2.03) |
| Cambodia | 44.28(36.82,53.03) | 62.72(51.98,73.85) | 7595 | 41.66(17.02,72.94) | 1.13(1.05,1.22) |
| Cameroon | 93.62(77.93,111.10) | 109.56(90.74,131.59) | 10694 | 17.03(-4.29,44.73) | 0.64(0.49,0.80) |
| Canada | 75.38(69.51,79.41) | 94.55(85.29,103.06) | 80601 | 25.43(18.85,33.02) | 0.81(0.76,0.86) |
| Central African Republic | 42.69(31.81,54.82) | 50.55(36.76,63.62) | 984 | 18.41(-4.19,51.40) | 0.07(-0.12,0.27) |
| Chad | 78.24(58.31,97.18) | 86.19(66.43,108.31) | 3916 | 10.16(-11.61,36.64) | 0.23(0.07,0.39) |
| Chile | 89.33(83.12,95.65) | 95.60(85.12,104.06) | 26802 | 7.02(0.70,12.96) | 0.17(0.05,0.29) |
| China | 92.40(80.59,108.17) | 77.87(67.44,91.16) | 1777986 | -15.72(-29.44,0.06) | -0.67(-0.79,-0.55) |
| Colombia | 71.31(64.99,77.43) | 78.01(70.62,84.24) | 45575 | 9.40(3.35,15.91) | 0.08(-0.03,0.20) |
| Commonwealth of the Bahamas | 87.92(81.62,95.83) | 121.99(110.95,132.64) | 454 | 38.74(27.65,52.18) | 1.12(0.98,1.27) |
| Comoros | 33.31(25.56,41.52) | 41.06(33.94,49.19) | 251 | 23.28(0.15,52.42) | 0.67(0.51,0.82) |
| Congo | 46.98(38.79,57.92) | 78.78(64.13,95.70) | 1595 | 67.70(36.58,110.41) | 1.60(1.47,1.73) |
| Cook Islands | 114.55(95.59,137.35) | 154.85(130.28,184.62) | 30 | 35.18(11.61,61.84) | 0.57(0.45,0.69) |
| Costa Rica | 72.10(65.47,79.10) | 92.16(82.94,103.07) | 5429 | 27.82(18.36,37.58) | 0.48(0.37,0.60) |
| Croatia | 80.92(75.40,86.06) | 87.24(79.25,95.40) | 8736 | 7.81(0.98,14.97) | 0.36(0.31,0.40) |
| Cuba | 62.19(57.82,66.25) | 86.05(76.59,96.16) | 18009 | 38.37(25.20,52.15) | 1.13(1.00,1.26) |
| Cyprus | 86.56(69.01,106.18) | 94.12(79.95,110.68) | 1897 | 8.73(-13.69,37.65) | 0.30(0.21,0.39) |
| Czech Republic | 76.02(71.40,81.34) | 81.22(74.15,88.00) | 20401 | 6.84(0.29,13.46) | 0.46(0.38,0.53) |
| Democratic People's Republic of Korea | 81.39(67.49,97.77) | 104.39(86.61,128.17) | 31931 | 28.26(4.02,58.11) | 0.74(0.65,0.84) |
| Democratic Republic of the Congo | 50.57(38.13,62.56) | 73.44(52.31,91.90) | 22400 | 45.24(12.83,85.10) | 1.06(0.82,1.30) |
| Denmark | 58.52(54.27,62.61) | 85.58(76.20,92.89) | 12451 | 46.24(37.52,54.75) | 1.19(1.13,1.26) |
| Djibouti | 53.24(42.09,65.47) | 67.03(54.86,80.79) | 352 | 25.91(5.16,55.93) | 0.74(0.60,0.87) |
| Dominica | 89.25(73.63,104.49) | 107.20(90.28,125.57) | 115 | 20.11(0.58,45.21) | 0.29(0.20,0.39) |
| Dominican Republic | 89.97(78.01,101.44) | 131.69(111.90,152.72) | 12109 | 46.37(28.14,71.22) | 1.16(0.94,1.38) |
| Ecuador | 78.10(71.71,85.47) | 95.69(85.72,107.54) | 16966 | 22.53(13.84,32.33) | 0.15(-0.24,0.53) |
| Egypt | 103.46(87.88,118.07) | 133.15(110.40,163.60) | 63062 | 28.69(8.26,54.85) | 0.39(0.20,0.58) |
| El Salvador | 83.93(73.97,97.00) | 124.00(109.31,141.51) | 7606 | 47.74(30.71,65.73) | 1.08(0.96,1.20) |
| Equatorial Guinea | 53.38(41.22,67.65) | 101.45(86.52,121.37) | 420 | 90.05(49.39,147.36) | 2.11(1.92,2.30) |
| Eritrea | 49.63(38.57,64.70) | 73.30(57.62,87.73) | 2166 | 47.68(20.92,86.78) | 0.99(0.86,1.12) |
| Estonia | 77.52(71.88,83.37) | 83.13(74.81,90.52) | 2730 | 7.23(0.33,15.83) | -0.07(-0.18,0.04) |
| Ethiopia | 54.95(41.01,73.83) | 119.26(88.77,152.03) | 47321 | 117.05(62.04,186.77) | 2.38(2.28,2.47) |
| Federated States of Micronesia | 138.34(117.57,163.69) | 125.72(108.76,147.79) | 71 | -9.13(-26.03,10.86) | -0.37(-0.43,-0.30) |
| Fiji | 84.99(71.22,99.30) | 109.08(92.54,128.55) | 703 | 28.35(4.61,55.61) | 0.57(0.50,0.65) |
| Finland | 72.93(67.20,77.95) | 99.42(87.13,108.35) | 15405 | 36.32(27.45,44.39) | 0.93(0.89,0.98) |
| France | 74.40(68.08,79.82) | 87.08(77.77,94.29) | 150671 | 17.05(11.57,22.60) | 0.41(0.36,0.47) |
| Gabon | 53.12(43.30,67.71) | 82.17(68.30,100.24) | 879 | 54.67(27.91,86.80) | 1.13(1.02,1.23) |
| Georgia | 75.35(67.52,85.32) | 73.36(65.34,80.48) | 4729 | -2.64(-20.56,10.55) | 0.20(0.04,0.37) |
| Germany | 68.65(63.19,72.43) | 106.46(94.75,116.41) | 250301 | 55.07(47.21,63.53) | 1.10(0.91,1.29) |
| Ghana | 61.13(50.67,71.23) | 81.66(67.88,94.93) | 11821 | 33.60(10.14,65.15) | 0.68(0.49,0.87) |
| Greece | 73.06(67.08,77.84) | 92.08(83.15,101.15) | 28437 | 26.04(18.57,34.96) | 0.80(0.72,0.89) |
| Greenland | 123.39(86.85,181.38) | 155.52(121.96,201.46) | 75 | 26.04(-0.68,64.32) | 0.90(0.80,0.99) |
| Grenada | 80.84(73.32,89.14) | 83.07(73.98,94.18) | 136 | 2.76(-6.29,13.47) | 0.19(0.08,0.31) |
| Guam | 84.94(75.70,94.90) | 89.83(81.17,100.40) | 184 | 5.76(-5.51,16.66) | -0.14(-0.26,-0.02) |
| Guatemala | 85.86(79.42,92.87) | 83.35(74.54,94.39) | 8374 | -2.92(-12.15,7.88) | -0.23(-0.33,-0.13) |
| Guinea | 74.36(58.49,94.70) | 79.24(64.57,94.28) | 4161 | 6.56(-14.50,36.92) | -0.28(-0.50,-0.05) |
| Guinea-Bissau | 58.56(47.89,70.42) | 83.75(69.52,101.83) | 516 | 43.01(15.70,79.87) | 0.82(0.69,0.96) |
| Guyana | 81.51(73.78,89.96) | 111.97(98.12,126.67) | 714 | 37.37(21.11,56.36) | 0.52(0.33,0.71) |
| Haiti | 67.84(56.03,82.89) | 97.88(82.34,116.44) | 5649 | 44.28(16.39,81.10) | 0.93(0.87,0.98) |
| Honduras | 94.63(79.31,109.41) | 134.10(111.44,164.32) | 8044 | 41.71(15.88,69.22) | 0.85(0.78,0.91) |
| Hungary | 76.37(71.67,81.13) | 86.00(79.41,93.02) | 18683 | 12.62(6.99,18.87) | 0.24(0.13,0.36) |
| Iceland | 95.72(87.06,102.76) | 124.48(109.80,136.78) | 821 | 30.05(20.10,42.04) | 0.78(0.68,0.87) |
| India | 71.47(55.14,86.07) | 94.16(75.76,114.17) | 1000079 | 31.74(3.70,65.33) | 0.81(0.73,0.90) |
| Indonesia | 52.48(42.34,62.49) | 69.61(57.83,84.45) | 153438 | 32.63(5.79,64.23) | 0.87(0.77,0.97) |
| Iraq | 88.49(69.36,115.98) | 140.94(117.21,168.36) | 26426 | 59.28(17.20,109.38) | 1.40(1.33,1.47) |
| Ireland | 81.03(75.06,87.07) | 94.55(84.15,102.40) | 8816 | 16.68(8.68,24.90) | 0.40(0.30,0.49) |
| Islamic Republic of Iran | 89.30(75.14,107.22) | 86.43(72.00,102.29) | 57980 | -3.22(-24.15,22.00) | 0.05(-0.03,0.12) |
| Israel | 88.42(79.92,95.41) | 89.18(78.76,99.10) | 12516 | 0.86(-5.71,8.18) | 0.33(0.23,0.43) |
| Italy | 92.36(81.48,101.91) | 100.27(86.88,112.22) | 175347 | 8.57(3.37,14.82) | 0.39(0.21,0.58) |
| Jamaica | 69.97(62.98,77.38) | 89.62(80.30,100.01) | 2917 | 28.10(16.45,41.32) | 0.71(0.57,0.84) |
| Japan | 55.49(49.70,59.57) | 69.76(60.95,76.93) | 329625 | 25.71(18.77,33.68) | 0.57(0.43,0.71) |
| Jordan | 86.94(70.25,107.29) | 92.58(76.55,107.38) | 6242 | 6.49(-15.72,37.30) | 0.09(0.03,0.16) |
| Kazakhstan | 65.31(60.78,69.32) | 82.27(73.50,89.00) | 14020 | 25.97(15.02,34.68) | 0.74(0.57,0.91) |
| Kenya | 52.51(39.58,66.05) | 59.98(49.05,72.26) | 13056 | 14.23(-9.17,46.51) | 0.19(0.01,0.38) |
| Kingdom of Eswatini | 52.82(42.30,64.92) | 46.37(38.66,55.25) | 313 | -12.20(-29.58,12.06) | -0.72(-0.93,-0.51) |
| Kiribati | 61.40(51.67,71.85) | 70.02(60.37,83.35) | 43 | 14.03(-1.17,32.83) | 0.22(0.17,0.26) |
| Kuwait | 163.09(148.80,175.94) | 83.60(73.93,93.12) | 1455 | -48.74(-53.08,-43.89) | -1.57(-2.10,-1.04) |
| Kyrgyzstan | 59.18(53.78,64.32) | 57.43(53.09,61.96) | 2118 | -2.96(-12.05,7.80) | -0.26(-0.53,0.00) |
| Lao People's Democratic Republic | 54.09(43.27,69.75) | 93.08(78.72,109.19) | 3868 | 72.08(35.80,117.58) | 1.65(1.60,1.71) |
| Latvia | 72.78(67.06,78.06) | 75.80(69.89,81.40) | 3602 | 4.15(-3.65,14.95) | 0.06(-0.03,0.16) |
| Lebanon | 46.42(36.37,59.65) | 69.20(58.09,81.33) | 4408 | 49.05(22.49,94.68) | 1.46(1.19,1.73) |
| Lesotho | 53.26(41.71,66.46) | 53.81(44.55,64.55) | 635 | 1.04(-21.50,27.64) | -0.76(-1.14,-0.39) |
| Liberia | 77.50(63.05,93.76) | 92.00(72.83,113.09) | 1688 | 18.71(-2.88,49.11) | 0.50(0.32,0.67) |
| Libya | 78.30(65.26,94.66) | 95.93(77.63,120.31) | 4440 | 22.51(0.25,50.02) | 0.60(0.49,0.71) |
| Lithuania | 61.33(56.90,65.34) | 74.79(67.69,80.32) | 5080 | 21.96(14.60,31.04) | 0.74(0.63,0.85) |
| Luxembourg | 93.87(86.58,100.27) | 98.97(88.95,109.20) | 1144 | 5.43(-3.06,13.27) | 0.27(0.19,0.35) |
| Madagascar | 95.86(73.45,119.69) | 122.42(91.17,155.27) | 9749 | 27.71(3.67,63.73) | 0.67(0.50,0.85) |
| Malawi | 62.07(46.49,77.50) | 82.50(62.96,98.07) | 5460 | 32.90(8.73,70.59) | 0.75(0.50,0.99) |
| Malaysia | 72.08(61.45,85.64) | 85.75(74.16,100.45) | 24416 | 18.97(-3.21,40.76) | 0.25(0.13,0.38) |
| Maldives | 69.49(59.93,82.69) | 154.67(129.45,177.05) | 346 | 122.59(79.67,178.54) | 2.17(2.08,2.27) |
| Mali | 52.89(41.42,65.52) | 66.34(52.25,80.70) | 4992 | 25.44(5.23,56.56) | 0.33(0.22,0.45) |
| Malta | 90.34(82.14,97.73) | 89.67(78.69,98.94) | 1046 | -0.74(-8.84,8.42) | 0.00(-0.14,0.14) |
| Marshall Islands | 148.74(123.18,180.66) | 181.93(152.54,215.08) | 34 | 22.32(-1.05,47.88) | 0.40(0.30,0.50) |
| Mauritania | 63.46(51.66,75.19) | 100.96(78.49,124.83) | 1916 | 59.09(29.85,99.24) | 1.22(1.07,1.36) |
| Mauritius | 79.92(73.76,85.17) | 94.16(85.28,102.56) | 1647 | 17.81(10.61,25.90) | 0.17(0.06,0.27) |
| Mexico | 84.50(76.22,91.75) | 87.25(78.95,96.87) | 113305 | 3.26(-1.55,8.54) | -0.04(-0.12,0.04) |
| Mongolia | 60.79(51.87,71.33) | 55.37(46.49,65.60) | 986 | -8.93(-26.21,8.50) | -0.59(-0.72,-0.46) |
| Montenegro | 97.71(85.34,111.87) | 99.63(87.52,114.28) | 1134 | 1.96(-10.94,19.11) | 0.15(0.05,0.24) |
| Morocco | 89.03(73.97,102.98) | 128.38(103.84,158.81) | 40545 | 44.19(15.29,83.52) | 1.07(0.94,1.21) |
| Mozambique | 42.31(30.99,52.81) | 48.79(38.32,58.65) | 5326 | 15.30(-9.36,49.00) | 0.11(-0.07,0.28) |
| Myanmar | 67.66(54.30,82.58) | 99.67(84.50,120.10) | 42746 | 47.32(21.03,83.47) | 1.27(1.17,1.36) |
| Namibia | 44.19(36.45,52.13) | 50.98(43.91,60.14) | 746 | 15.35(-2.69,40.18) | 0.43(0.30,0.56) |
| Nepal | 50.67(39.13,61.58) | 69.80(57.20,85.89) | 15722 | 37.76(13.11,73.04) | 0.40(0.21,0.59) |
| Netherlands | 93.97(85.40,99.99) | 88.49(78.94,96.34) | 37425 | -5.83(-10.41,-0.18) | -0.04(-0.14,0.05) |
| New Zealand | 71.93(65.30,77.31) | 89.77(82.06,97.84) | 8659 | 24.81(18.20,31.46) | 0.64(0.58,0.70) |
| Nicaragua | 75.20(64.35,85.64) | 89.60(76.50,104.01) | 3885 | 19.14(2.35,42.38) | 0.99(0.81,1.16) |
| Niger | 68.56(50.30,88.59) | 81.78(57.68,107.67) | 4839 | 19.29(-4.02,55.70) | 0.33(0.26,0.40) |
| Nigeria | 62.91(51.76,75.12) | 90.73(72.79,111.99) | 67343 | 44.24(13.69,81.25) | 1.17(1.08,1.25) |
| Northern Mariana Islands | 113.00(94.65,139.57) | 130.99(113.06,150.96) | 42 | 15.92(-5.34,43.45) | 0.40(0.31,0.50) |
| Norway | 65.20(59.15,69.26) | 93.24(81.98,102.14) | 10953 | 43.01(32.63,53.20) | 1.45(1.33,1.58) |
| Oman | 93.89(74.92,116.66) | 136.09(111.38,163.09) | 1993 | 44.94(3.59,89.75) | 1.72(1.55,1.90) |
| Pakistan | 58.47(47.10,70.73) | 74.64(63.27,87.38) | 87069 | 27.65(4.13,54.86) | 0.75(0.64,0.86) |
| Palestine | 75.67(63.23,90.68) | 93.38(79.56,107.29) | 2017 | 23.40(0.66,52.19) | 0.50(0.45,0.55) |
| Panama | 70.61(63.82,77.72) | 94.38(83.87,104.38) | 4347 | 33.67(22.82,44.76) | 0.70(0.60,0.81) |
| Papua New Guinea | 112.27(84.93,141.51) | 113.83(86.76,139.69) | 4499 | 1.39(-17.92,24.27) | -0.31(-0.42,-0.20) |
| Paraguay | 76.92(66.17,89.72) | 119.55(99.75,137.80) | 5925 | 55.42(31.47,75.16) | 1.39(1.34,1.44) |
| Peru | 68.01(60.46,76.01) | 107.99(91.90,125.05) | 38491 | 58.79(37.73,83.98) | 0.86(0.67,1.04) |
| Philippines | 64.04(55.71,72.77) | 77.64(68.68,87.55) | 54435 | 21.24(5.45,39.43) | 0.61(0.51,0.71) |
| Plurinational State of Bolivia | 85.61(71.54,102.16) | 140.46(120.29,164.94) | 12157 | 64.07(35.45,99.08) | 1.60(1.49,1.72) |
| Poland | 81.14(75.40,87.55) | 88.41(81.31,94.35) | 71671 | 8.97(2.85,15.19) | 0.09(-0.01,0.19) |
| Portugal | 66.35(60.83,71.43) | 77.54(68.01,84.71) | 23583 | 16.86(10.29,23.95) | 0.66(0.53,0.79) |
| Principality of Monaco | 92.64(74.66,109.33) | 114.81(93.49,143.51) | 144 | 23.93(-0.65,57.96) | 0.61(0.49,0.74) |
| Puerto Rico | 79.94(73.35,87.57) | 83.22(73.66,92.02) | 8180 | 4.11(-2.29,9.83) | -0.13(-0.27,0.00) |
| Qatar | 143.62(119.85,172.94) | 93.93(78.23,116.25) | 510 | -34.60(-48.22,-16.94) | -1.82(-2.03,-1.60) |
| Republic of CÃ´te d'Ivoire | 66.49(55.10,80.37) | 78.31(65.97,93.91) | 7446 | 17.77(-1.78,47.21) | 0.22(-0.04,0.47) |
| Republic of Cabo Verde | 67.29(53.68,81.72) | 73.69(61.69,88.22) | 303 | 9.51(-12.64,35.56) | 0.26(0.07,0.46) |
| Republic of Korea | 68.98(56.77,92.84) | 78.40(66.63,88.25) | 81111 | 13.65(-18.36,41.06) | 0.71(0.60,0.82) |
| Republic of Moldova | 70.82(66.03,75.52) | 50.25(46.06,54.83) | 3197 | -29.04(-33.95,-23.02) | -1.33(-1.49,-1.17) |
| Republic of Nauru | 189.37(156.77,238.21) | 194.65(161.15,241.00) | 7 | 2.79(-18.81,28.79) | 0.06(-0.13,0.24) |
| Republic of Niue | 102.27(85.86,119.19) | 101.87(84.67,124.80) | 2 | -0.39(-18.91,19.07) | -0.20(-0.30,-0.11) |
| Republic of Palau | 100.58(85.40,118.12) | 106.80(93.56,124.30) | 19 | 6.18(-10.86,26.78) | 0.03(-0.06,0.12) |
| Republic of San Marino | 59.00(49.09,69.30) | 44.56(36.76,52.30) | 42 | -24.46(-36.28,-8.35) | -1.19(-1.32,-1.05) |
| Republic of the Gambia | 45.07(35.83,54.65) | 60.25(46.46,73.46) | 600 | 33.68(10.13,71.34) | 0.56(0.28,0.83) |
| Romania | 77.04(72.63,81.40) | 85.69(78.99,92.66) | 35293 | 11.22(4.65,17.55) | 0.41(0.33,0.49) |
| Russian Federation | 65.86(61.39,69.95) | 74.29(68.00,79.20) | 190506 | 12.79(6.74,20.18) | 0.31(0.21,0.41) |
| Rwanda | 59.63(46.30,79.68) | 99.20(78.37,122.38) | 5537 | 66.36(30.30,116.55) | 1.72(1.51,1.93) |
| Saint Kitts and Nevis | 130.82(121.58,140.06) | 141.16(129.86,154.81) | 72 | 7.90(-1.20,18.17) | 0.15(0.00,0.31) |
| Saint Lucia | 102.62(93.65,112.73) | 97.01(86.66,108.18) | 240 | -5.46(-14.68,4.84) | -0.39(-0.49,-0.29) |
| Saint Vincent and the Grenadines | 76.59(69.10,84.73) | 81.23(71.98,92.92) | 135 | 6.06(-3.94,17.44) | 0.03(-0.12,0.19) |
| Samoa | 68.21(55.75,84.37) | 85.52(71.26,104.01) | 136 | 25.38(5.15,46.75) | 0.30(0.18,0.42) |
| Sao Tome and Principe | 64.20(54.69,73.76) | 92.00(76.43,109.80) | 91 | 43.30(14.99,77.80) | 1.28(1.18,1.38) |
| Saudi Arabia | 142.04(116.43,185.28) | 196.58(167.77,233.28) | 16675 | 38.40(2.69,78.34) | 1.66(1.36,1.97) |
| Senegal | 49.20(39.19,59.95) | 74.56(60.57,88.10) | 5664 | 51.55(24.50,87.57) | 1.17(0.99,1.35) |
| Serbia | 90.47(78.27,105.53) | 95.93(84.52,108.46) | 15920 | 6.03(-9.59,23.52) | 0.09(0.04,0.13) |
| Seychelles | 114.46(95.63,133.48) | 111.99(95.26,132.82) | 128 | -2.16(-19.21,14.16) | -0.35(-0.46,-0.25) |
| Sierra Leone | 69.81(53.52,88.43) | 90.65(72.29,109.06) | 2604 | 29.86(2.58,68.65) | 0.86(0.61,1.11) |
| Singapore | 64.36(60.19,67.60) | 60.83(53.78,66.51) | 5521 | -5.49(-12.84,3.40) | -0.32(-0.40,-0.24) |
| Slovakia | 77.09(68.15,90.20) | 77.87(68.72,87.09) | 8052 | 1.02(-11.32,16.90) | 0.09(0.06,0.12) |
| Slovenia | 84.23(78.35,90.17) | 84.30(75.06,92.50) | 4451 | 0.08(-7.81,7.09) | -0.09(-0.22,0.04) |
| Socialist Republic of Viet Nam | 67.56(54.04,86.96) | 85.07(66.95,106.96) | 86199 | 25.92(2.08,59.27) | 0.77(0.66,0.87) |
| Solomon Islands | 132.32(93.42,173.99) | 140.40(106.36,170.17) | 502 | 6.11(-16.37,34.87) | 0.00(-0.25,0.25) |
| Somalia | 30.18(20.49,41.41) | 38.39(28.44,47.64) | 2869 | 27.20(-1.58,70.25) | 0.67(0.51,0.84) |
| South Africa | 50.46(42.82,57.79) | 64.81(56.51,74.01) | 29120 | 28.44(9.35,51.27) | 0.79(0.55,1.03) |
| South Sudan | 77.29(58.16,101.32) | 97.24(73.62,121.39) | 2916 | 25.80(2.27,65.14) | 0.54(0.46,0.63) |
| Spain | 79.60(72.53,85.04) | 97.16(85.41,108.68) | 116251 | 22.06(14.72,29.59) | 0.82(0.71,0.93) |
| Sri Lanka | 77.54(68.05,87.42) | 93.32(80.90,105.56) | 28429 | 20.35(4.96,41.54) | 0.36(0.28,0.45) |
| Sudan | 87.73(73.41,105.07) | 105.96(84.67,132.21) | 19965 | 20.78(-5.41,58.60) | 0.49(0.42,0.57) |
| Suriname | 70.13(61.04,81.14) | 88.67(76.24,100.88) | 542 | 26.43(9.97,45.13) | 0.63(0.54,0.73) |
| Sweden | 76.69(70.77,82.50) | 86.64(76.98,95.94) | 23106 | 12.97(5.32,20.49) | 0.34(0.18,0.51) |
| Switzerland | 72.04(65.62,77.99) | 83.47(72.57,91.84) | 18199 | 15.87(7.66,23.34) | 0.56(0.50,0.62) |
| Syrian Arab Republic | 94.05(76.10,117.99) | 116.02(94.91,141.66) | 12512 | 23.36(-9.94,67.14) | 0.37(0.21,0.54) |
| Taiwan (Province of China) | 100.79(92.73,109.21) | 112.03(101.05,124.15) | 50982 | 11.16(4.40,19.39) | 0.56(0.38,0.74) |
| Tajikistan | 87.88(72.14,108.32) | 130.49(107.35,163.12) | 4947 | 48.48(15.95,97.44) | 0.90(0.61,1.18) |
| Thailand | 87.23(72.46,103.25) | 86.71(74.09,105.28) | 101283 | -0.59(-15.00,17.90) | -0.38(-0.60,-0.15) |
| The former Yugoslav Republic of Macedonia | 92.37(83.37,103.44) | 101.75(86.53,116.90) | 3492 | 10.15(-7.23,30.42) | 0.36(0.29,0.42) |
| Timor-Leste | 69.85(54.32,85.49) | 79.39(64.35,97.46) | 654 | 13.67(-7.50,39.84) | 0.24(0.15,0.32) |
| Togo | 66.02(52.41,80.51) | 74.11(58.76,89.41) | 2389 | 12.25(-8.03,38.84) | 0.08(-0.14,0.31) |
| Tokelau | 91.02(75.74,107.73) | 91.82(76.10,112.91) | 2 | 0.88(-16.32,19.54) | -0.16(-0.28,-0.05) |
| Tonga | 88.92(72.40,108.37) | 107.83(90.93,131.14) | 85 | 21.27(0.65,46.00) | 0.35(0.25,0.44) |
| Trinidad and Tobago | 101.30(93.82,108.89) | 109.58(99.82,120.79) | 1915 | 8.18(0.24,18.50) | -0.02(-0.14,0.11) |
| Tunisia | 74.99(60.68,94.90) | 82.91(70.47,96.92) | 10229 | 10.56(-6.37,35.16) | -0.08(-0.17,0.01) |
| Turkey | 92.74(72.51,114.43) | 123.16(102.48,144.66) | 113953 | 32.80(5.78,69.28) | 0.49(0.34,0.63) |
| Turkmenistan | 55.33(50.42,60.06) | 67.27(57.51,75.36) | 2263 | 21.59(3.98,37.29) | 0.55(0.44,0.67) |
| Tuvalu | 132.88(110.47,159.91) | 167.31(138.66,199.13) | 12 | 25.91(2.87,55.61) | 0.46(0.35,0.57) |
| Uganda | 65.33(46.14,86.25) | 80.84(62.19,99.04) | 10704 | 23.74(-2.49,61.93) | 0.65(0.45,0.85) |
| Ukraine | 48.06(43.82,51.86) | 56.22(50.03,63.07) | 46331 | 16.96(2.72,33.76) | 0.81(0.67,0.95) |
| United Arab Emirates | 98.90(75.26,137.78) | 89.78(71.31,108.87) | 2107 | -9.23(-32.93,25.69) | -0.27(-0.44,-0.10) |
| United Kingdom | 77.10(71.12,81.87) | 98.19(88.98,105.45) | 153981 | 27.35(22.82,32.05) | 0.83(0.69,0.98) |
| United Republic of Tanzania | 53.08(40.89,64.78) | 67.09(51.49,81.09) | 17066 | 26.38(2.25,61.31) | 0.80(0.71,0.90) |
| United States of America | 78.96(69.11,87.58) | 110.99(99.22,120.17) | 720710 | 40.56(32.09,49.56) | 1.02(0.93,1.12) |
| United States Virgin Islands | 123.54(111.49,136.97) | 126.44(110.20,145.27) | 282 | 2.34(-9.69,16.32) | -0.38(-0.59,-0.17) |
| Uruguay | 89.29(82.76,95.94) | 100.26(90.90,109.26) | 6429 | 12.29(6.01,20.07) | 0.24(0.10,0.37) |
| Uzbekistan | 55.64(50.41,60.46) | 63.68(58.99,68.31) | 12364 | 14.45(3.06,26.97) | 0.41(0.35,0.46) |
| Vanuatu | 71.68(54.75,88.24) | 74.12(60.58,88.20) | 126 | 3.41(-15.58,25.11) | -0.21(-0.42,0.00) |
| Yemen | 70.55(56.72,83.70) | 79.17(65.29,96.92) | 9134 | 12.23(-6.35,40.23) | 0.36(0.29,0.43) |
| Zambia | 65.97(52.18,79.90) | 86.97(70.02,107.61) | 4200 | 31.83(6.80,66.97) | 0.65(0.52,0.79) |
| Zimbabwe | 47.94(39.86,55.45) | 40.76(34.05,50.91) | 3226 | -14.98(-33.25,7.28) | -1.46(-1.87,-1.04) |

**Table S2:** Deaths data for countries and territories related to Parkinson disease. It includes location-based total numbers, age-standardized rates (ASR) per 100,000 for 1990 and 2023, percentage changes, and EAPC from 1990 to 2023.

| **Regions** | **ASMR, 1990 (95% CI)** | **ASMR, 2023 (95% CI)** | **Numbers, 2023** | **PC 1990-2023** | **EAPC, %, 1990-2023** |
| --- | --- | --- | --- | --- | --- |
| Afghanistan | 3.42(2.62,4.30) | 3.89(3.00,4.90) | 268 | 13.82(-8.85,43.03) | 0.29(0.15,0.43) |
| Albania | 5.55(4.63,6.52) | 5.98(4.65,7.35) | 263 | 7.83(-13.58,34.49) | 0.12(-0.01,0.25) |
| Algeria | 4.59(3.44,5.89) | 5.67(4.09,7.56) | 1762 | 23.58(-6.85,65.12) | 0.87(0.76,0.99) |
| American Samoa | 6.50(5.33,7.99) | 6.05(4.83,7.44) | 2 | -7.00(-26.41,17.43) | -0.48(-0.60,-0.35) |
| Andorra | 4.18(3.31,5.26) | 4.42(3.55,5.28) | 9 | 5.79(-16.79,39.45) | 0.11(0.05,0.16) |
| Angola | 2.85(2.20,3.72) | 3.78(2.94,4.63) | 331 | 32.82(6.57,73.53) | 1.04(0.90,1.18) |
| Antigua and Barbuda | 4.99(4.56,5.41) | 4.63(4.07,5.17) | 5 | -7.15(-18.33,3.90) | -0.14(-0.32,0.03) |
| Argentina | 5.46(5.10,5.73) | 5.69(5.18,6.08) | 3827 | 4.16(-1.84,10.53) | -0.10(-0.23,0.02) |
| Armenia | 3.56(3.17,4.00) | 2.98(2.64,3.27) | 157 | -16.38(-27.75,-4.10) | -0.30(-0.54,-0.06) |
| Australia | 4.62(4.25,4.85) | 4.61(3.99,4.96) | 2687 | -0.25(-6.87,5.17) | 0.04(-0.09,0.18) |
| Austria | 4.56(4.15,4.81) | 5.52(4.91,5.93) | 1294 | 20.87(14.08,27.44) | 0.64(0.46,0.82) |
| Azerbaijan | 4.30(3.38,5.47) | 4.41(3.35,5.59) | 331 | 2.59(-19.25,35.35) | 0.97(0.66,1.28) |
| Bahrain | 6.73(5.78,8.12) | 6.30(5.17,7.60) | 26 | -6.33(-24.30,21.53) | 0.05(-0.17,0.27) |
| Bangladesh | 4.54(3.50,5.80) | 7.62(5.56,9.99) | 6705 | 68.00(27.86,127.53) | 1.15(0.98,1.32) |
| Barbados | 4.26(3.98,4.55) | 5.53(5.00,6.05) | 31 | 30.02(17.79,41.68) | 0.52(0.32,0.72) |
| Belarus | 3.69(3.30,4.13) | 5.07(4.48,5.59) | 862 | 37.57(18.35,56.77) | 0.94(0.84,1.05) |
| Belgium | 4.49(4.06,4.76) | 5.10(4.42,5.52) | 1524 | 13.51(6.90,20.59) | 0.53(0.46,0.60) |
| Belize | 3.97(3.63,4.25) | 5.41(4.89,6.02) | 13 | 36.22(22.80,52.18) | 0.82(0.43,1.21) |
| Benin | 3.76(2.91,4.67) | 3.87(2.90,4.88) | 175 | 2.82(-21.61,33.09) | -0.09(-0.19,0.02) |
| Bermuda | 4.64(4.15,5.13) | 7.17(5.94,8.38) | 11 | 54.62(30.31,80.60) | 0.77(0.56,0.97) |
| Bhutan | 4.50(3.23,5.82) | 5.37(4.13,6.67) | 31 | 19.43(-7.83,61.23) | 0.32(0.20,0.44) |
| Bolivarian Republic of Venezuela | 4.62(4.26,4.86) | 5.80(5.23,6.39) | 1362 | 25.61(12.77,39.41) | 0.61(0.39,0.84) |
| Bosnia and Herzegovina | 5.58(4.87,6.31) | 5.26(4.41,6.04) | 340 | -5.72(-19.57,8.28) | 0.03(-0.07,0.13) |
| Botswana | 2.51(1.97,3.04) | 3.23(2.58,3.82) | 52 | 28.77(6.40,62.73) | 0.15(-0.22,0.53) |
| Brazil | 4.14(3.74,4.40) | 4.36(3.77,4.69) | 10904 | 5.26(-0.99,11.26) | 0.32(0.27,0.36) |
| Brunei Darussalam | 4.32(3.00,5.80) | 4.00(3.05,5.28) | 11 | -7.42(-29.79,25.44) | -0.18(-0.29,-0.06) |
| Bulgaria | 5.27(4.91,5.52) | 5.09(4.71,5.43) | 834 | -3.33(-10.21,3.45) | -0.04(-0.08,0.00) |
| Burkina Faso | 4.76(3.44,6.15) | 5.00(3.59,6.39) | 405 | 5.10(-16.28,34.66) | 0.01(-0.21,0.22) |
| Burundi | 2.84(2.04,3.77) | 4.52(3.15,5.79) | 146 | 59.16(21.16,111.01) | 1.81(1.61,2.01) |
| Cambodia | 2.42(1.92,3.03) | 3.32(2.66,4.04) | 361 | 37.13(8.81,74.06) | 1.03(0.94,1.11) |
| Cameroon | 6.03(4.86,7.26) | 6.94(5.57,8.56) | 581 | 15.16(-7.28,45.68) | 0.58(0.43,0.72) |
| Canada | 4.72(4.33,4.96) | 5.56(4.91,6.02) | 4940 | 17.77(11.38,24.77) | 0.46(0.36,0.55) |
| Central African Republic | 2.52(1.82,3.34) | 2.91(2.05,3.73) | 47 | 15.64(-8.73,53.39) | -0.02(-0.24,0.20) |
| Chad | 5.14(3.82,6.46) | 5.60(4.16,7.14) | 223 | 8.94(-12.83,38.69) | 0.17(0.01,0.33) |
| Chile | 5.19(4.81,5.47) | 5.55(4.93,5.96) | 1562 | 6.83(-0.78,13.41) | 0.19(0.14,0.25) |
| China | 4.69(4.00,5.67) | 3.78(3.15,4.51) | 82927 | -19.37(-35.33,-0.26) | -0.90(-1.07,-0.73) |
| Colombia | 3.90(3.57,4.12) | 4.21(3.78,4.50) | 2453 | 7.92(1.18,14.08) | 0.00(-0.12,0.11) |
| Commonwealth of the Bahamas | 4.52(4.19,4.86) | 6.67(6.11,7.25) | 22 | 47.76(33.68,61.08) | 1.36(1.19,1.52) |
| Comoros | 1.86(1.40,2.43) | 2.16(1.64,2.68) | 14 | 15.60(-11.12,55.49) | 0.47(0.27,0.68) |
| Congo | 2.73(2.15,3.45) | 4.60(3.64,5.69) | 82 | 68.36(34.20,114.81) | 1.61(1.47,1.75) |
| Cook Islands | 5.87(4.72,7.26) | 8.28(6.77,10.00) | 2 | 41.02(12.05,74.62) | 0.63(0.47,0.79) |
| Costa Rica | 3.86(3.54,4.11) | 4.89(4.44,5.22) | 282 | 26.53(17.37,35.78) | 0.31(0.14,0.48) |
| Croatia | 4.92(4.57,5.20) | 5.12(4.63,5.43) | 525 | 3.91(-2.45,10.80) | 0.24(0.20,0.28) |
| Cuba | 3.49(3.24,3.68) | 4.38(3.89,4.93) | 942 | 25.75(10.56,40.49) | 0.79(0.68,0.89) |
| Cyprus | 5.18(3.94,6.74) | 5.93(4.83,7.05) | 118 | 14.47(-12.47,49.56) | 0.46(0.36,0.55) |
| Czech Republic | 4.45(4.19,4.67) | 4.65(4.23,4.94) | 1172 | 4.63(-2.26,11.68) | 0.43(0.34,0.53) |
| Democratic People's Republic of Korea | 4.18(3.31,5.26) | 5.30(4.15,6.81) | 1489 | 27.03(-1.37,63.80) | 0.70(0.57,0.82) |
| Democratic Republic of the Congo | 3.04(2.22,3.87) | 4.36(3.03,5.73) | 1148 | 43.18(9.24,88.98) | 1.01(0.76,1.27) |
| Denmark | 3.54(3.28,3.75) | 5.21(4.63,5.63) | 786 | 47.31(38.16,58.23) | 1.28(1.20,1.36) |
| Djibouti | 3.30(2.51,4.19) | 3.99(3.10,5.01) | 18 | 20.85(-2.13,53.69) | 0.60(0.47,0.74) |
| Dominica | 4.67(3.76,5.70) | 5.48(4.50,6.68) | 6 | 17.39(-4.75,50.61) | 0.16(0.02,0.29) |
| Dominican Republic | 4.74(4.09,5.35) | 7.25(6.10,8.60) | 641 | 53.09(27.91,85.06) | 1.25(0.85,1.66) |
| Ecuador | 4.14(3.85,4.35) | 4.69(4.36,4.99) | 818 | 13.09(5.71,21.71) | -0.10(-0.58,0.38) |
| Egypt | 5.86(4.95,6.82) | 7.62(6.10,9.58) | 2896 | 29.99(3.41,64.77) | 0.26(0.03,0.50) |
| El Salvador | 4.53(3.94,5.40) | 7.03(5.91,8.31) | 427 | 55.22(33.61,77.38) | 1.19(1.02,1.35) |
| Equatorial Guinea | 3.19(2.39,4.24) | 5.94(4.90,7.25) | 22 | 85.89(43.89,148.82) | 2.10(1.91,2.29) |
| Eritrea | 3.14(2.33,4.24) | 4.48(3.43,5.56) | 123 | 42.72(12.27,86.94) | 0.86(0.70,1.01) |
| Estonia | 4.37(4.03,4.68) | 4.84(4.23,5.20) | 170 | 10.53(1.36,22.25) | 0.03(-0.09,0.15) |
| Ethiopia | 3.41(2.39,4.82) | 7.49(5.41,9.85) | 2788 | 119.92(57.80,207.85) | 2.43(2.33,2.53) |
| Federated States of Micronesia | 7.39(6.12,8.93) | 6.63(5.55,7.93) | 3 | -10.20(-27.05,12.52) | -0.42(-0.50,-0.33) |
| Fiji | 4.26(3.42,5.18) | 5.65(4.63,6.71) | 31 | 32.46(2.70,65.44) | 0.66(0.58,0.74) |
| Finland | 4.32(3.94,4.60) | 5.96(5.13,6.48) | 979 | 37.86(27.56,46.60) | 0.98(0.90,1.06) |
| France | 4.21(3.84,4.41) | 5.08(4.45,5.43) | 9598 | 20.69(14.74,26.43) | 0.48(0.40,0.56) |
| Gabon | 3.05(2.41,3.97) | 4.70(3.77,5.91) | 48 | 53.76(23.09,95.37) | 1.07(0.95,1.19) |
| Georgia | 5.01(4.42,5.76) | 4.65(4.05,5.14) | 302 | -7.14(-25.16,7.14) | 0.08(-0.18,0.34) |
| Germany | 3.92(3.57,4.12) | 5.64(4.90,6.08) | 14498 | 43.99(36.37,50.46) | 0.93(0.74,1.13) |
| Ghana | 3.77(3.06,4.62) | 4.85(3.95,5.80) | 635 | 28.54(2.24,63.26) | 0.53(0.34,0.72) |
| Greece | 4.62(4.20,4.89) | 5.24(4.68,5.62) | 1791 | 13.54(6.64,20.79) | 0.40(0.26,0.55) |
| Greenland | 8.20(5.63,12.28) | 10.53(7.97,13.96) | 4 | 28.44(0.78,71.72) | 0.92(0.83,1.01) |
| Grenada | 4.29(3.91,4.61) | 3.73(3.26,4.14) | 6 | -12.96(-22.72,-0.72) | -0.23(-0.40,-0.05) |
| Guam | 4.48(3.92,5.05) | 4.45(3.87,4.99) | 9 | -0.61(-12.90,12.81) | -0.38(-0.52,-0.23) |
| Guatemala | 4.86(4.47,5.17) | 4.33(3.91,4.88) | 387 | -10.88(-20.62,1.59) | -0.47(-0.60,-0.35) |
| Guinea | 4.83(3.69,6.21) | 5.05(4.00,6.10) | 253 | 4.69(-17.39,37.99) | -0.36(-0.59,-0.12) |
| Guinea-Bissau | 3.76(3.09,4.63) | 5.27(4.20,6.56) | 29 | 40.12(9.28,80.48) | 0.73(0.59,0.88) |
| Guyana | 4.04(3.64,4.40) | 5.67(4.92,6.47) | 33 | 40.38(19.87,65.30) | 0.56(0.33,0.79) |
| Haiti | 3.56(2.83,4.62) | 5.10(4.12,6.39) | 248 | 43.22(12.17,88.04) | 0.88(0.82,0.95) |
| Honduras | 5.48(4.48,6.57) | 8.49(6.83,10.95) | 458 | 54.92(21.03,95.13) | 1.07(0.96,1.18) |
| Hungary | 4.27(4.02,4.48) | 4.71(4.36,5.00) | 1046 | 10.32(4.04,17.14) | 0.40(0.29,0.51) |
| Iceland | 5.72(5.15,6.16) | 7.77(6.65,8.56) | 53 | 35.87(23.47,48.38) | 0.91(0.80,1.03) |
| India | 4.35(3.29,5.36) | 5.86(4.62,7.25) | 55904 | 34.77(4.20,75.25) | 0.88(0.78,0.97) |
| Indonesia | 2.75(2.17,3.40) | 3.46(2.72,4.44) | 6431 | 26.07(-5.39,65.45) | 0.63(0.53,0.74) |
| Iraq | 4.91(3.75,6.87) | 8.00(6.23,9.75) | 1211 | 62.87(11.78,129.48) | 1.44(1.35,1.53) |
| Ireland | 4.69(4.32,4.97) | 5.80(5.06,6.31) | 551 | 23.69(14.12,33.66) | 0.55(0.43,0.66) |
| Islamic Republic of Iran | 4.81(3.91,6.08) | 4.61(3.81,5.48) | 2687 | -4.20(-28.84,26.83) | 0.01(-0.08,0.10) |
| Israel | 4.80(4.42,5.08) | 4.90(4.30,5.36) | 714 | 2.20(-5.99,10.58) | 0.17(0.13,0.22) |
| Italy | 4.68(4.15,4.97) | 5.00(4.29,5.45) | 9905 | 6.92(0.59,14.28) | 0.30(0.11,0.49) |
| Jamaica | 3.60(3.29,3.90) | 4.57(3.98,5.12) | 151 | 26.78(11.80,44.45) | 0.70(0.54,0.86) |
| Japan | 3.20(2.83,3.41) | 3.96(3.28,4.40) | 21766 | 23.87(13.46,34.37) | 0.70(0.62,0.78) |
| Jordan | 4.89(3.77,6.37) | 5.09(4.02,6.10) | 299 | 4.17(-20.79,42.12) | 0.05(-0.06,0.16) |
| Kazakhstan | 4.11(3.80,4.35) | 5.60(4.92,6.10) | 860 | 36.31(23.55,47.17) | 0.98(0.77,1.19) |
| Kenya | 3.36(2.46,4.31) | 3.72(2.96,4.60) | 749 | 10.62(-14.52,48.34) | 0.08(-0.13,0.29) |
| Kingdom of Eswatini | 2.89(2.24,3.64) | 2.58(2.07,3.13) | 17 | -10.58(-30.15,14.95) | -0.69(-0.94,-0.44) |
| Kiribati | 2.76(2.21,3.24) | 2.98(2.36,3.58) | 2 | 7.97(-12.15,37.03) | 0.05(-0.01,0.11) |
| Kuwait | 9.50(8.55,10.30) | 4.40(3.94,4.85) | 49 | -53.70(-57.63,-49.14) | -1.72(-2.40,-1.05) |
| Kyrgyzstan | 3.78(3.42,4.15) | 3.68(3.38,3.97) | 115 | -2.62(-11.27,9.48) | -0.22(-0.47,0.03) |
| Lao People's Democratic Republic | 3.06(2.35,4.14) | 5.18(4.20,6.23) | 193 | 69.33(28.93,122.53) | 1.59(1.54,1.65) |
| Latvia | 4.51(4.12,4.89) | 4.63(4.18,4.96) | 233 | 2.73(-6.01,14.51) | 0.03(-0.10,0.15) |
| Lebanon | 2.06(1.41,2.81) | 3.40(2.70,4.23) | 225 | 64.98(26.71,141.51) | 1.82(1.44,2.20) |
| Lesotho | 3.28(2.46,4.18) | 3.15(2.52,3.90) | 35 | -3.88(-25.95,24.35) | -0.99(-1.38,-0.61) |
| Liberia | 4.90(3.88,6.06) | 5.76(4.47,7.22) | 92 | 17.59(-6.24,53.75) | 0.44(0.27,0.61) |
| Libya | 4.19(3.37,5.27) | 5.08(3.96,6.59) | 206 | 21.26(-4.31,58.18) | 0.49(0.39,0.59) |
| Lithuania | 3.77(3.47,4.02) | 4.68(4.23,5.02) | 334 | 24.05(15.77,34.44) | 0.85(0.71,1.00) |
| Luxembourg | 5.53(5.12,5.88) | 6.17(5.49,6.70) | 74 | 11.58(0.83,21.30) | 0.46(0.36,0.55) |
| Madagascar | 6.43(4.86,8.12) | 8.22(5.94,10.40) | 554 | 27.87(2.05,66.24) | 0.70(0.52,0.87) |
| Malawi | 3.91(2.90,5.02) | 5.11(3.78,6.23) | 309 | 30.66(2.92,70.79) | 0.72(0.45,0.99) |
| Malaysia | 4.14(3.39,5.09) | 4.96(4.19,5.91) | 1282 | 19.80(-5.43,46.99) | 0.15(-0.02,0.32) |
| Maldives | 3.78(3.08,4.67) | 10.08(7.95,11.62) | 20 | 166.87(111.04,242.77) | 2.68(2.58,2.79) |
| Mali | 3.34(2.51,4.20) | 4.10(3.08,5.06) | 274 | 22.64(-0.77,60.04) | 0.22(0.09,0.36) |
| Malta | 5.28(4.80,5.65) | 5.34(4.57,5.83) | 63 | 1.25(-8.26,11.30) | 0.04(-0.13,0.21) |
| Marshall Islands | 7.97(6.48,9.83) | 9.70(7.91,11.75) | 1 | 21.77(-4.78,51.24) | 0.38(0.28,0.49) |
| Mauritania | 4.02(3.16,4.97) | 6.40(4.78,8.04) | 112 | 59.03(24.77,105.51) | 1.22(1.07,1.37) |
| Mauritius | 4.41(4.02,4.67) | 5.20(4.67,5.62) | 85 | 18.05(9.61,27.58) | 0.10(-0.04,0.23) |
| Mexico | 4.41(4.08,4.66) | 4.30(4.01,4.57) | 5401 | -2.45(-8.09,4.10) | -0.29(-0.41,-0.17) |
| Mongolia | 3.76(3.13,4.48) | 3.50(2.84,4.22) | 54 | -7.14(-26.55,13.76) | -0.50(-0.64,-0.36) |
| Montenegro | 6.14(5.29,7.25) | 6.04(5.12,7.12) | 67 | -1.65(-17.12,17.80) | 0.03(-0.09,0.16) |
| Morocco | 5.44(4.43,6.65) | 7.64(5.91,9.89) | 2086 | 40.36(5.89,90.84) | 0.99(0.82,1.15) |
| Mozambique | 2.69(1.90,3.47) | 2.95(2.24,3.61) | 310 | 9.65(-15.34,45.67) | -0.08(-0.28,0.12) |
| Myanmar | 3.89(3.05,4.95) | 5.60(4.53,7.07) | 2156 | 44.00(13.78,86.30) | 1.22(1.12,1.32) |
| Namibia | 2.57(2.06,3.10) | 2.95(2.41,3.52) | 40 | 14.78(-5.98,42.40) | 0.39(0.25,0.53) |
| Nepal | 3.05(2.26,3.83) | 4.10(3.22,5.21) | 836 | 34.28(3.94,78.48) | 0.27(0.06,0.48) |
| Netherlands | 5.55(5.03,5.85) | 5.28(4.69,5.66) | 2315 | -4.86(-9.77,0.65) | 0.11(-0.02,0.25) |
| New Zealand | 4.10(3.74,4.36) | 5.29(4.70,5.71) | 525 | 29.05(20.17,37.40) | 0.72(0.66,0.78) |
| Nicaragua | 4.15(3.47,4.91) | 4.71(3.86,5.61) | 184 | 13.46(-7.70,43.75) | 1.00(0.74,1.25) |
| Niger | 4.52(3.27,5.94) | 5.32(3.57,7.19) | 270 | 17.71(-7.44,59.27) | 0.30(0.23,0.37) |
| Nigeria | 3.85(3.07,4.76) | 5.72(4.47,7.30) | 3816 | 48.39(12.70,91.70) | 1.26(1.17,1.35) |
| Northern Mariana Islands | 5.84(4.70,7.64) | 7.36(6.16,8.62) | 2 | 26.04(-2.58,63.91) | 0.66(0.53,0.80) |
| Norway | 4.17(3.72,4.44) | 5.51(4.78,6.00) | 677 | 32.10(22.08,41.24) | 1.25(1.09,1.42) |
| Oman | 4.88(3.69,6.25) | 7.81(5.82,9.63) | 87 | 60.09(2.76,129.59) | 2.29(2.04,2.54) |
| Pakistan | 3.54(2.69,4.52) | 4.39(3.59,5.36) | 4633 | 24.13(-1.31,58.13) | 0.60(0.46,0.75) |
| Palestine | 4.11(3.25,5.26) | 4.91(4.03,5.74) | 92 | 19.63(-7.73,56.27) | 0.39(0.33,0.46) |
| Panama | 3.66(3.34,3.91) | 4.96(4.43,5.38) | 230 | 35.45(25.53,48.03) | 0.63(0.45,0.82) |
| Papua New Guinea | 5.89(4.27,7.56) | 5.75(4.15,7.40) | 187 | -2.42(-24.04,26.01) | -0.46(-0.59,-0.34) |
| Paraguay | 3.49(3.00,4.10) | 5.65(4.32,6.47) | 259 | 61.59(23.26,97.11) | 1.53(1.47,1.59) |
| Peru | 3.64(3.18,4.19) | 6.32(5.23,7.46) | 2194 | 73.67(42.20,108.74) | 0.90(0.66,1.13) |
| Philippines | 3.77(3.21,4.32) | 4.42(3.84,5.08) | 2676 | 17.22(-0.47,39.38) | 0.44(0.32,0.57) |
| Plurinational State of Bolivia | 4.58(3.65,5.82) | 6.88(5.74,8.17) | 549 | 50.00(17.76,90.40) | 1.32(1.20,1.44) |
| Poland | 4.59(4.27,4.91) | 5.05(4.57,5.35) | 4144 | 10.03(2.98,17.36) | 0.13(0.01,0.24) |
| Portugal | 3.77(3.50,3.98) | 4.33(3.73,4.68) | 1443 | 14.79(5.12,22.90) | 0.65(0.50,0.80) |
| Principality of Monaco | 5.71(4.48,6.95) | 7.46(5.71,9.77) | 10 | 30.63(-4.25,74.11) | 0.76(0.58,0.93) |
| Puerto Rico | 3.86(3.56,4.06) | 4.28(3.80,4.56) | 451 | 10.97(3.56,16.60) | -0.04(-0.23,0.14) |
| Qatar | 8.61(6.81,10.97) | 5.26(4.11,6.80) | 16 | -38.96(-52.81,-18.46) | -2.16(-2.43,-1.89) |
| Republic of CÃ´te d'Ivoire | 4.29(3.49,5.34) | 4.89(3.93,6.07) | 400 | 14.15(-7.28,47.89) | 0.08(-0.19,0.34) |
| Republic of Cabo Verde | 4.64(3.54,5.78) | 4.61(3.74,5.70) | 20 | -0.76(-24.39,25.04) | -0.07(-0.27,0.13) |
| Republic of Korea | 3.99(3.18,5.57) | 4.41(3.59,5.07) | 4521 | 10.40(-24.72,44.57) | 0.56(0.46,0.66) |
| Republic of Moldova | 4.39(4.04,4.70) | 2.72(2.43,2.95) | 174 | -38.07(-43.59,-32.30) | -1.79(-2.00,-1.59) |
| Republic of Nauru | 10.04(8.09,13.01) | 10.07(8.18,12.59) | 0 | 0.30(-22.34,29.71) | 0.01(-0.19,0.21) |
| Republic of Niue | 5.33(4.40,6.43) | 5.12(4.11,6.52) | 0 | -4.00(-25.64,19.49) | -0.32(-0.44,-0.21) |
| Republic of Palau | 5.60(4.60,6.70) | 5.77(4.77,6.98) | 1 | 3.05(-17.90,30.08) | -0.09(-0.20,0.01) |
| Republic of San Marino | 3.45(2.73,4.23) | 2.33(1.76,2.81) | 2 | -32.33(-44.70,-11.97) | -1.54(-1.71,-1.36) |
| Republic of the Gambia | 2.74(2.10,3.40) | 3.66(2.67,4.60) | 36 | 33.48(7.15,79.50) | 0.51(0.20,0.82) |
| Romania | 4.57(4.30,4.78) | 4.94(4.52,5.26) | 2039 | 8.10(2.06,14.41) | 0.35(0.24,0.46) |
| Russian Federation | 3.95(3.63,4.20) | 4.60(4.17,4.90) | 11441 | 16.59(8.89,25.73) | 0.40(0.25,0.54) |
| Rwanda | 3.71(2.81,5.07) | 6.13(4.67,7.74) | 300 | 65.31(26.89,120.05) | 1.72(1.50,1.94) |
| Saint Kitts and Nevis | 7.26(6.79,7.77) | 7.64(6.98,8.23) | 4 | 5.24(-3.28,17.30) | -0.07(-0.28,0.14) |
| Saint Lucia | 5.24(4.77,5.67) | 4.76(4.18,5.37) | 12 | -9.21(-21.11,4.03) | -0.54(-0.67,-0.41) |
| Saint Vincent and the Grenadines | 3.93(3.60,4.28) | 3.71(3.19,4.17) | 6 | -5.70(-17.41,9.76) | -0.32(-0.54,-0.10) |
| Samoa | 3.21(2.52,4.12) | 3.99(3.19,4.92) | 6 | 24.47(-0.35,54.31) | 0.22(0.09,0.36) |
| Sao Tome and Principe | 3.97(3.32,4.67) | 5.54(4.46,6.74) | 5 | 39.52(9.06,76.57) | 1.16(1.04,1.27) |
| Saudi Arabia | 8.65(6.67,11.35) | 11.97(9.85,14.41) | 752 | 38.42(-3.11,93.04) | 1.70(1.34,2.06) |
| Senegal | 3.01(2.30,3.79) | 4.58(3.54,5.53) | 332 | 52.29(22.53,92.65) | 1.20(1.00,1.40) |
| Serbia | 5.52(4.64,6.58) | 5.97(5.15,6.93) | 931 | 8.09(-12.20,30.65) | 0.17(0.13,0.21) |
| Seychelles | 6.59(5.53,7.97) | 6.00(4.89,7.49) | 6 | -8.92(-25.07,9.15) | -0.59(-0.72,-0.46) |
| Sierra Leone | 4.44(3.35,5.71) | 5.73(4.47,7.08) | 151 | 28.98(-0.70,70.72) | 0.85(0.60,1.10) |
| Singapore | 3.77(3.52,4.00) | 3.62(3.09,3.96) | 323 | -4.12(-12.41,5.44) | -0.29(-0.37,-0.21) |
| Slovakia | 4.55(3.96,5.39) | 4.45(3.80,4.97) | 449 | -2.05(-15.68,15.25) | 0.03(-0.01,0.06) |
| Slovenia | 5.09(4.72,5.40) | 4.98(4.39,5.34) | 277 | -2.14(-10.70,4.69) | -0.16(-0.32,0.01) |
| Socialist Republic of Viet Nam | 3.70(2.86,5.02) | 4.46(3.25,5.95) | 4371 | 20.69(-7.96,58.50) | 0.64(0.51,0.76) |
| Solomon Islands | 7.05(4.73,9.43) | 7.35(5.41,9.09) | 23 | 4.21(-20.62,37.82) | -0.07(-0.33,0.19) |
| Somalia | 1.83(1.12,2.63) | 2.26(1.61,2.91) | 154 | 23.24(-7.52,80.33) | 0.57(0.37,0.77) |
| South Africa | 2.99(2.44,3.53) | 3.79(3.23,4.39) | 1573 | 26.69(4.94,54.71) | 0.76(0.48,1.03) |
| South Sudan | 4.93(3.62,6.61) | 6.20(4.60,7.91) | 160 | 25.78(1.21,69.92) | 0.53(0.45,0.62) |
| Spain | 4.76(4.30,5.01) | 5.24(4.54,5.64) | 6892 | 9.89(3.91,15.31) | 0.41(0.34,0.49) |
| Sri Lanka | 4.37(3.78,5.05) | 5.08(4.24,5.75) | 1500 | 16.25(-1.33,41.90) | 0.28(0.19,0.36) |
| Sudan | 5.05(4.11,6.20) | 5.79(4.45,7.58) | 928 | 14.58(-13.10,59.57) | 0.32(0.24,0.41) |
| Suriname | 3.57(3.05,4.31) | 4.33(3.63,5.02) | 25 | 21.12(-1.19,50.45) | 0.46(0.33,0.58) |
| Sweden | 4.32(3.96,4.59) | 4.58(4.02,4.95) | 1319 | 5.93(-1.52,12.18) | 0.08(-0.11,0.28) |
| Switzerland | 4.14(3.73,4.42) | 4.91(4.20,5.33) | 1155 | 18.49(9.47,26.40) | 0.61(0.54,0.68) |
| Syrian Arab Republic | 5.49(4.20,7.16) | 7.27(5.67,9.07) | 622 | 32.44(-7.63,90.24) | 0.54(0.36,0.72) |
| Taiwan (Province of China) | 5.13(4.74,5.38) | 5.40(4.78,5.84) | 2482 | 5.11(-3.34,13.79) | 0.15(0.09,0.21) |
| Tajikistan | 5.84(4.60,7.36) | 8.42(6.69,10.72) | 280 | 44.17(9.34,96.08) | 0.93(0.63,1.22) |
| Thailand | 5.17(4.15,6.36) | 4.74(3.91,5.97) | 5554 | -8.34(-23.15,13.26) | -0.75(-1.04,-0.45) |
| The former Yugoslav Republic of Macedonia | 5.93(5.25,6.73) | 6.05(4.88,6.91) | 198 | 2.00(-17.37,25.21) | 0.13(0.02,0.24) |
| Timor-Leste | 4.16(3.12,5.25) | 4.39(3.44,5.60) | 33 | 5.65(-18.71,35.24) | 0.00(-0.08,0.08) |
| Togo | 4.25(3.25,5.25) | 4.64(3.54,5.82) | 130 | 9.35(-11.83,41.07) | -0.02(-0.25,0.21) |
| Tokelau | 4.92(3.95,6.11) | 4.79(3.86,6.09) | 0 | -2.61(-22.52,23.39) | -0.27(-0.40,-0.15) |
| Tonga | 4.58(3.62,5.83) | 5.45(4.47,6.59) | 4 | 18.99(-6.35,47.97) | 0.27(0.16,0.38) |
| Trinidad and Tobago | 5.83(5.46,6.12) | 6.21(5.69,6.73) | 94 | 6.62(-2.44,16.92) | -0.05(-0.22,0.12) |
| Tunisia | 4.25(3.26,5.61) | 4.19(3.44,5.05) | 467 | -1.56(-20.13,27.24) | -0.44(-0.55,-0.34) |
| Turkey | 5.24(3.82,6.81) | 5.49(4.58,6.87) | 4677 | 4.79(-21.35,39.93) | -0.13(-0.32,0.07) |
| Turkmenistan | 3.36(3.03,3.67) | 4.01(3.37,4.51) | 121 | 19.39(0.60,37.90) | 0.54(0.41,0.66) |
| Tuvalu | 7.46(6.09,9.12) | 9.15(7.29,11.02) | 1 | 22.57(-1.55,56.39) | 0.43(0.30,0.55) |
| Uganda | 4.22(2.91,5.69) | 5.14(3.77,6.55) | 634 | 21.86(-5.04,62.54) | 0.60(0.39,0.81) |
| Ukraine | 2.84(2.55,3.11) | 3.35(2.92,3.79) | 2753 | 17.89(2.76,35.17) | 0.94(0.79,1.09) |
| United Arab Emirates | 5.64(4.02,8.26) | 5.74(4.25,7.24) | 61 | 1.64(-28.58,50.90) | 0.20(-0.04,0.44) |
| United Kingdom of Great Britain and Northern Ireland | 4.47(4.10,4.69) | 5.83(5.20,6.21) | 9745 | 30.22(24.33,36.10) | 0.96(0.79,1.13) |
| United Republic of Tanzania | 3.32(2.50,4.09) | 4.12(3.04,5.16) | 1011 | 24.01(-2.32,62.93) | 0.71(0.60,0.82) |
| United States of America | 3.84(3.39,4.08) | 6.38(5.53,6.88) | 42105 | 66.17(58.55,75.07) | 1.67(1.57,1.76) |
| United States Virgin Islands | 7.66(6.79,8.59) | 6.77(5.76,7.82) | 16 | -11.53(-25.15,5.10) | -1.01(-1.34,-0.67) |
| Uruguay | 5.06(4.70,5.34) | 5.33(4.80,5.71) | 365 | 5.37(-1.04,12.84) | 0.11(-0.01,0.23) |
| Uzbekistan | 3.65(3.30,4.00) | 3.94(3.61,4.19) | 688 | 8.20(-3.73,22.41) | 0.20(0.13,0.28) |
| Vanuatu | 3.47(2.49,4.54) | 3.41(2.66,4.14) | 5 | -1.67(-22.99,26.66) | -0.40(-0.63,-0.16) |
| Yemen | 3.99(3.14,4.87) | 4.28(3.42,5.23) | 406 | 7.53(-13.78,41.65) | 0.20(0.13,0.28) |
| Zambia | 4.25(3.22,5.32) | 5.45(4.24,6.86) | 223 | 28.34(0.50,69.33) | 0.55(0.40,0.69) |
| Zimbabwe | 2.84(2.32,3.39) | 2.12(1.72,2.76) | 163 | -25.31(-43.33,-2.67) | -1.95(-2.41,-1.49) |

**Table S3:** Incidence data for countries and territories related to Parkinson disease. It includes location-based total numbers, age-standardized rates (ASR) per 100,000 for 1990 and 2023, percentage changes, and EAPC from 1990 to 2023.

| **Regions** | **ASIR, 1990 (95% CI)** | **ASIR, 2023 (95% CI)** | **Numbers, 2023** | **PC 1990-2023** | **EAPC, %, 1990-2023** |
| --- | --- | --- | --- | --- | --- |
| Afghanistan | 10.68(9.40,12.42) | 14.25(12.51,16.34) | 1530 | 33.43(18.40,48.85) | 0.91(0.86,0.95) |
| Albania | 10.04(8.92,11.15) | 15.09(13.23,17.20) | 699 | 50.36(36.44,67.19) | 1.24(1.19,1.29) |
| Algeria | 11.90(10.35,14.13) | 17.65(14.94,20.61) | 6295 | 48.34(31.57,68.12) | 1.15(1.12,1.18) |
| American Samoa | 16.58(14.67,18.64) | 19.46(17.29,22.60) | 8 | 17.36(5.66,30.87) | 0.35(0.30,0.40) |
| Andorra | 12.23(10.67,14.09) | 11.41(9.96,13.19) | 20 | -6.74(-17.13,5.59) | -0.26(-0.35,-0.17) |
| Angola | 6.72(6.02,7.49) | 8.81(7.98,9.89) | 930 | 31.11(20.07,43.70) | 0.95(0.91,0.99) |
| Antigua and Barbuda | 17.03(14.52,19.48) | 20.08(16.87,23.42) | 23 | 17.89(4.23,34.73) | 0.47(0.41,0.53) |
| Argentina | 15.08(13.75,16.55) | 18.61(17.01,20.04) | 11818 | 23.44(9.50,37.49) | 0.44(0.17,0.71) |
| Armenia | 5.86(5.52,6.31) | 5.88(5.37,6.36) | 309 | 0.41(-6.73,9.62) | -0.03(-0.13,0.07) |
| Australia | 15.95(14.77,17.04) | 17.65(15.21,20.71) | 9119 | 10.66(-1.80,28.45) | 0.17(-0.12,0.46) |
| Austria | 11.18(10.32,12.24) | 14.55(13.42,16.19) | 2997 | 30.16(18.24,43.54) | 0.91(0.83,0.98) |
| Azerbaijan | 6.34(5.84,6.94) | 7.54(7.04,7.99) | 664 | 18.96(12.02,24.97) | 0.53(0.48,0.58) |
| Bahrain | 15.56(13.72,17.81) | 20.67(16.97,24.19) | 176 | 32.80(17.07,52.33) | 0.80(0.76,0.84) |
| Bangladesh | 10.14(9.13,11.29) | 12.02(10.52,13.61) | 12978 | 18.60(8.20,30.66) | 0.53(0.48,0.58) |
| Barbados | 16.92(14.46,19.24) | 21.67(18.96,25.15) | 123 | 28.10(13.29,45.35) | 0.63(0.55,0.72) |
| Belarus | 8.75(7.87,9.90) | 8.83(7.95,9.73) | 1517 | 0.88(-5.52,8.08) | 0.14(0.08,0.21) |
| Belgium | 11.53(10.46,12.98) | 13.83(12.59,15.49) | 3532 | 19.92(11.18,29.66) | 0.66(0.60,0.72) |
| Belize | 15.79(13.40,18.76) | 23.14(20.19,26.46) | 65 | 46.54(26.65,74.86) | 1.22(1.09,1.36) |
| Benin | 6.75(6.08,7.60) | 8.87(7.94,9.87) | 440 | 31.30(19.85,44.73) | 0.86(0.82,0.89) |
| Bermuda | 16.17(13.97,19.24) | 18.87(16.53,21.39) | 29 | 16.74(3.29,35.97) | 0.40(0.34,0.46) |
| Bhutan | 7.74(6.91,8.79) | 11.97(10.64,13.38) | 74 | 54.59(41.19,71.63) | 1.47(1.42,1.53) |
| Bolivarian Republic of Venezuela | 15.99(13.98,18.29) | 17.63(14.95,20.75) | 5249 | 10.29(-3.13,22.22) | 0.29(0.10,0.49) |
| Bosnia and Herzegovina | 9.84(8.87,10.93) | 13.68(12.17,15.23) | 896 | 38.99(26.33,58.50) | 1.15(1.11,1.19) |
| Botswana | 8.03(7.28,9.03) | 8.46(7.47,9.45) | 137 | 5.32(-3.90,16.01) | 0.19(0.08,0.30) |
| Brazil | 23.49(19.92,27.88) | 29.74(25.13,35.13) | 78549 | 26.63(19.91,33.81) | 0.12(-0.10,0.34) |
| Brunei Darussalam | 10.65(9.54,12.11) | 12.01(10.47,13.89) | 43 | 12.81(2.58,23.58) | 0.31(0.27,0.34) |
| Bulgaria | 14.34(12.85,15.79) | 13.48(12.28,14.56) | 2295 | -5.98(-16.60,8.83) | -0.77(-1.06,-0.48) |
| Burkina Faso | 7.24(6.48,8.05) | 7.94(7.20,8.86) | 714 | 9.68(1.45,19.16) | 0.26(0.21,0.31) |
| Burundi | 7.58(6.79,8.41) | 8.76(7.89,9.77) | 346 | 15.61(6.11,24.69) | 0.51(0.48,0.53) |
| Cambodia | 7.92(7.17,8.86) | 11.74(10.56,13.35) | 1519 | 48.20(35.38,60.68) | 1.27(1.23,1.31) |
| Cameroon | 8.73(7.87,9.69) | 9.95(8.90,11.15) | 1034 | 13.93(5.44,24.34) | 0.40(0.39,0.42) |
| Canada | 9.82(9.11,10.75) | 14.89(13.60,16.67) | 12119 | 51.70(39.42,66.04) | 2.03(1.67,2.39) |
| Central African Republic | 6.66(5.99,7.46) | 7.17(6.48,7.93) | 151 | 7.73(0.59,17.27) | 0.22(0.19,0.25) |
| Chad | 6.38(5.75,7.07) | 7.09(6.32,7.94) | 355 | 11.19(1.88,24.00) | 0.36(0.31,0.41) |
| Chile | 13.34(12.71,14.31) | 17.09(15.59,19.56) | 4808 | 28.14(16.79,46.00) | 0.64(0.20,1.08) |
| China | 13.17(11.10,15.57) | 16.26(13.86,18.88) | 379687 | 23.39(17.17,30.97) | 0.84(0.76,0.92) |
| Colombia | 11.51(10.14,13.54) | 13.79(13.15,14.45) | 8130 | 19.81(3.46,33.64) | 0.47(0.37,0.57) |
| Commonwealth of the Bahamas | 15.17(12.79,17.70) | 20.67(18.15,23.49) | 84 | 36.29(19.00,60.35) | 0.95(0.88,1.02) |
| Comoros | 8.40(7.53,9.29) | 11.39(10.37,12.51) | 68 | 35.60(24.04,48.46) | 1.01(0.97,1.05) |
| Congo | 8.23(7.42,9.04) | 9.05(8.16,10.18) | 191 | 9.93(0.57,20.30) | 0.33(0.29,0.37) |
| Cook Islands | 15.32(13.16,17.96) | 16.04(13.38,19.07) | 3 | 4.71(-5.16,15.03) | 0.01(-0.08,0.10) |
| Costa Rica | 14.20(12.42,16.22) | 18.72(16.36,21.50) | 1134 | 31.79(14.26,56.97) | 1.10(0.95,1.25) |
| Croatia | 10.73(9.85,12.29) | 13.65(12.33,15.41) | 1288 | 27.22(16.71,42.74) | 0.90(0.76,1.05) |
| Cuba | 9.66(8.75,10.84) | 18.68(16.19,21.62) | 3831 | 93.41(72.08,116.00) | 2.11(1.77,2.46) |
| Cyprus | 13.36(11.61,15.44) | 13.86(12.17,16.06) | 279 | 3.74(-7.11,15.83) | 0.05(0.01,0.09) |
| Czech Republic | 9.76(8.97,10.70) | 13.43(12.19,14.38) | 3131 | 37.62(28.93,48.95) | 1.08(1.03,1.14) |
| Democratic People's Republic of Korea | 11.75(10.27,13.56) | 14.84(12.97,17.05) | 4758 | 26.22(13.49,38.87) | 0.76(0.72,0.80) |
| Democratic Republic of the Congo | 6.29(5.64,7.09) | 7.20(6.49,8.01) | 2339 | 14.43(4.38,24.84) | 0.48(0.45,0.51) |
| Denmark | 7.92(7.54,8.33) | 12.56(11.79,13.72) | 1693 | 58.56(50.22,73.61) | 1.31(1.26,1.37) |
| Djibouti | 7.35(6.62,8.11) | 10.31(9.40,11.41) | 57 | 40.25(30.79,50.77) | 1.12(1.08,1.16) |
| Dominica | 19.16(16.65,21.91) | 24.74(21.29,28.09) | 26 | 29.13(14.43,46.59) | 0.67(0.60,0.73) |
| Dominican Republic | 15.94(13.83,18.70) | 22.91(20.04,26.88) | 2198 | 43.77(30.12,62.11) | 1.04(0.91,1.17) |
| Ecuador | 14.96(12.87,17.36) | 24.68(21.50,28.27) | 4404 | 64.92(40.74,97.91) | 1.45(1.37,1.53) |
| Egypt | 15.17(13.37,17.52) | 21.05(17.82,25.35) | 12333 | 38.76(22.68,62.93) | 1.33(1.12,1.53) |
| El Salvador | 13.90(12.11,16.29) | 18.96(16.48,22.61) | 1168 | 36.45(20.69,55.49) | 1.02(0.96,1.09) |
| Equatorial Guinea | 6.44(5.78,7.05) | 9.15(8.26,10.22) | 39 | 42.19(31.77,54.49) | 1.25(1.12,1.38) |
| Eritrea | 7.68(6.93,8.59) | 10.18(9.34,11.26) | 309 | 32.60(23.68,44.19) | 0.91(0.79,1.03) |
| Estonia | 11.47(10.77,11.99) | 13.47(12.62,14.47) | 401 | 17.52(10.59,28.13) | 0.48(0.43,0.53) |
| Ethiopia | 9.99(8.35,11.91) | 11.36(9.69,13.20) | 4596 | 13.66(6.88,21.43) | 0.43(0.36,0.49) |
| Federated States of Micronesia | 14.09(12.54,16.12) | 20.33(17.90,23.24) | 13 | 44.33(28.83,63.01) | 1.08(1.01,1.14) |
| Fiji | 14.90(13.10,16.94) | 16.48(14.19,18.99) | 116 | 10.63(-4.85,23.27) | 0.19(0.14,0.25) |
| Finland | 9.99(9.49,10.53) | 14.14(13.10,15.23) | 1989 | 41.56(31.55,52.38) | 1.11(1.04,1.19) |
| France | 12.86(11.80,13.85) | 13.90(13.30,14.65) | 21418 | 8.11(0.02,16.34) | 0.31(0.25,0.36) |
| Gabon | 9.34(8.38,10.40) | 10.89(9.88,12.24) | 118 | 16.62(6.55,27.90) | 0.46(0.41,0.52) |
| Georgia | 6.94(6.48,7.44) | 7.81(7.26,8.56) | 491 | 12.54(5.07,22.06) | 0.39(0.36,0.42) |
| Germany | 11.71(10.50,13.06) | 23.01(21.94,23.98) | 48865 | 96.52(79.46,116.32) | 1.72(1.50,1.95) |
| Ghana | 6.75(6.10,7.52) | 9.17(8.18,10.43) | 1353 | 35.91(25.25,47.65) | 0.96(0.94,0.98) |
| Greece | 10.40(9.38,11.65) | 16.37(14.89,17.65) | 4446 | 57.35(37.10,76.70) | 1.64(1.39,1.89) |
| Greenland | 10.08(9.01,11.40) | 15.91(14.21,18.10) | 9 | 57.80(42.46,74.20) | 1.48(1.43,1.53) |
| Grenada | 16.46(14.12,19.23) | 23.17(20.04,26.97) | 38 | 40.75(23.22,58.81) | 1.05(0.98,1.13) |
| Guam | 14.99(13.16,17.25) | 16.92(14.80,19.66) | 35 | 12.85(2.28,27.56) | 0.23(0.16,0.29) |
| Guatemala | 13.70(12.14,15.89) | 18.82(15.59,22.40) | 2071 | 37.35(22.93,60.01) | 0.86(0.82,0.90) |
| Guinea | 6.50(5.94,7.15) | 7.35(6.58,8.30) | 402 | 13.05(3.55,22.50) | 0.37(0.30,0.44) |
| Guinea-Bissau | 8.11(7.26,8.88) | 8.57(7.67,9.41) | 57 | 5.62(-2.15,15.33) | 0.13(0.06,0.20) |
| Guyana | 15.13(13.03,17.20) | 17.15(14.40,20.52) | 117 | 13.38(-0.80,32.85) | 0.15(0.05,0.26) |
| Haiti | 11.97(10.31,14.17) | 15.50(13.26,17.83) | 1013 | 29.51(17.37,45.80) | 0.69(0.63,0.76) |
| Honduras | 15.92(13.90,18.64) | 20.70(17.89,23.86) | 1415 | 30.05(18.17,43.46) | 0.89(0.77,1.00) |
| Hungary | 10.86(10.05,11.57) | 14.74(14.27,15.24) | 3034 | 35.79(28.19,45.17) | 0.18(-0.08,0.44) |
| Iceland | 13.18(12.06,14.92) | 19.23(17.19,22.33) | 123 | 45.94(32.59,64.07) | 1.16(1.08,1.23) |
| India | 7.32(6.14,8.57) | 9.76(8.43,11.14) | 111036 | 33.30(26.58,40.43) | 0.90(0.85,0.94) |
| Indonesia | 7.64(6.42,8.91) | 11.41(9.82,13.06) | 26299 | 49.45(42.59,57.65) | 1.27(1.23,1.32) |
| Iraq | 14.41(12.26,16.84) | 21.68(18.66,24.81) | 4957 | 50.48(31.23,70.92) | 1.34(1.31,1.38) |
| Ireland | 12.39(11.21,13.84) | 14.24(13.04,15.63) | 1276 | 14.94(5.62,24.34) | 0.60(0.54,0.67) |
| Islamic Republic of Iran | 12.54(10.58,14.57) | 18.72(16.12,21.67) | 14072 | 49.30(41.17,57.94) | 1.39(1.33,1.45) |
| Israel | 16.63(15.36,18.88) | 17.91(16.29,20.98) | 2403 | 7.74(-1.00,19.64) | 0.90(0.66,1.13) |
| Italy | 21.74(18.62,25.48) | 22.13(19.00,25.83) | 33560 | 1.83(-4.56,8.85) | 0.10(-0.03,0.23) |
| Jamaica | 13.93(11.95,15.68) | 17.38(14.90,21.01) | 564 | 24.79(11.53,45.64) | 0.57(0.48,0.66) |
| Japan | 8.02(6.74,9.46) | 11.14(9.64,12.83) | 45658 | 38.94(32.23,46.92) | 0.40(-0.19,0.99) |
| Jordan | 13.44(12.08,15.24) | 14.98(13.20,17.30) | 1119 | 11.46(0.04,23.84) | 0.17(-0.05,0.40) |
| Kazakhstan | 6.50(6.07,7.05) | 7.55(6.97,8.23) | 1375 | 16.08(8.21,23.37) | 0.55(0.46,0.64) |
| Kenya | 4.96(4.17,5.82) | 6.42(5.49,7.39) | 1476 | 29.41(22.97,35.67) | 0.84(0.80,0.88) |
| Kingdom of Eswatini | 7.63(6.83,8.58) | 8.55(7.69,9.57) | 58 | 11.99(4.24,21.26) | 0.34(0.21,0.47) |
| Kiribati | 15.67(14.01,17.78) | 18.24(16.23,20.84) | 12 | 16.39(7.23,25.74) | 0.35(0.29,0.40) |
| Kuwait | 19.28(16.70,22.39) | 19.26(16.50,22.26) | 501 | -0.10(-15.83,16.16) | -0.13(-0.19,-0.06) |
| Kyrgyzstan | 4.89(4.58,5.22) | 5.12(4.72,5.70) | 234 | 4.68(-3.26,13.35) | 0.16(0.07,0.25) |
| Lao People's Democratic Republic | 6.37(5.71,7.17) | 9.61(8.46,11.03) | 442 | 50.83(36.11,67.70) | 1.37(1.33,1.41) |
| Latvia | 8.54(7.90,9.30) | 9.53(8.72,10.26) | 425 | 11.54(4.17,19.57) | 0.45(0.37,0.52) |
| Lebanon | 12.55(10.88,14.75) | 14.96(12.91,17.78) | 923 | 19.26(2.10,38.89) | 0.51(0.49,0.54) |
| Lesotho | 6.13(5.55,6.82) | 7.98(7.25,8.76) | 97 | 30.20(19.91,41.56) | 0.82(0.71,0.93) |
| Liberia | 7.88(7.08,8.77) | 8.71(7.78,9.98) | 169 | 10.55(1.48,19.61) | 0.32(0.25,0.39) |
| Libya | 10.68(9.44,12.25) | 17.25(14.77,20.25) | 900 | 61.50(41.63,82.44) | 1.52(1.29,1.74) |
| Lithuania | 7.31(6.68,7.88) | 9.49(8.64,10.62) | 615 | 29.93(21.17,40.48) | 0.89(0.84,0.94) |
| Luxembourg | 13.03(11.30,14.91) | 15.89(14.21,17.57) | 177 | 21.94(12.03,34.74) | 0.66(0.62,0.70) |
| Madagascar | 6.01(5.44,6.64) | 6.92(6.23,7.79) | 648 | 15.05(7.11,25.91) | 0.45(0.38,0.52) |
| Malawi | 6.49(5.84,7.20) | 7.58(6.83,8.42) | 513 | 16.70(8.19,26.92) | 0.51(0.47,0.55) |
| Malaysia | 9.35(8.29,10.62) | 11.37(9.95,13.06) | 3535 | 21.59(8.64,35.67) | 0.58(0.51,0.66) |
| Maldives | 9.58(8.54,11.18) | 13.54(12.10,15.51) | 40 | 41.41(25.50,56.65) | 1.14(1.06,1.23) |
| Mali | 6.76(6.05,7.60) | 7.75(6.89,8.82) | 626 | 14.56(4.35,27.53) | 0.40(0.33,0.48) |
| Malta | 13.11(11.76,14.74) | 15.40(13.70,17.74) | 173 | 17.46(0.88,32.87) | 0.48(0.44,0.52) |
| Marshall Islands | 15.90(14.05,17.98) | 18.99(16.28,21.91) | 4 | 19.41(7.07,31.36) | 0.39(0.33,0.45) |
| Mauritania | 7.97(7.21,8.92) | 8.93(7.99,10.20) | 175 | 11.99(3.09,21.00) | 0.32(0.30,0.34) |
| Mauritius | 10.95(10.02,12.46) | 13.44(11.89,15.08) | 249 | 22.71(13.74,35.54) | 0.55(0.47,0.62) |
| Mexico | 16.97(14.39,19.89) | 19.60(16.83,22.85) | 26308 | 15.53(9.86,22.26) | 0.34(0.27,0.41) |
| Mongolia | 5.75(5.30,6.19) | 5.16(4.86,5.48) | 102 | -10.24(-15.64,-5.46) | -0.42(-0.47,-0.37) |
| Montenegro | 10.73(9.62,11.92) | 14.09(12.30,16.14) | 164 | 31.36(18.92,46.43) | 1.00(0.94,1.06) |
| Morocco | 10.31(8.73,12.09) | 15.57(13.26,18.25) | 5506 | 51.03(34.61,72.56) | 1.24(1.20,1.28) |
| Mozambique | 7.10(6.43,7.92) | 7.69(6.90,8.55) | 880 | 8.40(0.70,17.08) | 0.21(0.14,0.28) |
| Myanmar | 6.99(6.23,7.96) | 9.62(8.41,11.04) | 4428 | 37.70(25.90,52.30) | 1.00(0.97,1.04) |
| Namibia | 7.00(6.26,7.84) | 8.88(8.00,9.93) | 132 | 26.92(16.87,39.70) | 0.81(0.74,0.88) |
| Nepal | 6.89(6.13,7.70) | 10.36(9.30,11.53) | 2478 | 50.31(36.47,65.78) | 1.38(1.33,1.44) |
| Netherlands | 15.25(14.25,16.08) | 14.65(13.56,16.34) | 5852 | -3.93(-11.77,5.54) | -0.44(-0.68,-0.20) |
| New Zealand | 11.05(9.28,13.27) | 14.02(11.99,16.47) | 1300 | 26.85(18.61,35.97) | 0.91(0.79,1.02) |
| Nicaragua | 12.99(11.38,14.93) | 18.05(15.46,21.14) | 849 | 39.01(20.43,56.23) | 1.07(1.00,1.13) |
| Niger | 6.10(5.49,6.90) | 7.12(6.40,8.07) | 476 | 16.75(7.57,26.44) | 0.52(0.49,0.55) |
| Nigeria | 7.26(6.07,8.54) | 9.16(7.81,10.70) | 6961 | 26.16(20.20,32.99) | 0.81(0.76,0.87) |
| Northern Mariana Islands | 15.53(13.50,17.92) | 17.36(14.92,20.49) | 7 | 11.84(-0.53,26.29) | 0.22(0.14,0.29) |
| Norway | 6.46(5.49,7.52) | 13.91(11.93,16.06) | 1503 | 115.31(95.10,135.18) | 2.42(2.14,2.71) |
| Oman | 14.43(12.30,16.61) | 26.98(23.43,30.55) | 528 | 86.93(61.95,123.59) | 2.03(1.99,2.06) |
| Pakistan | 9.02(7.47,10.56) | 11.33(9.70,13.07) | 14274 | 25.55(18.13,32.83) | 0.80(0.69,0.91) |
| Palestine | 13.27(11.35,15.23) | 17.19(14.88,19.77) | 427 | 29.59(14.73,47.16) | 0.76(0.72,0.79) |
| Panama | 13.87(11.78,16.17) | 19.21(16.06,22.32) | 880 | 38.51(15.76,65.25) | 1.09(1.00,1.18) |
| Papua New Guinea | 15.25(13.44,17.59) | 18.84(16.46,21.76) | 820 | 23.56(10.59,37.91) | 0.54(0.51,0.57) |
| Paraguay | 21.16(17.68,25.19) | 30.91(25.86,35.98) | 1670 | 46.04(28.72,70.62) | 1.12(1.04,1.20) |
| Peru | 13.34(11.60,14.99) | 17.94(15.32,20.71) | 6591 | 34.54(20.91,47.60) | 0.88(0.80,0.96) |
| Philippines | 8.86(7.55,10.45) | 10.57(9.10,12.12) | 8292 | 19.23(13.13,27.42) | 0.56(0.51,0.60) |
| Plurinational State of Bolivia | 17.12(14.66,19.87) | 25.09(21.87,29.07) | 2326 | 46.59(26.46,67.47) | 1.22(1.14,1.31) |
| Poland | 12.90(10.78,15.07) | 14.02(12.50,15.60) | 11059 | 8.65(-1.20,21.09) | 0.04(-0.05,0.13) |
| Portugal | 10.77(9.68,12.50) | 14.85(13.49,16.56) | 4085 | 37.90(27.71,52.19) | 1.05(0.98,1.13) |
| Principality of Monaco | 11.62(10.30,13.53) | 16.38(14.38,18.49) | 19 | 40.96(26.85,59.66) | 1.13(1.10,1.17) |
| Puerto Rico | 17.14(14.89,20.12) | 17.89(15.60,20.22) | 1633 | 4.37(-8.54,22.87) | 0.02(-0.05,0.09) |
| Qatar | 18.17(15.72,21.28) | 19.28(16.13,22.62) | 191 | 6.14(-7.93,26.59) | 0.08(-0.02,0.18) |
| Republic of CÃ´te d'Ivoire | 7.75(6.99,8.66) | 8.49(7.62,9.58) | 865 | 9.67(1.76,19.78) | 0.29(0.22,0.35) |
| Republic of Cabo Verde | 7.84(6.95,8.79) | 10.97(9.75,12.56) | 44 | 39.98(27.98,52.98) | 1.11(1.05,1.17) |
| Republic of Korea | 8.51(7.67,9.47) | 14.17(13.35,15.25) | 14707 | 66.61(48.55,85.37) | 2.00(1.77,2.23) |
| Republic of Moldova | 8.37(7.56,9.38) | 6.96(6.16,7.72) | 449 | -16.77(-26.09,-6.10) | -0.54(-0.57,-0.51) |
| Republic of Nauru | 17.84(15.91,20.38) | 19.94(17.91,23.05) | 1 | 11.80(0.82,26.21) | 0.23(0.20,0.27) |
| Republic of Niue | 16.54(14.49,18.97) | 18.68(16.57,21.44) | 0 | 12.97(3.31,26.19) | 0.30(0.21,0.39) |
| Republic of Palau | 13.69(12.00,16.03) | 17.39(15.09,20.33) | 4 | 27.00(16.86,41.92) | 0.61(0.57,0.66) |
| Republic of San Marino | 8.88(7.71,10.17) | 10.88(9.30,12.80) | 9 | 22.51(11.60,34.03) | 0.56(0.51,0.61) |
| Republic of the Gambia | 7.43(6.71,8.28) | 8.27(7.38,9.51) | 85 | 11.22(3.12,22.74) | 0.36(0.30,0.43) |
| Romania | 9.88(9.03,10.73) | 13.37(12.20,14.64) | 5385 | 35.25(25.41,46.04) | 1.01(0.89,1.13) |
| Russian Federation | 8.17(6.82,9.65) | 9.20(7.90,10.69) | 24093 | 12.67(6.87,18.53) | 0.42(0.39,0.45) |
| Rwanda | 7.70(6.93,8.53) | 8.76(7.94,9.68) | 506 | 13.79(3.25,23.27) | 0.43(0.40,0.46) |
| Saint Kitts and Nevis | 20.94(18.45,23.97) | 28.67(25.88,31.79) | 16 | 36.91(16.68,59.74) | 1.15(1.09,1.21) |
| Saint Lucia | 20.96(18.06,24.10) | 23.21(20.53,26.32) | 58 | 10.73(-2.63,25.25) | 0.19(0.13,0.26) |
| Saint Vincent and the Grenadines | 16.14(14.13,18.61) | 20.25(16.74,23.62) | 33 | 25.51(7.17,39.06) | 0.57(0.44,0.69) |
| Samoa | 14.56(12.79,16.71) | 16.55(14.44,19.52) | 27 | 13.69(1.35,29.38) | 0.25(0.19,0.31) |
| Sao Tome and Principe | 8.63(7.80,9.60) | 12.13(10.87,13.50) | 12 | 40.57(30.47,54.03) | 1.15(1.12,1.19) |
| Saudi Arabia | 20.41(17.76,23.68) | 33.39(29.01,38.47) | 3762 | 63.56(39.56,86.12) | 1.57(1.51,1.63) |
| Senegal | 6.70(5.99,7.55) | 8.49(7.62,9.59) | 674 | 26.74(13.88,40.28) | 0.73(0.69,0.78) |
| Serbia | 12.18(10.70,13.98) | 11.97(10.73,13.37) | 2115 | -1.70(-13.34,9.56) | -0.13(-0.20,-0.07) |
| Seychelles | 13.21(11.91,14.84) | 17.30(15.68,19.51) | 21 | 30.92(18.45,42.02) | 0.77(0.72,0.83) |
| Sierra Leone | 6.52(5.88,7.31) | 8.10(7.14,9.17) | 240 | 24.17(13.59,36.90) | 0.67(0.65,0.68) |
| Singapore | 6.95(6.34,7.65) | 8.43(7.60,9.49) | 782 | 21.31(11.76,37.05) | 0.53(0.44,0.61) |
| Slovakia | 9.46(8.45,10.52) | 11.80(10.48,13.41) | 1229 | 24.75(13.85,35.34) | 0.60(0.58,0.62) |
| Slovenia | 10.11(9.16,11.16) | 14.07(12.48,15.44) | 682 | 39.15(27.57,55.06) | 0.93(0.86,1.00) |
| Socialist Republic of Viet Nam | 8.44(7.55,9.59) | 12.95(11.51,14.89) | 13697 | 53.38(41.49,69.72) | 1.41(1.35,1.47) |
| Solomon Islands | 13.05(11.38,14.87) | 14.75(12.88,17.25) | 56 | 13.03(2.23,25.13) | 0.22(0.17,0.26) |
| Somalia | 7.22(6.45,8.07) | 7.72(7.02,8.43) | 607 | 6.87(-1.27,16.59) | 0.18(0.12,0.25) |
| South Africa | 7.12(5.99,8.34) | 8.86(7.61,10.36) | 4089 | 24.40(18.13,32.12) | 0.77(0.71,0.83) |
| South Sudan | 6.93(6.19,7.68) | 7.67(6.89,8.57) | 247 | 10.62(1.45,19.79) | 0.28(0.19,0.37) |
| Spain | 13.16(12.56,13.88) | 19.57(17.37,22.21) | 21275 | 48.77(34.83,63.52) | 1.56(1.34,1.77) |
| Sri Lanka | 10.46(9.27,11.87) | 13.41(12.06,15.06) | 4108 | 28.15(17.20,40.75) | 0.81(0.78,0.84) |
| Sudan | 9.81(8.54,11.39) | 14.86(12.82,17.17) | 3220 | 51.45(35.10,71.97) | 1.25(1.23,1.28) |
| Suriname | 13.50(11.81,15.92) | 20.67(17.90,23.82) | 132 | 53.14(35.41,73.60) | 1.24(1.19,1.30) |
| Sweden | 15.18(13.84,16.39) | 17.93(16.31,19.64) | 4164 | 18.13(4.44,33.04) | 0.89(0.71,1.07) |
| Switzerland | 11.86(10.72,13.49) | 14.62(12.98,16.27) | 2893 | 23.23(12.39,34.16) | 0.75(0.64,0.86) |
| Syrian Arab Republic | 15.26(13.34,17.50) | 19.47(16.63,22.30) | 2661 | 27.62(15.32,40.84) | 0.73(0.64,0.81) |
| Taiwan (Province of China) | 18.15(16.62,20.10) | 25.25(24.38,26.03) | 11498 | 39.12(25.47,52.38) | 2.17(1.51,2.83) |
| Tajikistan | 7.51(6.91,8.18) | 10.65(9.78,11.61) | 452 | 41.76(34.12,50.28) | 1.17(1.03,1.30) |
| Thailand | 10.54(9.32,11.88) | 13.45(11.95,15.39) | 15813 | 27.55(16.14,42.73) | 0.90(0.86,0.93) |
| The former Yugoslav Republic of Macedonia | 10.62(9.50,11.82) | 14.21(12.39,16.01) | 499 | 33.86(22.00,48.13) | 0.98(0.92,1.04) |
| Timor-Leste | 7.04(6.31,8.01) | 10.75(9.64,12.00) | 92 | 52.82(39.46,68.92) | 1.33(1.31,1.35) |
| Togo | 6.78(6.10,7.52) | 8.02(7.14,8.88) | 272 | 18.33(8.52,30.43) | 0.53(0.51,0.55) |
| Tokelau | 16.11(14.00,18.42) | 17.27(15.01,19.97) | 0 | 7.20(-2.92,20.26) | 0.12(0.02,0.21) |
| Tonga | 14.34(12.43,16.43) | 16.95(14.86,19.51) | 14 | 18.26(4.78,32.21) | 0.42(0.38,0.46) |
| Trinidad and Tobago | 18.79(16.43,21.37) | 21.33(18.13,25.30) | 411 | 13.52(-6.05,31.35) | 0.25(0.18,0.32) |
| Tunisia | 12.23(10.32,14.18) | 16.24(13.86,18.93) | 2143 | 32.78(15.34,52.05) | 0.71(0.65,0.78) |
| Turkey | 15.08(13.02,17.52) | 33.72(29.67,38.46) | 34043 | 123.56(95.96,156.06) | 2.17(1.97,2.37) |
| Turkmenistan | 5.16(4.84,5.50) | 6.24(5.78,6.82) | 231 | 20.94(14.41,27.16) | 0.66(0.63,0.69) |
| Tuvalu | 11.16(9.89,12.79) | 15.64(13.68,17.88) | 1 | 40.20(30.01,51.61) | 0.94(0.90,0.98) |
| Uganda | 6.71(5.94,7.62) | 7.93(7.11,8.79) | 1105 | 18.19(7.75,33.12) | 0.55(0.50,0.59) |
| Ukraine | 6.65(5.61,7.80) | 6.13(5.26,7.06) | 5032 | -7.82(-14.42,-0.95) | -0.25(-0.40,-0.10) |
| United Arab Emirates | 11.92(10.35,13.92) | 13.20(11.17,15.79) | 746 | 10.71(-3.05,25.51) | 0.24(0.16,0.32) |
| United Kingdom of Great Britain and Northern Ireland | 10.83(9.32,12.40) | 14.30(12.55,16.24) | 20627 | 32.09(26.83,37.80) | 0.86(0.77,0.95) |
| United Republic of Tanzania | 6.67(6.03,7.36) | 8.08(7.36,8.85) | 2119 | 21.21(12.46,33.70) | 0.73(0.68,0.78) |
| United States of America | 18.34(15.66,21.20) | 19.38(17.29,21.52) | 123731 | 5.70(-4.16,17.03) | 0.01(-0.16,0.17) |
| United States Virgin Islands | 19.88(16.88,23.53) | 22.73(19.09,27.00) | 48 | 14.33(-1.39,30.71) | 0.27(0.16,0.38) |
| Uruguay | 13.77(12.49,15.11) | 19.85(17.97,21.68) | 1172 | 44.20(29.70,67.69) | 1.03(0.72,1.34) |
| Uzbekistan | 4.92(4.60,5.30) | 5.80(5.33,6.35) | 1307 | 17.88(11.00,23.69) | 0.53(0.48,0.57) |
| Vanuatu | 13.01(11.48,14.77) | 14.84(13.04,17.00) | 26 | 14.05(1.86,26.35) | 0.30(0.28,0.33) |
| Yemen | 8.57(7.35,9.98) | 12.49(10.73,14.44) | 1743 | 45.80(32.54,63.78) | 1.19(1.15,1.23) |
| Zambia | 6.45(5.79,7.14) | 7.30(6.43,8.18) | 390 | 13.16(1.99,25.00) | 0.40(0.37,0.43) |
| Zimbabwe | 7.90(7.07,8.88) | 10.09(9.15,11.28) | 780 | 27.63(17.19,38.60) | 0.72(0.68,0.76) |

**Table S4:** Prevalence data for countries and territories related to Parkinson disease. It includes location-based total numbers, age-standardized rates (ASR) per 100,000 for 1990 and 2023, percentage changes, and EAPC from 1990 to 2023.

| **Regions** | **ASPR, 1990 (95% CI)** | **ASPR, 2023 (95% CI)** | **Numbers, 2023** | **PC 1990-2023** | **EAPC, %, 1990-2023** |
| --- | --- | --- | --- | --- | --- |
| Afghanistan | 86.89(71.36,107.41) | 138.23(117.73,168.26) | 13525 | 59.09(35.86,81.58) | 1.48(1.44,1.51) |
| Albania | 68.19(55.83,81.20) | 111.27(91.42,134.85) | 5111 | 63.17(40.46,90.44) | 1.43(1.39,1.47) |
| Algeria | 96.77(79.74,118.50) | 158.43(128.06,192.65) | 55082 | 63.72(42.75,91.01) | 1.40(1.35,1.45) |
| American Samoa | 128.45(109.57,150.92) | 168.62(145.88,199.57) | 67 | 31.27(14.86,49.86) | 0.62(0.54,0.69) |
| Andorra | 92.78(76.54,112.38) | 96.72(80.87,115.82) | 171 | 4.25(-10.62,19.27) | 0.06(-0.02,0.14) |
| Angola | 36.27(29.57,43.23) | 52.26(43.01,62.59) | 5675 | 44.08(22.57,71.02) | 1.24(1.20,1.29) |
| Antigua and Barbuda | 140.09(114.65,165.21) | 172.59(142.96,204.61) | 192 | 23.20(6.94,43.82) | 0.57(0.50,0.65) |
| Argentina | 117.21(99.71,132.87) | 144.92(129.35,159.49) | 94139 | 23.63(6.40,43.55) | 0.39(0.10,0.67) |
| Armenia | 29.03(24.79,34.35) | 36.19(30.30,42.44) | 1905 | 24.65(6.83,49.78) | 0.65(0.55,0.75) |
| Australia | 134.05(120.94,147.47) | 154.87(126.96,185.24) | 83726 | 15.53(-3.01,37.40) | 0.24(-0.10,0.58) |
| Austria | 81.69(70.61,93.92) | 109.22(95.54,125.88) | 22602 | 33.69(16.87,56.36) | 1.00(0.90,1.09) |
| Azerbaijan | 26.31(21.60,31.79) | 34.28(30.17,38.91) | 3645 | 30.29(14.02,49.23) | 0.69(0.61,0.78) |
| Bahrain | 130.47(109.53,159.03) | 179.88(147.08,215.29) | 1427 | 37.88(22.27,59.40) | 0.92(0.84,0.99) |
| Bangladesh | 54.13(44.34,65.19) | 82.76(66.81,101.61) | 92636 | 52.89(31.09,76.53) | 1.33(1.28,1.39) |
| Barbados | 143.54(121.09,166.72) | 189.10(155.63,226.82) | 1069 | 31.74(13.53,49.97) | 0.70(0.62,0.78) |
| Belarus | 48.99(40.09,62.51) | 52.43(43.19,62.58) | 8955 | 7.01(-6.32,22.29) | 0.46(0.37,0.55) |
| Belgium | 87.05(76.69,100.42) | 108.99(94.86,128.84) | 28307 | 25.21(13.25,37.81) | 0.80(0.73,0.88) |
| Belize | 129.07(104.64,161.07) | 190.53(158.34,226.54) | 517 | 47.61(24.14,82.13) | 1.20(1.10,1.31) |
| Benin | 37.38(31.46,45.84) | 54.42(44.68,65.01) | 2744 | 45.56(25.99,67.85) | 1.13(1.08,1.18) |
| Bermuda | 132.29(108.78,166.96) | 166.74(139.30,193.66) | 256 | 26.04(6.71,48.16) | 0.63(0.56,0.71) |
| Bhutan | 49.43(40.12,60.37) | 82.50(67.51,97.35) | 512 | 66.89(43.09,92.46) | 1.68(1.62,1.74) |
| Bolivarian Republic of Venezuela | 129.38(104.46,156.63) | 157.67(124.87,189.73) | 44600 | 21.86(1.60,38.04) | 0.61(0.41,0.80) |
| Bosnia and Herzegovina | 70.32(59.52,83.40) | 107.68(87.34,128.40) | 7034 | 53.14(31.78,84.39) | 1.44(1.39,1.50) |
| Botswana | 44.24(36.92,52.83) | 51.77(42.62,61.04) | 850 | 17.03(4.52,33.75) | 0.54(0.45,0.62) |
| Brazil | 216.29(176.67,263.50) | 283.96(229.02,346.06) | 731773 | 31.29(24.27,38.65) | 0.24(0.01,0.46) |
| Brunei Darussalam | 60.07(49.52,72.85) | 87.26(70.93,106.21) | 308 | 45.26(24.84,68.50) | 1.03(0.98,1.08) |
| Bulgaria | 107.83(93.15,122.53) | 109.09(90.57,124.80) | 18887 | 1.16(-14.54,22.54) | -0.81(-1.25,-0.36) |
| Burkina Faso | 36.94(30.53,45.73) | 47.06(39.01,55.49) | 4290 | 27.41(10.49,43.90) | 0.71(0.66,0.75) |
| Burundi | 34.29(27.59,42.83) | 46.67(37.58,56.67) | 1904 | 36.10(19.83,57.79) | 1.06(1.02,1.09) |
| Cambodia | 54.16(45.30,65.42) | 92.01(77.64,110.86) | 11888 | 69.88(48.57,90.91) | 1.68(1.63,1.74) |
| Cameroon | 43.38(36.53,52.14) | 57.77(47.44,68.41) | 6179 | 33.17(16.13,56.23) | 0.89(0.86,0.93) |
| Canada | 58.81(50.46,68.79) | 118.01(101.77,138.78) | 98356 | 100.64(77.07,133.82) | 3.45(2.79,4.12) |
| Central African Republic | 35.13(28.36,42.79) | 43.09(35.87,52.08) | 934 | 22.68(8.67,42.31) | 0.62(0.59,0.64) |
| Chad | 34.36(27.54,41.63) | 43.10(35.02,52.24) | 2211 | 25.43(8.62,49.34) | 0.74(0.70,0.79) |
| Chile | 98.85(91.55,110.40) | 126.54(111.10,152.15) | 35507 | 28.01(12.55,55.92) | 0.53(-0.02,1.08) |
| China | 106.56(86.16,131.29) | 148.74(122.07,183.52) | 3445692 | 39.59(32.33,48.31) | 1.30(1.17,1.44) |
| Colombia | 96.78(79.36,118.61) | 118.97(111.38,126.16) | 69997 | 22.94(3.01,46.80) | 0.46(0.32,0.60) |
| Commonwealth of the Bahamas | 128.21(103.38,154.23) | 179.10(152.08,210.67) | 693 | 39.69(18.17,68.41) | 0.98(0.92,1.04) |
| Comoros | 42.63(35.50,51.80) | 64.20(55.15,74.47) | 385 | 50.59(31.83,75.30) | 1.32(1.28,1.35) |
| Congo | 41.58(34.84,49.79) | 53.40(44.47,65.87) | 1160 | 28.43(10.17,48.02) | 0.84(0.81,0.87) |
| Cook Islands | 126.27(102.96,153.34) | 147.26(120.03,176.55) | 28 | 16.62(4.55,28.70) | 0.32(0.24,0.40) |
| Costa Rica | 120.45(98.83,148.22) | 169.49(141.31,202.08) | 10143 | 40.71(16.51,76.41) | 1.32(1.15,1.48) |
| Croatia | 75.95(65.89,93.06) | 110.24(91.87,132.75) | 10674 | 45.14(27.78,69.45) | 1.29(1.08,1.50) |
| Cuba | 71.59(61.87,84.08) | 154.80(127.91,185.22) | 32314 | 116.21(83.12,146.74) | 2.47(2.03,2.91) |
| Cyprus | 99.81(84.26,118.35) | 113.51(94.36,132.52) | 2267 | 13.73(0.27,31.42) | 0.34(0.31,0.37) |
| Czech Republic | 71.38(60.00,85.59) | 112.00(92.90,125.94) | 27251 | 56.92(42.40,77.76) | 1.49(1.41,1.57) |
| Democratic People's Republic of Korea | 94.75(76.89,116.67) | 133.79(111.34,162.40) | 41493 | 41.20(25.03,59.96) | 1.10(1.05,1.14) |
| Democratic Republic of the Congo | 34.43(27.72,41.65) | 44.46(36.34,52.84) | 14687 | 29.14(9.25,49.01) | 0.81(0.75,0.87) |
| Denmark | 52.82(48.51,57.09) | 95.46(86.63,108.12) | 12928 | 80.74(66.28,104.53) | 1.61(1.53,1.70) |
| Djibouti | 36.10(29.66,43.18) | 55.39(45.69,65.76) | 315 | 53.44(32.01,77.48) | 1.40(1.36,1.44) |
| Dominica | 152.19(128.23,184.09) | 204.90(168.62,237.69) | 217 | 34.63(15.81,61.08) | 0.76(0.70,0.83) |
| Dominican Republic | 134.30(111.79,161.21) | 202.98(168.36,238.51) | 18996 | 51.14(34.45,75.20) | 1.13(0.96,1.31) |
| Ecuador | 131.60(108.68,161.86) | 222.80(187.02,263.03) | 39829 | 69.30(38.46,111.05) | 1.49(1.39,1.59) |
| Egypt | 112.66(91.46,136.46) | 183.74(148.19,224.54) | 99642 | 63.09(38.70,96.70) | 2.02(1.71,2.32) |
| El Salvador | 114.29(94.96,139.97) | 167.72(137.12,202.80) | 10297 | 46.74(24.82,72.17) | 1.28(1.20,1.36) |
| Equatorial Guinea | 33.08(27.11,40.51) | 55.12(45.29,65.35) | 238 | 66.63(44.33,92.36) | 1.75(1.60,1.91) |
| Eritrea | 33.87(27.64,41.86) | 52.58(44.03,63.37) | 1639 | 55.23(34.93,80.84) | 1.36(1.27,1.45) |
| Estonia | 95.26(87.42,101.84) | 107.72(98.39,118.27) | 3360 | 13.09(4.28,26.58) | 0.27(0.17,0.38) |
| Ethiopia | 42.56(34.48,52.87) | 57.28(46.25,70.74) | 23655 | 34.60(24.62,45.33) | 0.98(0.92,1.05) |
| Federated States of Micronesia | 100.89(84.25,120.93) | 158.29(131.60,186.81) | 97 | 56.89(35.86,82.80) | 1.29(1.20,1.39) |
| Fiji | 113.44(94.88,136.02) | 141.17(119.32,169.22) | 942 | 24.44(3.65,43.80) | 0.53(0.47,0.59) |
| Finland | 72.06(65.91,78.23) | 110.60(97.88,123.62) | 15854 | 53.49(38.85,68.61) | 1.36(1.21,1.51) |
| France | 101.95(89.41,113.26) | 116.58(108.70,124.24) | 186712 | 14.35(3.81,26.19) | 0.52(0.47,0.58) |
| Gabon | 47.81(39.88,58.07) | 62.94(52.41,74.41) | 689 | 31.64(12.73,57.22) | 0.82(0.78,0.86) |
| Georgia | 30.54(25.35,35.97) | 42.54(35.67,51.89) | 2656 | 39.28(19.16,64.74) | 1.04(1.01,1.07) |
| Germany | 80.59(70.43,91.33) | 197.84(187.34,209.66) | 447635 | 145.49(120.47,173.89) | 2.18(1.85,2.50) |
| Ghana | 37.66(31.06,45.87) | 55.44(46.28,66.61) | 8291 | 47.21(28.52,71.81) | 1.13(1.10,1.16) |
| Greece | 67.09(57.28,78.35) | 136.04(119.95,152.45) | 38991 | 102.78(69.93,137.95) | 2.65(2.16,3.15) |
| Greenland | 52.72(41.92,65.38) | 100.11(81.22,119.33) | 58 | 89.89(56.56,124.72) | 2.06(2.02,2.10) |
| Grenada | 133.88(107.16,163.60) | 193.25(163.42,228.19) | 319 | 44.35(20.26,66.05) | 1.13(1.04,1.22) |
| Guam | 112.36(93.31,136.00) | 139.52(117.53,166.53) | 285 | 24.17(7.74,44.11) | 0.48(0.41,0.55) |
| Guatemala | 107.95(90.38,134.87) | 172.46(136.64,208.87) | 18094 | 59.75(34.92,90.81) | 1.29(1.24,1.34) |
| Guinea | 33.41(27.77,39.73) | 44.37(36.62,53.08) | 2450 | 32.82(13.38,53.20) | 0.85(0.79,0.90) |
| Guinea-Bissau | 39.20(32.30,47.44) | 50.21(41.68,59.47) | 348 | 28.07(10.50,46.03) | 0.70(0.64,0.75) |
| Guyana | 128.45(103.08,151.38) | 161.48(133.91,194.43) | 1043 | 25.72(7.20,52.18) | 0.45(0.35,0.54) |
| Haiti | 95.62(78.51,117.29) | 136.88(112.71,162.80) | 8203 | 43.15(26.04,67.70) | 0.98(0.91,1.05) |
| Honduras | 130.32(106.03,161.65) | 181.33(150.84,212.41) | 11910 | 39.14(22.52,60.75) | 1.09(0.99,1.19) |
| Hungary | 85.62(75.48,95.75) | 122.14(116.15,129.27) | 25501 | 42.64(28.78,59.49) | 0.10(-0.24,0.44) |
| Iceland | 94.84(81.77,111.94) | 143.82(121.01,180.29) | 923 | 51.64(31.86,78.01) | 1.21(1.12,1.30) |
| India | 42.66(34.33,52.87) | 67.44(55.11,82.69) | 778881 | 58.09(50.35,66.77) | 1.34(1.21,1.47) |
| Indonesia | 56.90(45.39,70.11) | 88.03(72.72,108.56) | 202140 | 54.70(47.01,62.98) | 1.31(1.23,1.39) |
| Iraq | 118.92(96.64,146.59) | 197.49(165.27,236.75) | 41828 | 66.07(39.79,94.21) | 1.64(1.60,1.67) |
| Ireland | 93.78(80.38,111.99) | 109.41(94.49,125.31) | 9842 | 16.67(4.38,29.31) | 0.66(0.58,0.74) |
| Islamic Republic of Iran | 96.95(79.41,119.39) | 161.11(133.08,196.62) | 116554 | 66.19(57.69,74.11) | 1.76(1.70,1.82) |
| Israel | 145.61(131.22,169.24) | 153.95(135.26,184.43) | 20999 | 5.73(-5.51,20.71) | 1.09(0.79,1.39) |
| Italy | 181.18(152.95,212.96) | 215.66(178.46,263.38) | 341460 | 19.03(11.28,28.09) | 0.73(0.52,0.94) |
| Jamaica | 119.32(97.45,138.38) | 152.89(123.94,185.75) | 5042 | 28.14(12.61,53.08) | 0.63(0.53,0.73) |
| Japan | 61.32(49.64,74.84) | 88.20(71.73,108.06) | 377198 | 43.84(36.13,52.16) | -0.04(-1.00,0.94) |
| Jordan | 94.19(77.44,113.15) | 125.10(102.64,153.23) | 9204 | 32.82(14.84,53.19) | 0.63(0.17,1.09) |
| Kazakhstan | 30.10(24.89,36.65) | 41.92(34.77,51.03) | 8289 | 39.26(20.35,59.46) | 1.16(1.08,1.23) |
| Kenya | 27.34(21.99,34.13) | 37.26(30.36,45.18) | 8725 | 36.28(29.69,42.99) | 0.93(0.86,1.01) |
| Kingdom of Eswatini | 41.20(33.99,50.80) | 51.14(42.34,60.89) | 352 | 24.12(12.38,39.65) | 0.69(0.60,0.77) |
| Kiribati | 115.57(96.14,139.45) | 147.50(124.91,177.94) | 94 | 27.62(13.38,43.81) | 0.58(0.50,0.66) |
| Kuwait | 159.77(131.17,199.91) | 181.08(152.17,215.37) | 4185 | 13.34(-7.89,34.31) | 0.25(0.18,0.33) |
| Kyrgyzstan | 23.14(19.53,27.81) | 32.25(27.24,39.57) | 1591 | 39.36(19.68,59.92) | 1.09(1.02,1.17) |
| Lao People's Democratic Republic | 46.29(37.93,55.54) | 80.45(67.25,96.44) | 3649 | 73.80(51.91,102.12) | 1.81(1.75,1.86) |
| Latvia | 51.61(44.28,59.00) | 61.54(51.76,69.46) | 2775 | 19.25(7.76,31.65) | 0.74(0.62,0.86) |
| Lebanon | 107.06(88.46,131.46) | 146.96(123.88,175.96) | 9213 | 37.27(14.04,59.74) | 0.96(0.93,1.00) |
| Lesotho | 32.17(26.27,39.07) | 44.12(36.47,51.57) | 536 | 37.15(19.28,56.64) | 0.97(0.92,1.01) |
| Liberia | 41.60(33.99,49.96) | 53.85(43.87,66.23) | 1079 | 29.44(13.01,51.29) | 0.80(0.73,0.88) |
| Libya | 88.03(75.29,104.46) | 162.00(133.23,198.65) | 8009 | 84.02(54.33,115.25) | 1.92(1.66,2.17) |
| Lithuania | 46.79(38.98,53.34) | 59.67(49.57,69.54) | 3876 | 27.52(12.44,45.46) | 0.80(0.75,0.86) |
| Luxembourg | 95.63(77.82,113.62) | 118.09(99.00,135.40) | 1319 | 23.49(10.09,38.99) | 0.69(0.64,0.74) |
| Madagascar | 29.44(24.45,35.52) | 39.07(32.03,48.67) | 3817 | 32.72(14.74,53.75) | 0.87(0.82,0.91) |
| Malawi | 29.54(24.02,36.16) | 39.43(32.17,46.86) | 2753 | 33.49(15.92,55.76) | 0.89(0.86,0.93) |
| Malaysia | 74.17(61.80,89.83) | 100.93(83.85,120.52) | 30973 | 36.08(15.97,56.25) | 0.96(0.91,1.01) |
| Maldives | 73.96(61.21,90.62) | 110.95(93.72,130.34) | 325 | 50.02(29.25,71.99) | 1.32(1.25,1.39) |
| Mali | 39.58(32.74,47.79) | 52.28(43.62,62.72) | 4246 | 32.09(14.29,53.28) | 0.83(0.75,0.90) |
| Malta | 99.92(83.22,117.35) | 121.00(100.83,144.03) | 1351 | 21.10(0.25,40.11) | 0.58(0.52,0.64) |
| Marshall Islands | 119.79(100.15,144.35) | 164.58(136.99,199.22) | 32 | 37.39(18.77,56.74) | 0.77(0.71,0.84) |
| Mauritania | 40.55(33.98,49.35) | 53.19(43.47,64.97) | 1051 | 31.16(14.93,47.76) | 0.74(0.71,0.78) |
| Mauritius | 86.50(75.89,100.79) | 118.48(100.40,135.28) | 2152 | 36.97(23.26,52.46) | 0.92(0.82,1.03) |
| Mexico | 148.54(120.58,178.39) | 182.53(148.95,223.32) | 240180 | 22.89(15.84,29.56) | 0.49(0.39,0.58) |
| Mongolia | 23.41(19.15,28.50) | 27.83(24.85,31.07) | 619 | 18.84(3.46,34.31) | 0.41(0.32,0.49) |
| Montenegro | 77.72(64.13,91.70) | 109.83(88.78,132.39) | 1267 | 41.31(21.90,64.26) | 1.20(1.15,1.25) |
| Morocco | 90.26(72.32,109.52) | 150.10(123.64,180.29) | 50705 | 66.29(44.36,90.75) | 1.53(1.50,1.57) |
| Mozambique | 32.83(26.86,39.83) | 43.14(35.33,51.73) | 5019 | 31.39(17.27,48.86) | 0.77(0.70,0.83) |
| Myanmar | 52.28(42.77,64.18) | 82.57(67.00,100.48) | 37919 | 57.95(40.12,80.92) | 1.44(1.40,1.48) |
| Namibia | 38.35(31.46,46.37) | 52.09(43.24,62.13) | 788 | 35.82(19.48,55.67) | 1.04(0.97,1.10) |
| Nepal | 43.26(35.33,53.63) | 74.36(61.62,87.76) | 17882 | 71.91(48.57,102.76) | 1.77(1.71,1.83) |
| Netherlands | 115.94(107.47,124.21) | 111.22(99.73,128.20) | 44765 | -4.07(-13.40,9.25) | -0.52(-0.86,-0.17) |
| New Zealand | 83.33(67.48,102.74) | 108.17(87.41,134.83) | 10212 | 29.80(20.94,40.58) | 1.05(0.85,1.25) |
| Nicaragua | 111.91(93.62,132.94) | 161.47(134.29,193.83) | 7272 | 44.28(21.18,66.92) | 1.19(1.13,1.25) |
| Niger | 33.36(27.61,40.97) | 44.04(36.63,53.09) | 2994 | 32.03(14.67,51.47) | 0.88(0.84,0.92) |
| Nigeria | 37.79(30.47,46.91) | 51.41(41.91,63.06) | 40515 | 36.03(28.04,43.78) | 1.05(1.00,1.11) |
| Northern Mariana Islands | 117.65(98.51,139.55) | 147.58(122.81,175.61) | 56 | 25.44(6.53,49.35) | 0.54(0.47,0.62) |
| Norway | 30.94(25.51,38.28) | 114.66(94.56,140.94) | 12563 | 270.60(238.16,305.78) | 3.59(3.02,4.17) |
| Oman | 114.66(92.44,142.43) | 215.58(176.35,253.24) | 3915 | 88.02(57.15,137.45) | 1.94(1.90,1.97) |
| Pakistan | 57.51(46.14,70.40) | 85.77(70.46,104.80) | 107236 | 49.13(40.03,59.55) | 1.41(1.27,1.55) |
| Palestine | 106.26(86.84,127.63) | 153.92(130.38,180.47) | 3622 | 44.85(23.71,71.78) | 1.11(1.07,1.14) |
| Panama | 119.60(98.27,144.54) | 171.84(135.69,200.75) | 7902 | 43.68(15.58,78.40) | 1.17(1.09,1.25) |
| Papua New Guinea | 107.50(89.46,130.88) | 146.47(120.92,177.84) | 6066 | 36.24(17.32,58.24) | 0.79(0.74,0.83) |
| Paraguay | 197.39(158.21,239.90) | 291.70(236.96,344.58) | 14784 | 47.78(28.12,78.27) | 1.16(1.07,1.25) |
| Peru | 108.77(89.34,127.82) | 160.70(130.64,190.30) | 58428 | 47.74(29.07,69.55) | 1.13(1.00,1.27) |
| Philippines | 63.99(51.70,78.65) | 82.52(67.86,102.14) | 64620 | 28.96(21.71,36.51) | 0.80(0.75,0.86) |
| Plurinational State of Bolivia | 148.57(120.40,181.24) | 234.25(196.36,276.44) | 20480 | 57.67(29.22,88.85) | 1.46(1.37,1.54) |
| Poland | 99.43(79.51,122.02) | 112.86(100.89,125.75) | 90074 | 13.52(0.18,33.44) | 0.05(-0.07,0.17) |
| Portugal | 86.25(75.48,101.17) | 123.64(109.36,141.44) | 34947 | 43.35(31.78,59.40) | 1.13(1.05,1.22) |
| Principality of Monaco | 84.86(70.73,102.50) | 119.27(96.75,140.16) | 139 | 40.54(22.91,63.91) | 1.07(1.05,1.08) |
| Puerto Rico | 147.74(123.64,175.69) | 164.64(141.07,187.22) | 15670 | 11.44(-3.01,29.69) | 0.25(0.15,0.34) |
| Qatar | 145.62(119.97,181.36) | 177.35(144.54,215.63) | 1505 | 21.79(3.14,51.27) | 0.50(0.40,0.61) |
| Republic of CÃ´te d'Ivoire | 41.85(34.42,51.87) | 52.70(43.60,62.63) | 5540 | 25.93(11.67,43.88) | 0.71(0.65,0.76) |
| Republic of Cabo Verde | 45.67(37.41,54.60) | 69.17(56.48,82.67) | 282 | 51.47(30.17,72.80) | 1.32(1.29,1.36) |
| Republic of Korea | 56.42(47.15,67.80) | 119.35(112.85,128.29) | 124070 | 111.55(77.64,154.02) | 3.35(2.89,3.82) |
| Republic of Moldova | 50.28(41.41,61.62) | 49.41(39.23,58.48) | 3146 | -1.73(-16.71,20.26) | 0.05(0.01,0.09) |
| Republic of Nauru | 124.71(102.32,151.64) | 160.24(135.82,192.45) | 6 | 28.49(10.97,50.70) | 0.61(0.54,0.67) |
| Republic of Niue | 123.62(103.23,147.18) | 156.60(132.05,182.70) | 4 | 26.68(11.59,46.12) | 0.65(0.55,0.75) |
| Republic of Palau | 109.11(88.66,133.95) | 151.67(125.04,183.13) | 29 | 39.01(23.22,60.22) | 0.87(0.82,0.93) |
| Republic of San Marino | 76.20(63.00,90.10) | 93.96(78.04,112.63) | 82 | 23.30(8.51,37.49) | 0.56(0.50,0.62) |
| Republic of the Gambia | 40.75(32.72,49.16) | 50.89(41.47,61.60) | 528 | 24.88(10.16,42.52) | 0.74(0.68,0.79) |
| Romania | 67.22(56.87,77.12) | 104.55(86.92,118.85) | 42649 | 55.53(37.92,78.52) | 1.44(1.27,1.61) |
| Russian Federation | 47.19(38.26,58.16) | 57.20(45.86,69.06) | 148666 | 21.21(15.14,27.60) | 0.68(0.63,0.74) |
| Rwanda | 32.52(26.48,40.13) | 45.45(36.73,55.29) | 2697 | 39.78(20.80,57.14) | 1.12(1.07,1.16) |
| Saint Kitts and Nevis | 155.32(126.31,189.22) | 228.85(195.54,264.09) | 120 | 47.34(17.13,88.06) | 1.40(1.34,1.47) |
| Saint Lucia | 166.67(137.26,199.51) | 190.63(161.62,224.79) | 473 | 14.38(-0.73,31.45) | 0.27(0.19,0.36) |
| Saint Vincent and the Grenadines | 134.89(112.05,159.17) | 179.18(142.91,211.19) | 300 | 32.84(13.66,51.61) | 0.72(0.61,0.84) |
| Samoa | 113.66(95.36,136.02) | 144.97(119.91,170.01) | 232 | 27.55(10.23,46.73) | 0.58(0.51,0.65) |
| Sao Tome and Principe | 45.89(38.24,55.28) | 69.09(56.87,82.60) | 72 | 50.56(30.79,75.19) | 1.35(1.30,1.39) |
| Saudi Arabia | 141.82(113.80,177.28) | 256.04(203.84,307.43) | 27123 | 80.54(45.52,114.19) | 1.84(1.78,1.89) |
| Senegal | 39.82(32.58,48.21) | 55.64(46.11,66.89) | 4435 | 39.72(15.80,62.74) | 0.98(0.92,1.03) |
| Serbia | 81.32(67.14,101.61) | 100.06(83.64,116.36) | 17503 | 23.05(3.14,42.41) | 0.55(0.50,0.59) |
| Seychelles | 90.38(75.59,108.72) | 137.53(118.87,164.55) | 164 | 52.18(31.87,69.77) | 1.23(1.19,1.27) |
| Sierra Leone | 35.79(29.67,43.28) | 49.44(39.33,61.09) | 1497 | 38.15(19.40,64.28) | 0.92(0.87,0.96) |
| Singapore | 44.27(36.37,53.22) | 61.05(51.04,72.68) | 5589 | 37.90(21.36,63.90) | 0.78(0.63,0.93) |
| Slovakia | 66.96(55.15,78.60) | 94.82(77.48,114.29) | 9858 | 41.62(21.34,60.41) | 0.94(0.90,0.98) |
| Slovenia | 75.16(61.39,86.72) | 112.97(93.38,128.41) | 5673 | 50.30(33.94,79.23) | 1.11(1.05,1.18) |
| Socialist Republic of Viet Nam | 63.08(52.77,75.47) | 101.16(83.25,122.58) | 106304 | 60.37(41.17,82.53) | 1.53(1.46,1.59) |
| Solomon Islands | 94.54(76.74,114.67) | 120.12(99.32,145.74) | 432 | 27.06(10.30,47.44) | 0.54(0.47,0.60) |
| Somalia | 33.95(27.61,41.50) | 41.17(33.94,48.93) | 3301 | 21.26(7.60,37.83) | 0.58(0.54,0.61) |
| South Africa | 41.83(34.21,51.76) | 54.36(44.75,65.85) | 25327 | 29.94(22.45,37.49) | 0.89(0.85,0.93) |
| South Sudan | 32.65(26.87,40.20) | 41.99(34.35,51.62) | 1405 | 28.58(12.95,48.01) | 0.74(0.66,0.82) |
| Spain | 96.43(91.00,103.31) | 169.60(143.95,195.11) | 192762 | 75.87(53.61,101.20) | 2.22(1.89,2.56) |
| Sri Lanka | 80.46(66.00,97.59) | 113.27(96.23,131.67) | 34407 | 40.79(24.50,61.18) | 1.09(1.06,1.13) |
| Sudan | 83.38(67.49,103.37) | 141.20(116.24,166.24) | 28581 | 69.34(45.96,98.02) | 1.56(1.53,1.59) |
| Suriname | 115.50(96.24,142.69) | 182.21(153.79,214.38) | 1135 | 57.75(36.27,83.85) | 1.32(1.25,1.38) |
| Sweden | 109.49(99.28,120.44) | 163.89(146.10,182.91) | 40573 | 49.69(32.52,68.61) | 1.62(1.39,1.85) |
| Switzerland | 91.31(79.26,108.09) | 117.07(98.74,132.21) | 23642 | 28.21(12.16,46.98) | 0.84(0.71,0.98) |
| Syrian Arab Republic | 113.88(94.27,137.83) | 169.42(139.07,200.35) | 22049 | 48.77(31.24,68.69) | 1.17(1.07,1.27) |
| Taiwan (Province of China) | 159.00(143.17,180.29) | 239.85(232.60,248.10) | 109957 | 50.85(33.77,67.61) | 3.38(2.30,4.47) |
| Tajikistan | 28.96(24.39,34.92) | 47.64(39.62,56.36) | 2365 | 64.51(45.86,87.52) | 1.59(1.50,1.68) |
| Thailand | 75.82(62.59,91.41) | 108.94(90.28,127.80) | 127930 | 43.69(25.54,67.09) | 1.36(1.27,1.46) |
| The former Yugoslav Republic of Macedonia | 75.13(62.32,90.28) | 113.23(91.04,134.19) | 3936 | 50.71(31.10,76.85) | 1.33(1.26,1.40) |
| Timor-Leste | 49.71(40.99,61.21) | 88.84(74.71,104.73) | 755 | 78.71(56.67,109.87) | 1.74(1.73,1.76) |
| Togo | 34.82(28.61,42.48) | 45.88(37.23,54.66) | 1604 | 31.74(13.99,52.38) | 0.83(0.80,0.86) |
| Tokelau | 123.55(102.73,149.69) | 148.18(123.46,174.84) | 3 | 19.93(5.31,37.10) | 0.46(0.35,0.58) |
| Tonga | 111.06(92.06,132.25) | 143.67(122.16,172.40) | 113 | 29.36(11.88,48.31) | 0.65(0.60,0.69) |
| Trinidad and Tobago | 151.66(125.74,179.40) | 185.04(152.53,230.26) | 3472 | 22.01(0.58,46.46) | 0.41(0.34,0.48) |
| Tunisia | 95.76(75.60,116.20) | 148.71(120.32,178.13) | 18965 | 55.30(32.58,82.15) | 1.18(1.11,1.26) |
| Turkey | 117.10(95.33,142.11) | 341.67(293.78,396.48) | 327447 | 191.78(146.37,239.78) | 2.89(2.64,3.15) |
| Turkmenistan | 23.55(19.78,27.90) | 36.07(30.47,43.45) | 1431 | 53.15(34.67,71.75) | 1.45(1.39,1.52) |
| Tuvalu | 83.16(70.03,100.67) | 132.06(108.56,159.42) | 10 | 58.80(43.06,75.36) | 1.30(1.26,1.34) |
| Uganda | 33.15(27.04,41.55) | 45.03(36.45,53.96) | 6395 | 35.81(14.98,57.90) | 0.97(0.92,1.02) |
| Ukraine | 37.74(30.27,46.71) | 26.59(21.99,32.06) | 21664 | -29.55(-35.14,-23.84) | -1.17(-1.55,-0.79) |
| United Arab Emirates | 100.04(82.03,120.71) | 131.44(107.60,158.23) | 6390 | 31.39(14.02,51.60) | 0.76(0.68,0.84) |
| United Kingdom of Great Britain and Northern Ireland | 87.55(72.38,104.14) | 116.37(97.22,139.07) | 169246 | 32.91(27.15,38.66) | 0.81(0.67,0.96) |
| United Republic of Tanzania | 32.14(26.32,37.92) | 44.28(36.38,52.63) | 11781 | 37.76(19.46,58.96) | 1.28(1.17,1.39) |
| United States of America | 167.93(138.17,200.54) | 154.57(137.71,173.13) | 998267 | -7.95(-16.04,4.52) | -0.53(-0.78,-0.28) |
| United States Virgin Islands | 159.10(129.25,195.38) | 195.69(160.51,240.53) | 421 | 23.00(3.78,44.66) | 0.50(0.40,0.60) |
| Uruguay | 100.47(83.79,115.20) | 156.03(136.38,171.22) | 9714 | 55.29(31.64,85.95) | 1.16(0.78,1.54) |
| Uzbekistan | 23.40(19.37,28.86) | 34.96(29.16,42.27) | 8574 | 49.37(29.30,66.82) | 1.29(1.25,1.34) |
| Vanuatu | 95.71(79.07,115.92) | 121.94(103.51,142.39) | 207 | 27.41(10.26,46.27) | 0.59(0.55,0.63) |
| Yemen | 71.00(57.54,85.52) | 115.23(95.99,135.47) | 14935 | 62.29(43.08,87.15) | 1.50(1.48,1.53) |
| Zambia | 33.84(27.44,40.31) | 45.02(35.64,54.67) | 2474 | 33.04(12.35,58.13) | 0.93(0.90,0.97) |
| Zimbabwe | 42.17(33.95,50.55) | 48.89(41.14,59.18) | 3899 | 15.93(0.40,30.37) | 0.36(0.29,0.42) |
